# The effect of type 2 diabetes genetic predisposition on non-cardiovascular comorbidities

**DOI:** 10.1101/2025.05.05.25326966

**Authors:** Ana Luiza Arruda, Ozvan Bocher, Henry J. Taylor, Davis Cammann, Satoshi Yoshiji, Xianyong Yin, Chi Zhao, Jingchun Chen, Alexis C. Wood, Ken Suzuki, Josep M. Mercader, Cassandra N. Spracklen, James B. Meigs, Marijana Vujkovic, George Davey Smith, Jerome I. Rotter, Benjamin F. Voight, Andrew P. Morris, Eleftheria Zeggini

## Abstract

Type 2 diabetes (T2D) is epidemiologically associated with a wide range of non-cardiovascular comorbidities, yet their shared etiology has not been fully elucidated. Leveraging eight non-overlapping mechanistic clusters of T2D genetic profiles, each representing distinct biological pathways, we investigate putative causal links between cluster-stratified T2D genetic predisposition and 21 non-cardiovascular comorbidities. Most of the identified putative causal effects are driven by distinct T2D genetic clusters. For example, the risk-increasing effects of T2D genetic predisposition on cataracts and erectile dysfunction are primarily attributed to obesity and glucose regulation mechanisms, respectively. When surveyed in populations across the globe, we observe opposing effect directions for depression, asthma and chronic obstructive pulmonary disease between populations. We identify a putative causal link between T2D genetic predisposition and osteoarthritis. To underscore the translational potential of our findings, we intersect high-confidence effector genes for osteoarthritis with targets of T2D-approved drugs and identify metformin as a potential candidate for drug repurposing in osteoarthritis.

## Introduction

Worldwide, over 500 million individuals have type 2 diabetes (T2D), a number expected to exceed 1.27 billion by 2050[1]. In 2022, diabetes-related healthcare expenditures were estimated to be over $412 billion in the United States of America alone[2]. The cumulative economic strain on healthcare resources is much greater when considering the comorbidities associated with T2D. For instance, a study looking at multimorbid T2D patients found that 73% also suffered from hypertension, followed by 69% with a diagnosis of back pain, 67% of depression, 45% of asthma, and 36% of osteoarthritis[3]. While advances in treatment, such as improved blood sugar monitoring[4] and glucagon-like peptide-1 (GLP-1) receptor agonists[5], can mitigate some of the challenges associated with T2D, its rising prevalence and diverse comorbidities call for a paradigm shift toward prevention strategies. Despite their prevalence and significant impact on quality of life[6], non-cardiovascular T2D comorbidities remain understudied compared to cardiovascular conditions. Building on the understanding of shared etiology, we can identify common biological mechanisms to develop targeted interventions tailored to an individual’s genetic predisposition to multimorbidity profiles[7, 8].

Genetic and environmental factors are involved in the pathophysiology of T2D, a biologically heterogeneous disease characterized by the interplay of different cell-type specific mechanisms[9]. Efforts to identify different T2D genetic profiles have yielded promising results[10, 11]. The T2D Global Genomics Initiative (T2DGGI) has identified 1,289 genetic risk variants in a multi-ancestry genome-wide association study (GWAS) meta-analysis [11] and grouped these variants into eight non-overlapping mechanistic clusters representing distinct biological pathways. Clustering was performed based on the association profile of the T2D genetic risk variants with cardiometabolic traits, yielding the following clusters: body fat (n=233 variants), obesity (n=233), beta cell associated with increased proinsulin (PI) levels (n=91), beta cell associated with decreased PI levels (n=89), metabolic syndrome (n=166), lipodystrophy (n=45), liver/lipid metabolism (n=3) and residual glycaemic (n=389).

Associations between the T2D partitioned genetic risk scores (GRS) and cardiovascular outcomes, including coronary artery disease and diabetic retinopathy, have now been described[10, 11]. However, previous studies investigating the causal links between genetic predisposition to T2D and its non-cardiovascular comorbidities have not considered genetic subtypes of disease heterogeneity[12–20]. Here, we apply a cluster-stratified causal inference analysis framework using Mendelian randomization (MR) to 21 diseases epidemiologically associated with T2D, spanning five classes: musculoskeletal, respiratory, reproductive, neuropsychiatric, and ophthalmic conditions. We identify causal links between the genetic burden of T2D mechanistic subtypes and 15 commonly co-occurring diseases and highlight repurposing opportunities for T2D-approved drugs. Our findings demonstrate the potential of translational genomics to identify targeted preventive strategies.

## Results

### Overview of the conceptual framework of the study

We investigate potential causal effects between T2D genetic predisposition and 21 non-cardiovascular comorbidities (SupTab1) using MR. The T2D comorbidities were selected after a comprehensive literature review of epidemiological studies and the availability of GWAS data. In addition to performing pairwise bi-directional two-sample MR analyses (Figure 1), we investigated cluster-stratified effects of T2D genetic predisposition on the comorbidities by restricting the genetic instrumental variables (IVs) to T2D genetic risk variants assigned to each mechanistic cluster described in the latest T2DGGI GWAS meta-analysis (Figure 1). We performed all analyses within each genetic ancestry group and meta-analyzed the single ancestry estimates where possible. Although genetic ancestry is on a continuous scale, here we use the continental genetic ancestry groups defined by the 1000 Genomes Project phase 3[21]. We corrected for multiple testing using false-discovery-rate (FDR) across all MR analyses and defined relationships showing an FDR-adjusted p-value (q-value) < 0.05 as putative causal. We applied multiple sensitivity analyses to assess the validity of the MR assumptions, including adjusting the estimates for potential confounders or mediators of the identified potentially causal relationships through multivariable MR (Methods, Supplemental Note, Figure 1). Causal estimates are expressed as the odds ratio (OR) for each comorbidity per 2-fold increase in genetically determined dichotomous T2D risk. Finally, to validate the MR findings, we performed a phenome-wide association study (PheWAS) for each T2D genetic cluster using data from the All of Us program, an ancestrally and culturally diverse cohort that was not included in the T2DGGI GWAS meta-analysis[22]. Similarly to the MR analysis, we meta-analyzed the PheWAS results across genetic ancestry groups (Methods).

**Figure 1:**
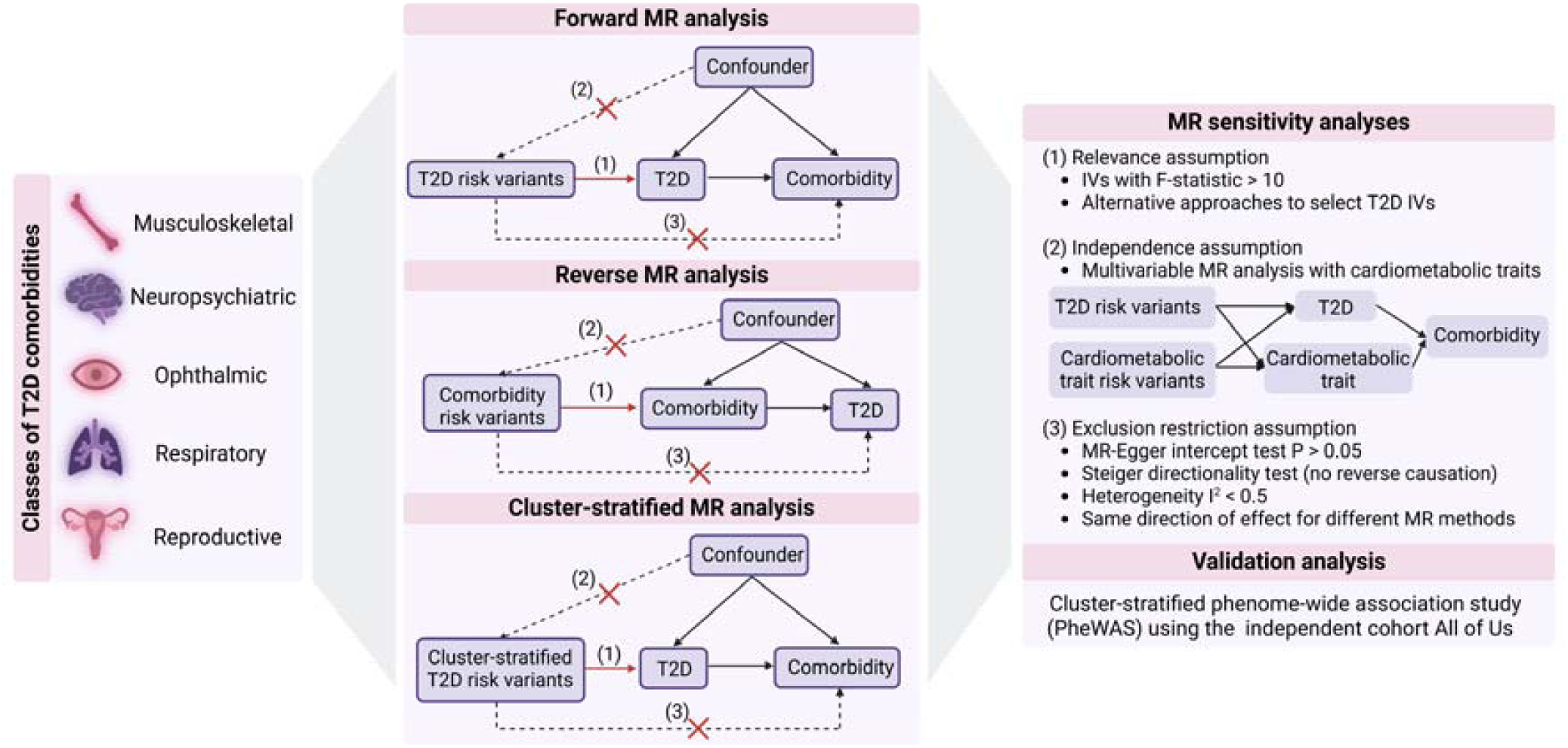
Overview of study design (MR = Mendelian randomization; T2D = type 2 diabetes; IVs = genetic instrumental variables; P = p-value).

### Genetic predisposition to T2D is a potential driver of comorbidities

We find statistical evidence (q-value < 0.05) of a putative causal link between overall T2D genetic predisposition and 10 comorbidities (Figure 2, SupTab2). Our findings show that genetic predisposition to T2D has risk-increasing effects on carpal tunnel syndrome (CTS) (OR=1.10, q-value=3.12×10^-28^), osteoarthritis (OR=1.01, q-value=4.43×10^-2^), attention-deficit/hyperactivity disorder (ADHD) (OR=1.09, q-value=5.26×10^-14^), primary open-angle glaucoma (glaucoma) (OR=1.07, q-value=6.23×10^-8^), cataracts (OR=1.04, q-value=5.82×10^-9^), polycystic ovary syndrome (PCOS) (OR=1.12, q-value=1.43×10^-5^), and erectile dysfunction (OR=1.09, q-value=7.95×10^-8^). For anorexia nervosa (OR=0.96, q-value=1.72×10^-2^), osteoporosis (OR=0.95, q-value=1.15×10^-3^), and obsessive-compulsive disorder (OCD) (OR=0.90, q-value=2.57×10^-3^), we find T2D genetic predisposition to have a protective effect. In the reverse direction, we find statistical evidence of a protective effect of genetic predisposition to bipolar disorder (OR=0.96, q-value=7.95×10^-3^) and clinically diagnosed Alzheimer’s disease (OR=0.98, q-value=4.33×10^-2^) on risk for T2D (Supplemental Figure 1, SupTab3).

**Figure 2:**
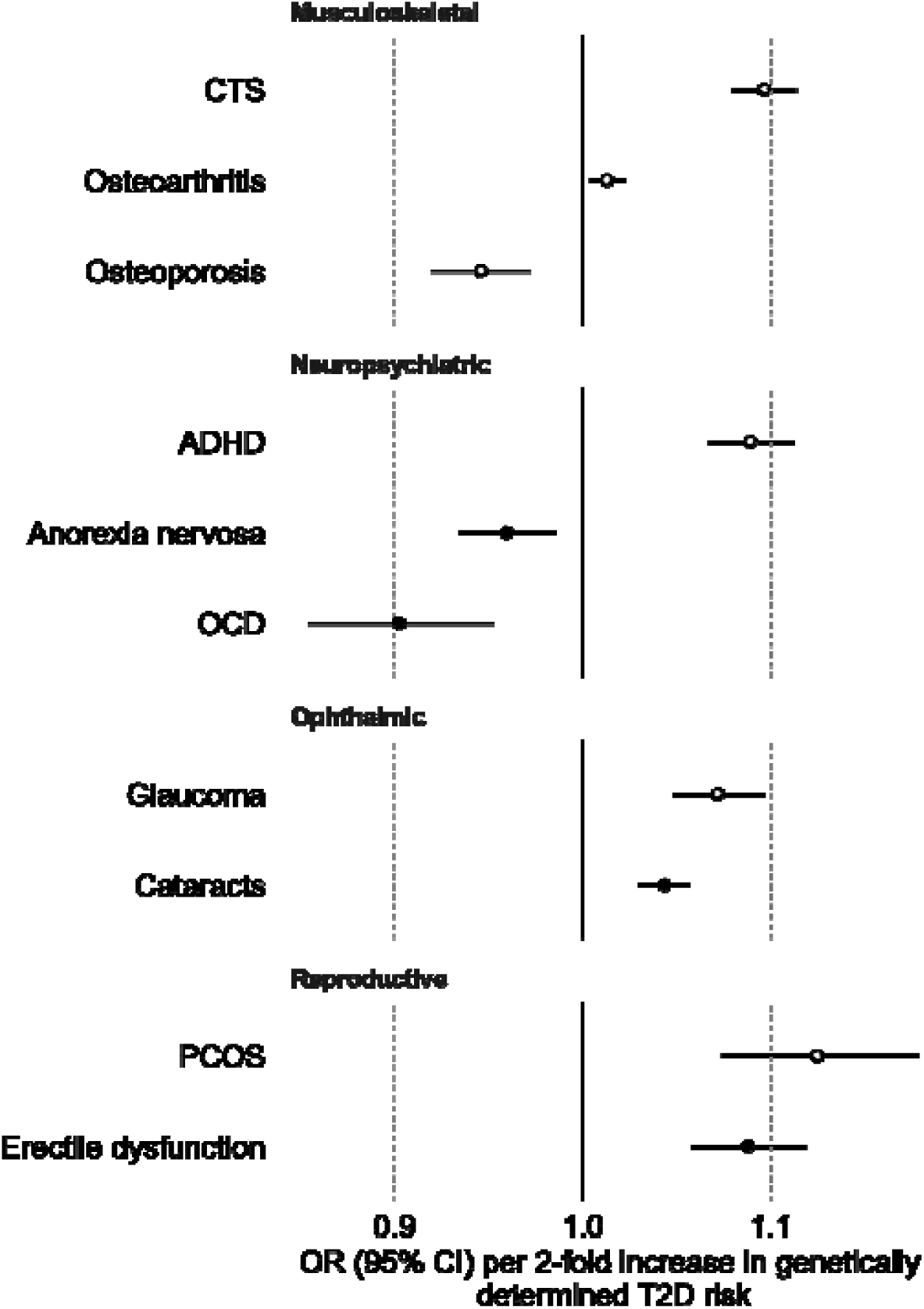
Results of two-sample Mendelian randomization (MR) analysis of genetic predisposition to type 2 diabetes (T2D) on non-cardiovascular comorbidity risk for the putative causal relationships (q-value < 0.05). Causal estimates are expressed as the odds ratio (OR) for each comorbidity per doubling (2-fold increase) in genetically determined dichotomous T2D risk. Filled circles mark estimates that passed the sensitivity analyses to assess the validity of the MR assumptions. (CI = confidence interval; CTS = Carpal tunnel syndrome; ADHD = attention-deficit/hyperactivity disorder; OCD = obsessive-compulsive disorder; PCOS = polycystic ovary syndrome)

### Distinct biological mechanisms contribute to the effect of T2D genetic predisposition on comorbidities

In addition to the effects of overall genetic predisposition to T2D on comorbidity risk, we investigated the effects of genetic burden for T2D mechanistic subtypes using the T2DGGI genetic clusters (Extended Figure 1). All identified putative causal relationships between overall genetic predisposition to T2D and its comorbidities, except for anorexia nervosa, are accompanied by at least one cluster-stratified effect, suggesting specific underlying biological pathways (Figure 3).

**Figure 3:**
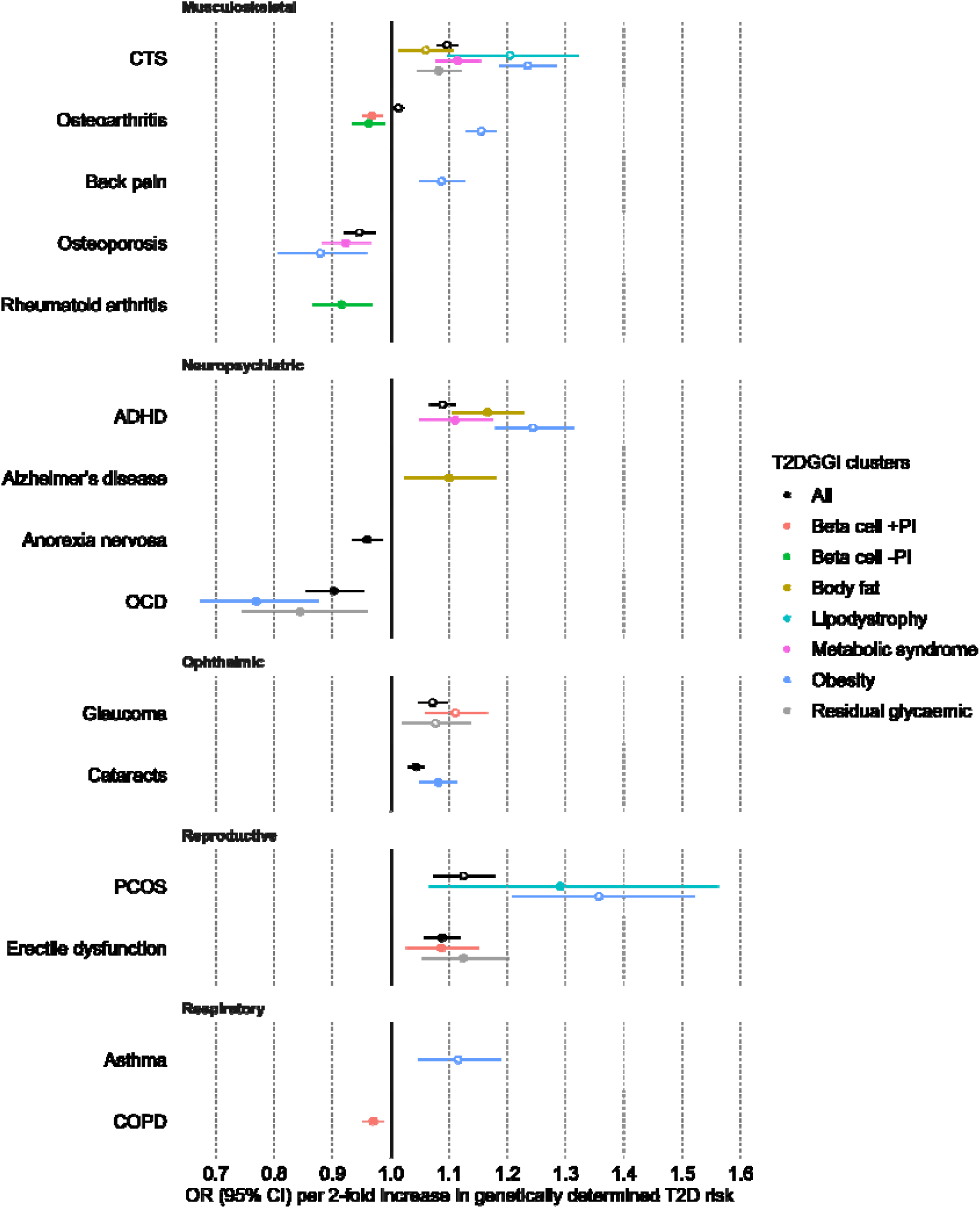
Results of cluster-stratified two-sample Mendelian randomization (MR) analysis of genetic predisposition to type 2 diabetes (T2D) on non-cardiovascular comorbidities risk for the putative causal relationships (q-value < 0.05). Causal estimates are expressed as the odds ratio (OR) for each comorbidity per doubling (2-fold increase) in genetically determined dichotomous T2D risk. Filled circles mark estimates that passed all sensitivity analyses to assess the validity of the MR assumptions. (T2DGGI = Type 2 Diabetes Global Genomics Initiative; CI = confidence interval; PI = proinsulin; CTS = Carpal tunnel syndrome; ADHD = attention-deficit/hyperactivity disorder; OCD = obsessive-compulsive disorder; PCOS = polycystic ovary syndrome; COPD = chronic obstructive pulmonary disease)

Within the power constraints of our study, we also provide evidence of putative causal effects of certain T2D genetic clusters without the presence of a causal effect of overall genetic predisposition to T2D on five further comorbidities: chronic back pain, rheumatoid arthritis, Alzheimer’s disease, asthma, and chronic obstructive pulmonary disease (COPD) (Figure 3, SupTab2). To investigate whether the observed effects are specific to the respective clusters, we performed leave-one-cluster-out MR analyses using all T2D genetic risk variants except those assigned to the putative causal cluster (Methods, Supplemental Note, SupTab4). In the leave-one-cluster-out MR analyses, we find no evidence of a causal effect of T2D genetic predisposition on any of the five diseases, indicating specific biological mechanisms, represented by the putative causal clusters, that may underlie these relationships.

### The impact of T2D obesity-related pathways on cataracts, asthma, and back pain

We find potential causal effects of the obesity cluster on 9 out of the 15 comorbidities (60%) causally affected by any genetic predisposition to T2D. The effects of the obesity cluster are consistently the largest across all clusters (Figure 3, SupTab2. For chronic back pain (OR=1.09, q-value=6.35×10^-5^), cataracts (OR=1.08, q-value=3.84×10^-6^), and asthma (OR=1.12, q-value=5.95×10^-3^), we find a risk-increasing effect of the obesity cluster but not of any of the other T2D genetic clusters. Concordantly, we observe a positive association between the obesity cluster genetic risk variants and all three diseases in the independent All of Us cohort dataset (back pain: OR=1.03, q-value=1.52×10^-5^; asthma: OR=1.04, q-value=7.9×10^-7^; cataracts: OR=1.08, q-value=2.1×10^-2^). Within the power bracket of our study, our findings indicate that obesity is the main biological mechanism driving the observed effect of T2D genetic predisposition on chronic back pain, cataracts, and asthma.

Body mass index (BMI) can act as a confounder of the identified relationships as it is a shared risk factor of T2D and the 9 comorbidities putatively causally affected by the obesity cluster (Supplemental Figure 2, Methods, SupTab5). When we adjust the obesity cluster estimates for BMI through multivariable MR, we no longer observe an effect of T2D genetic predisposition on any disease except cataracts, CTS, and osteoporosis (Supplemental Figure 28-33, SupTab6). Our sensitivity analyses show statistical evidence of a robust risk-increasing causal effect of T2D genetic predisposition on cataracts driven by the obesity genetic cluster.

### The impact of T2D glucose regulation mechanisms on glaucoma, COPD, rheumatoid arthritis, and erectile dysfunction

Beta cell-related clusters drive the putative protective effect of T2D genetic predisposition on rheumatoid arthritis (-PI: OR=0.92, q-value=1.35×10^-2^) and COPD (+PI: OR=0.97, q-value=7.42×10^-3^), adding genetic evidence to the potential role of beta cell dysfunction in these diseases[23–26]. In addition to the beta cell clusters, variants in the residual glycaemic cluster, which are most strongly associated with fasting glucose, also drive the risk-increasing effects of genetic predisposition to T2D on erectile dysfunction (residual glycaemic: OR=1.12, q-value=4.54×10^-3^; beta cell +PI: OR=1.09, q-value=3.06×10^-2^) and glaucoma (residual glycaemic: OR=1.08, q-value=4.78×10^-2^; beta cell +PI: OR=1.11, q-value=2.61×10^-4^). For erectile dysfunction, sensitivity analyses show statistical evidence of a robust risk-increasing causal effect of T2D genetic predisposition driven by glucose regulation clusters (Supplemental Note).

### The impact of T2D non-glucose-related pathways on ADHD, PCOS, Alzheimer’s disease, and osteoporosis

The protective effect of genetic predisposition to T2D on osteoporosis is driven by the metabolic syndrome (OR=0.89, q-value=3.62×10^-3^) and obesity clusters (OR=0.83, q-value=2.44×10^-2^). Similarly, we find negative associations in the All of Us cohort between T2D genetic risk variants assigned to the obesity cluster and osteoporosis (obesity: OR=0.94, q-value=3.99×10^-7^). For PCOS, the risk-increasing effect of genetic predisposition to T2D is driven by the obesity (OR=1.55, q-value=3.84×10^-6^) and lipodystrophy clusters (OR=1.44, q-value=4.93×10^-2^).

For Alzheimer’s disease, our findings indicate a risk-increasing effect of T2D genetic predisposition only through the body fat cluster (OR=1.10, q-value=4.83×10^2^). Adjusting for high-density lipoprotein (HDL) levels fully attenuates this effect (Supplemental Figure 47, SupTab6). For ADHD, a combination of the obesity (OR=1.24, q-value=7.57×10^-14^), body fat (OR=1.17, q-value=2.83×10^-7^), and metabolic syndrome clusters (OR=1.11, q-value=2.77×10^-3^) drive the risk-increasing effect of T2D genetic predisposition. These results suggest that non-glucose-related metabolic and adiposity pathways may contribute to the link between T2D genetic predisposition and increased ADHD and Alzheimer’s disease risks.

### The multifactorial impact of T2D on CTS, OCD and osteoarthritis

We find evidence of a putative causal link between all T2D genetic clusters except the beta cell clusters and CTS (Figure 3, SupTab2), indicating a multifactorial association mediated by diverse biological pathways. The strongest effects are observed for the obesity (OR=1.23, q-value=6.77×10^-24^) and lipodystrophy clusters (OR=1.20, q-value=1.15×10^-3^). In line with these findings, T2D genetic risk variants assigned to these two clusters are positively associated with CTS in the All of Us data (obesity: OR=1.07, q-value=5.02×10^-8^, lipodystrophy: OR=1.04, q-value=4.08×10^-2^) (SupTab7). The obesity (OR=0.77, q-value=1.15×10^-3^) and residual glycaemic clusters (OR=0.84, q-value=4.85×10^-2^) drive the protective effect of T2D genetic predisposition on OCD.

Osteoarthritis is the only disease for which we find divergent patterns of potentially causal effects across the T2D genetic clusters. The identified risk-increasing effect of genetic predisposition to T2D on osteoarthritis is driven by the obesity cluster (OR=1.15, q-value=3.55×10^-35^). Supporting our findings, the T2D risk variants assigned to the obesity cluster are positively associated with osteoarthritis risk in the All of Us cohort (OR=1.06, q-value=3.4×10^-13^). In contrast, genetic predisposition to T2D restricted to the beta cell clusters is associated with reduced osteoarthritis risk (+PI: OR=0.97, q-value=2.17×10^-3^, -PI: OR=0.96, q-value=4.43×10^-2^). All estimates of T2D genetic predisposition on osteoarthritis risk are attenuated after adjusting for the effects of BMI, waist-to-hip ratio (WHR), and subcutaneous adipose tissue (SAT) volume (Supplemental Figure 32,34,37, SupTab6).

### Potential causal effects of genetic predisposition to T2D vary across genetic ancestry groups

We examined the MR findings within each genetic ancestry group prior to meta-analysis. (Extended Figure 2). The groups comprise individuals genetically similar to Europeans (EUR), East Asians (EAS), South Asians (SAS), and Admixed Americans (AMR) as defined by the 1000 Genomes Project phase 3[21]. We observe divergent patterns of association between T2D genetic predisposition and depression, asthma, and COPD risk among different genetic ancestry groups (Figure 4, SupTab8). Although no statistical evidence of a causal effect of T2D genetic predisposition on depression was found in the meta-analysis across global populations, a protective effect driven by the obesity cluster was found in EAS. On the contrary, in EUR we find evidence of a risk-increasing effect of T2D genetic predisposition on depression linked to the obesity and body fat clusters (Figure 4, SupTab8). All T2D genetic risk variants and those assigned to the obesity cluster are positively associated with depression in the All of Us cohort (all: OR=1.03, q-value=5.35×10^-4^, obesity: OR=1.05, q-value=4.02×10^-4^).

**Figure 4:**
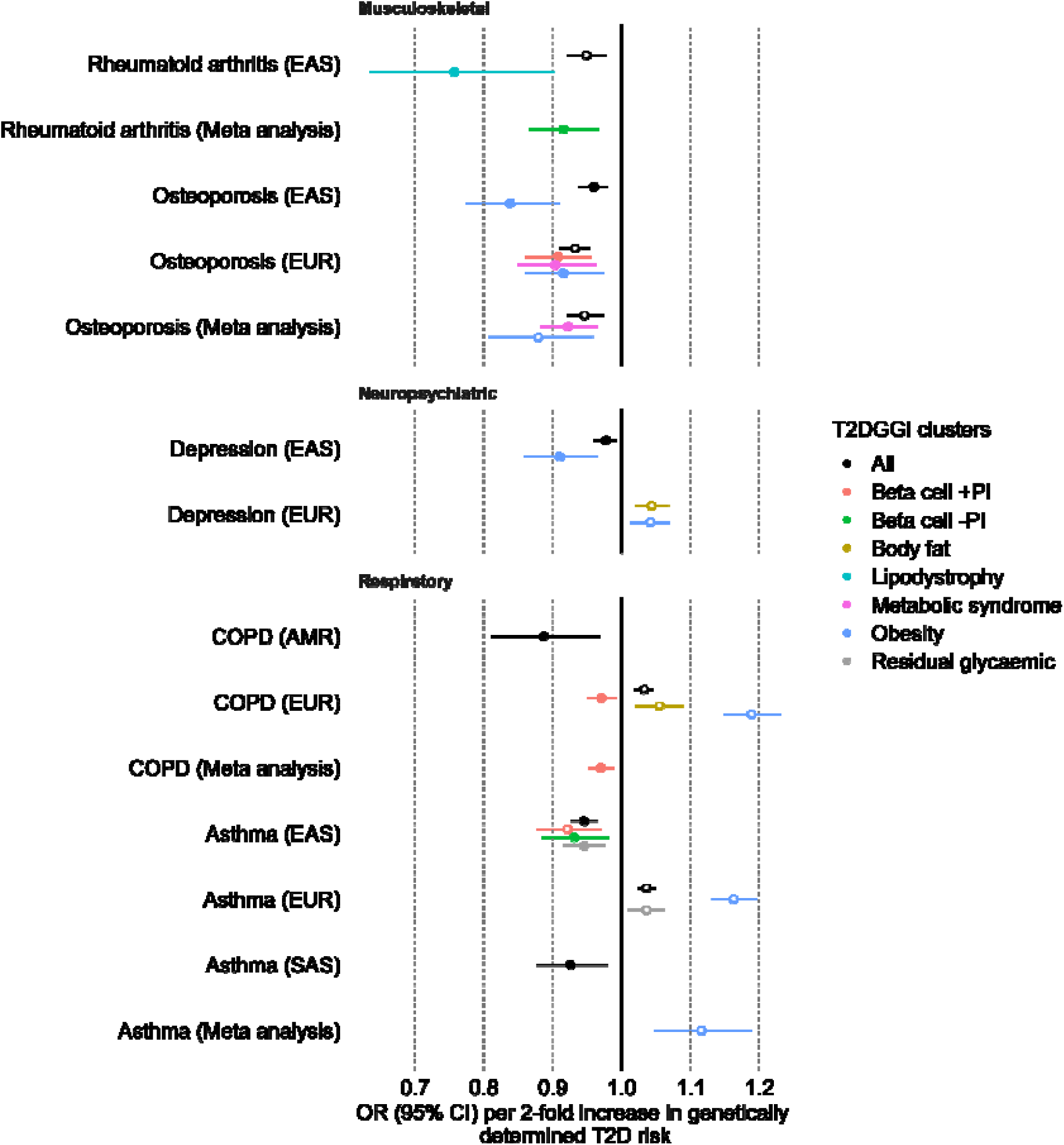
Mendelian randomization (MR) results for different global genetic ancestry groups using cluster-stratified genetic predisposition to T2D and risk for T2D non-cardiovascular comorbidities (EUR=individuals genetically similar to Europeans, EAS=individuals genetically similar to East Asians) for the putative causal relationships (q-value < 0.05). Causal estimates are expressed as the odds ratio (OR) of each comorbidity per doubling (2-fold increase) in genetically determined dichotomous T2D risk. Filled circles mark robust estimates that passed all sensitivity analyses (T2DGGI = Type 2 Diabetes Global Genomics Initiative; CI = confidence interval; PI = proinsulin).

We find a protective effect of T2D genetic predisposition on COPD in AMR, contrasting a risk-increasing effect in EUR. For EUR, we find additional risk-increasing effects from the body fat and obesity clusters and a protective effect from the beta cell +PI cluster (Figure 4, SupTab8). In addition to COPD, divergent effects across T2D genetic clusters are observed in osteoarthritis. For both diseases, the risk-increasing effects of the obesity cluster oppose the protective effects of beta cell-related clusters. Results in EAS and SAS show a putative protective effect of T2D genetic predisposition on asthma. In EAS, the effect is driven by the beta cell and the residual glycaemic clusters. Conversely, in EUR, we identify a risk-increasing effect driven by the obesity cluster. These opposing effect directions across different genetic ancestry groups may reflect differences in environmental factors, such as distinct effects of adiposity on T2D pathophysiology across global populations[27, 28]. A further explanation may be the different strengths of association between the T2D genetic clusters and the genetic ancestry groups. For example, in the latest T2DGGI work it was shown that variants assigned to the obesity cluster have greater allelic effects on T2D in EUR than EAS, whereas those assigned to the beta cell clusters have a greater allelic effect on T2D in EAS compared to EUR[11].

### Genetic evidence pointing to metformin as a drug repurposing opportunity for osteoarthritis

Drugs approved for the treatment of T2D may be relevant repurposing candidates for non-cardiovascular comorbidities. Given the availability of a curated list of high-confidence effector genes based on orthogonal functional genomics evidence for osteoarthritis[29], we have applied this hypothesis to the T2D-osteoarthritis comorbidity as proof of principle. By overlaying the osteoarthritis prioritized effector genes with genes targeted by T2D-approved drugs, we find that metformin, an antihyperglycemic drug, targets the osteoarthritis effector gene *NDUFS3* (GRCh38 chr11:47,565,336-47,584,562). In addition to its effect on lowering blood glucose levels, metformin also has anti-inflammatory and body weight-reducing effects[30]. The T2D index genetic risk variant closest to *NDUSF3*, rs2125838 (GRCh38 chr11:47,790,381), is assigned to the obesity cluster, for which we find a risk-increasing effect on osteoarthritis. In GTEx v10, rs2125838 is a *cis*-eQTL for *NDUFS3* in fibroblasts and cerebellar hemisphere. Previous studies, including Phase III clinical trials, have shown evidence of the beneficial effects of metformin treatment on a subset of osteoarthritis symptoms[31] and in delaying the onset of this disease in T2D patients[32]. Here we provide genetic evidence supporting the weight-reducing effect of metformin as a potential mechanism of action in alleviating the burden of osteoarthritis.

## Discussion

T2D is a leading global health concern that impacts individuals and healthcare systems. Effective prevention strategies for T2D and its comorbidities require a deeper understanding of the shared etiology underlying these relationships. Here, we sought to investigate the biological mechanisms underlying putative causal effects of cluster-stratified T2D genetic predisposition on 21 non-cardiovascular, non-oncogenic comorbidities. Our results show that T2D genetic predisposition is a potential driver of comorbidities, rather than the risk of T2D being causally affected by genetic predisposition to its comorbidities. Moreover, T2D genetic predisposition is primarily linked to risk-increasing effects on its comorbidities, reinforcing the role of genetic burden to T2D as a risk factor. The obesity cluster drives most of the observed associations and shows the strongest effects, supporting the well-known role of obesity as a common risk factor for multiple chronic diseases[33]. Finally, we identify evidence of potential causal effects of T2D genetic predisposition on asthma, depression, and COPD with opposing directions across global populations. This observed variation may be largely due to differences in environmental factors across populations [28].

Our genetic findings align with epidemiological observations for many of the identified associations, including cataracts[34], glaucoma[35, 36], PCOS[37, 38], and erectile dysfunction[39–41]. For instance, cohort studies have shown a decreased risk of T2D in anorexia nervosa patients[42], and a longitudinal observational study using the Polish National Health Fund data showed a decreasing trend of anorexia nervosa prevalence in T2D patients[43]. The risk of fractures, a proxy for osteoporosis, is lower in T2D patients than in healthy controls[44], in line with the identified protective effect of T2D genetic predisposition on osteoporosis. We validate previously described evidence of putative causal effects of T2D genetic predisposition on CTS[12], osteoporosis[13], glaucoma[14, 15], cataracts[16], PCOS[17], erectile dysfunction[18], and ADHD risk[19, 20] and prioritize the biological mechanisms underlying these associations. For instance, we show that glucose regulation mechanisms contribute to the associations between T2D genetic predisposition and erectile dysfunction and glaucoma, which aligns with previous genetics-based studies using glycaemic traits data[15, 18, 45, 46]. Obesity has been positively associated with cataracts in a meta-analysis of over 1.6 million individuals[47]. Concordantly, our findings highlight obesity as one of the mechanisms underlying the putative causal effect of T2D genetic predisposition on cataract risk. We also find evidence that genetic variants associated with increased HDL levels have a putative protective effect on clinically diagnosed Alzheimer’s disease risk, opposing the previously established risk-increasing effect[48, 49]. Possible explanations for the observed opposing effect directions include Alzheimer’s disease case definitions of previous reports relying on proxy cases and non-age-matched controls[50]. Further studies are needed to disentangle the relationship between HDL levels, T2D, and Alzheimer’s disease. For osteoarthritis, we find evidence of diverging cluster-stratified effects of genetic predisposition to T2D, validating the previously shown opposite association between both diseases in addition to shared risk-increasing obesity mechanisms[51].

In addition, our work highlights the T2D-approved drug metformin as a drug repurposing opportunity for osteoarthritis. Metformin is an inhibitor of the mitochondrial complex I, leading to reduced ATP production and activation of the AMPK pathway, a central regulator of cellular energy homeostasis[30]. It targets the osteoarthritis effector gene *NDUFS3* that encodes a core subunit of the mitochondrial complex 1, essential for its catalytic activity and assembly. Homozygous or compound heterozygous mutations in *NDUFS3* lead to mitochondrial complex 1 deficiency nuclear type 8 in humans[52]. Patients suffering from this condition show a wide range of phenotypes, including lactic acidosis and kyphoscoliosis. NDUFS3 is more highly expressed in degraded compared to intact osteoarthritis cartilage[53], pointing to a potential role of this protein in advancing cartilage degradation.

Cluster-stratified MR analysis has been used previously to identify specific biological mechanisms driving putatively causal relationships and potential pleiotropic pathways[54–56]. Although derived from biological associations, one limitation of the eight T2D mechanistic clusters is that they reflect cardiometabolic processes, excluding other potential biological mechanisms involved in T2D. Moreover, the T2D genetic risk variants were clustered based on a hard-clustering approach, which assigns all risk variants to exactly one cluster, with no overlap. The lack of outliers in this approach might decrease the robustness of each cluster. Given the broad nature of the cardiometabolic traits used to cluster the T2D genetic risk variants, they are posited to influence the risk for several diseases, including the comorbidities investigated here. We addressed the potential of confounding or mediation relationships by adjusting the MR estimates for the T2D-related cardiometabolic traits in a multivariable MR approach. If the adjusted effect of T2D genetic predisposition on comorbidity risk is attenuated to zero, care should be taken when interpreting the results given that the unadjusted estimates might be biased.

We recognize that our findings are constrained by the discovery power of the GWAS used. Larger sample sizes may reveal associations with additional T2D genetic clusters. Moreover, the sample size of the T2DGGI GWAS meta-analysis used to identify T2D genetic risk variants and create the mechanistic clusters is much larger than some of the non-cardiovascular comorbidity GWAS used in this work. This may influence the power to detect causal effects in the reverse direction MR analysis. We acknowledge that validation of the findings using sex-specific T2D GWAS data is needed for sex-specific diseases. A well-known limiting factor in statistical genetics analyses is the relative paucity of GWAS data from diverse global populations[57]. In our work, this bias might lead to T2D genetic IVs having a different effect in under-represented global populations and, hence, being less powerful IVs for these populations. Despite the limitations, we identify evidence of a potential causal effect of genetic predisposition to T2D on rheumatoid arthritis only when meta-analyzing the causal estimates across global populations, highlighting the discovery power gain of integrating diverse GWAS data. Our results underscore the need for the community to continue to pursue efforts to further increase diversity in genetic studies.

In conclusion, we provide evidence of potential causal links between different mechanistic subtypes of T2D genetic predisposition and its non-cardiovascular comorbidities. These findings can inform preventive strategies to mitigate the onset of long-term co-occurring conditions by stratifying and monitoring patients based on their genetic burden to T2D mechanistic subtypes. Moreover, we explore opportunities for drug repurposing, with T2D-approved drugs holding promise for targeting comorbidities. Our work paves the way for more stratified treatment approaches aligned with the genetic and multimorbidity profiles of patients.

## Methods

### Datasets

The T2DGGI consortium considered six genetic ancestry groups, which refer to the 1000 Genomes Project phase 3[21]: European, East Asian, African American, admixed American, South Asian, and South African (Table_T2DGGI_samplesize). In the meta-analysis across all global populations (2,535,601 individuals including 428,452 cases), 1,289 index (r^2^ < 0.05) T2D genetic risk variants were identified at genome-wide significance (p-value < 5×10^-8^) (SupTab9).

Previous efforts from the T2DGGI consortium have clustered the 1,289 T2D genetic risk variants based on their association profile with 37 cardiometabolic traits. The traits used for clustering included glycaemic traits, anthropometric measures, body fat and adipose tissue volume, blood pressure, circulating plasma lipids levels, and liver function and lipid metabolism biomarkers. A hard clustering approach was performed in an unsupervised manner. This resulted in eight non-overlapping mechanistic clusters of T2D risk variants that represent distinct biological pathways: obesity (n=233), beta-cell associated with positive proinsulin (n=91), beta-cell associated with negative proinsulin (n=89), lipodystrophy (n=45), liver and lipid metabolism (n=3), residual glycaemic (n=389), body fat (n=273) and metabolic syndrome (n=166). We used the T2DGGI GWAS summary statistics to perform MR analyses within genetic ancestry groups[11]. For all other diseases and quantitative traits employed in this work, an overview of the sample sizes and underlying populations is found in SupTable10 and SupTab1.

### Approaches to select instrumental variables for T2D

To best maintain the robustness of the T2D genetic risk clusters, we used all 1,289 T2D GWAS index risk variants identified in the T2DGGI global meta-analysis to derive the main results. Index variants were defined as genome-wide significant (p-value < 5×10^-8^) variants with r^2^ < 0.05 over a 5Mb window[11]. If any genetic instrumental variable (IV) was not present in the outcome trait, we replaced it with an LD-based proxy (r^2^ >0.8) if available using the *LDlinkR::Ldproxy()* R function (v1.3.0)[58].

Using all T2D index risk variants, our definition of independent IVs is not as strict as the one commonly employed in MR analyses, defined as LD-based clumped genome-wide significant (p-value < 5×10^-8^) variants with r^2^ < 0.001 over a 10Mb window. To address this, we performed sensitivity analyses to select IVs for T2D (Supplemental Figures 3-27). Firstly, we have removed all IVs with evidence of weak instrumental bias, defined as an F-statistic = (*beta^2^*/*se^2^*) < 10, where *beta* is the effect size estimate, and *se* is its standard error from the T2D GWAS. Secondly, we compared the effect magnitude of our results with alternative approaches to select T2D IVs. We have employed three additional approaches to define T2D IVs: selecting one variant per locus, LD-based clumping all the T2D index risk variants, and performing cluster-wise LD-based clumping (SupTab11, SupTab12).

### Selection of instrumental variables for other traits

For all other traits employed here (non-cardiovascular comorbidities and cardiometabolic traits), we defined IVs as LD-based clumped genome-wide significant (p-value < 5×10^-8^) variants with r^2^ < 0.001 over a 10Mb window. Clumping was performed with *plink* (v1.9), and LD was calculated based on the 1000 Genomes Project phase 3 release[21] with matching genetic ancestry groups. If any IV was absent in the T2D matching genetic ancestry group GWAS, we replaced it with a proxy. We used the output of plink, which assigns all variants to a clumped result, to search for proxies using the *ieugwasr::ld_matrix()* R function (v1.0.2). We removed variants with an F-statistic<10.

### Two-sample Mendelian randomization analysis

Following the STROBE-MR guidelines[59], we performed bi-directional two-sample MR analyses[60] between genetic predisposition to T2D and 21 non-cardiovascular comorbidities. We used the *TwoSampleMR* R package (v0.5.7), which is curated by MR-Base[61]. It has been shown that the estimates of two-sample MR remain unbiased in the presence of sample overlap between exposure and outcome when using large sample sizes[62]. All analyses were performed within genetic ancestry groups using the LD panel from the corresponding genetic ancestry from the 1000 Genomes Project phase 3 release[21] to conduct clumping when necessary. Using matching genetic ancestry group data between exposure and outcome increases the accuracy of the causal estimate by reducing bias due to heterogeneous underlying data distributions.

For the main results, we applied the inverse variance weighted (IVW) method, which performs a random-effects meta-analysis of the Wald ratio estimate of each IV. If only one IV was available, the Wald ratio estimate was used. We applied additional MR methods to ensure consistency of causal effect direction under different methodological assumptions, namely the weighted median, MR-Egger[63], MR-PRESSO[64], and Steiger-filtered IVW methods [65]. We ran the MR-PRESSO distortion test with 1000 iterations to compute the null distribution for the outlier test and increased it to 1500 if it failed. If the distortion test was significant (p-value < 0.05), we considered the effect estimate of the MR-PRESSO outlier-corrected method. Otherwise, we used the estimate of the raw MR-PRESSO result. To facilitate interpretability when using binary traits as exposure, we report the MR results as OR for the outcome per doubling (2-fold change) in genetically determined dichotomous exposure risk. This unit is calculated by multiplying the MR effect sizes by ln(2) and converting them to OR via exponentiation[66].

### Assessment of the Mendelian randomization assumptions

MR relies on three main assumptions: relevance, independence, and the exclusion restriction criteria (Supplemental Note). The relevance assumption states that the IVs need to be strongly associated with the exposure and can be directly tested by selecting genome-wide significant and strong (F-statistic > 10) IVs. The independence assumption ensures that the IVs are not associated with any confounder of the exposure-outcome association. The exclusion restriction criterion assumes no horizontal pleiotropy by ensuring that the IVs affect the outcome only through the exposure and not through any alternative pathways. Both assumptions cannot be directly tested but can be assessed via sensitivity analyses. We have conducted multivariable MR analyses to address the independence assumption by adjusting the univariable MR effect for known biological confounders or mediators of the exposure and outcome relationship. To test for horizontal pleiotropy, we applied the MR-Egger intercept test (*TwoSampleMR::mr_pleiotropy_test()*) and the MR-PRESSO outlier and distortion test. Both methods provide causal estimates corrected for the detected pleiotropy, and we looked for a concordant effect direction across different MR methods and the IVW estimate. Moreover, we assessed heterogeneity using I^2^, a measure based on Cochran’s Q-statistic (*TwoSampleMR::mr_heterogeneity()*) that is more interpretable and independent of the number of studies[67]. Evidence for heterogeneity, defined as I^2^ > 50, implies that some IVs may influence the outcome through pathways other than the exposure, possibly violating the exclusion restriction assumption. Finally, we have tested for reverse causation by applying a bi-directional MR framework and a directionality test based on the Steiger filter (*TwoSampleMR::directionality_test()*) (SupTab13). Finally, we compared our results with the MR-Clust method, which groups together IVs with similar causal effect estimates on the outcome trait into distinct cluster[55] (Supplemental Note, SupTab14).

### Cluster-stratified two-sample Mendelian randomization analysis

To infer the putative causal effects of the eight mechanistic clusters of T2D genetic risk on the analyzed non-cardiovascular comorbidities, we conducted additional MR analyses restricting the T2D IVs to risk variants assigned to each cluster. For these analyses, we followed the same sensitivity analysis procedures described above. In addition, we used the correlated IVW method implemented in the *MendelianRandomization* R package (v0.10)[68], which allows for correlated IVs. This method is designed for less than 500 IVs and, therefore, could not be employed in the MR analyses using all 1,289 T2D genetic risk variants. As a sensitivity analysis to assess the specificity of the clusters, we performed a leave-one-out cluster MR analysis using as IVs the T2D genetic risk variants excluding all variants assigned to one cluster at a time.

### Meta-analysis across genetic ancestry groups

In cases of multiple publicly available GWAS summary statistics for a comorbidity from different genetic ancestry groups, we conducted a meta-analysis of the MR estimates across these groups. We used the *rma.uni()* function of the *metafor* R package (v4.6)[69] to conduct a random-effects meta-analysis using the IVW estimate. We tested for heterogeneity using a restricted maximum likelihood estimator (REML). We considered evidence for heterogeneity if I^2^ > 50.

### Definition of significance

We account for multiple testing burden for both tested directions (T2D genetic predisposition on comorbidity risk and genetic predisposition to comorbidity on T2D risk) together by correcting the p-values of the IVW estimate and, if applicable, the correlated IVW estimate across all analyses (including the meta-analyses) using the FDR method (referred to as q-values). We defined statistical significance as IVW estimates and, if applicable, correlated IVW estimates with a q-value<0.05.

### Multivariable Mendelian randomization analysis with cardiometabolic traits

For the cluster-stratified genetic predisposition to T2D MR results with FDR<5%, we subsequently performed multivariable MR (MVMR) analyses using the mechanistic clusters of T2D genetic predisposition and several cardiometabolic traits as exposure and the non-cardiovascular comorbidities as outcome. By adjusting cluster-stratified MR estimates for cardiometabolic traits causally associated with the comorbidities, we can estimate the direct effect of the mechanistic clusters of T2D genetic predisposition on its comorbidities. To compare the univariable and multivariable MR results, we first performed univariable MR analyses using the cardiometabolic traits as exposures and the investigated T2D comorbidities as outcomes (Supplemental Figure 2). The results were adjusted for multiple testing using FDR correction. IVs were selected using the same approach as for the comorbidities. We then conducted an MVMR analysis only for the cardiometabolic traits that showed a potential causal effect on comorbidity at FDR of 5%. We applied the *TwoSampleMR::mv_multiple()* function, which uses the IVW method to perform the MVMR analysis. We performed sensitivity analyses using the *MVMR* R package (v0.4)[70], including testing for heterogeneity (*qhet_mvmr()*), pleiotropy (*pleiotropy_mvmr()*), and instrumental strength of each exposure (*strength_mvmr()*).

### Phenome-wide association study of cluster-stratified T2D risk variants

To identify non-cardiovascular phenotypes associated with the T2DGGI clusters, we performed PheWAS in each cluster of genetic predisposition to T2D using the National Institutes of Health (NIH) All of Us cohort[22], which was not part of the T2DGGI meta-analysis. All research was carried out on the All of Us Researcher Workbench. Quality control (QC) and genetic ancestry group assignment were performed on 206,000 participants with EHR and WGS data. We pruned the dataset to a maximal set of unrelated individuals using a kinship coefficient < 0.1 and removed samples with ambiguous sex (SupTab15). To reduce the computational complexity, we used the All of Us ACAF variant set, which retains those variants with an allele frequency (AF) > 1% or allele count (AC) >100 in any computed genetic ancestry group.

For each genetic ancestry group, we calculated the first 10 principal components (PCs) using smartpca (v7.2.1)[71] with LD-pruned genotypes and the *fastmode* option enabled. To derive the LD-pruned genotypes, we restricted variants to autosomal variants that were present in 1000 Genomes Project phase 3 release[21], not in the major histocompatibility complex (MHC) region, minor allele frequency (MAF) ≥ 1%, and Hardy-Weinberg equilibrium (HWE) p-value ≥ 1×10-6. LD pruning was conducted using plink2[72] with r^2^= 0.05 in a 1000 KB window with an 80b step size. For each genetic ancestry group and T2D genetic cluster, we calculated a GRS summing the cluster-stratified risk alleles, weighted by the effect sizes from each genetic ancestry group. The GRS was adjusted for the 10 first PCs and scaled within each genetic ancestry group.

For each genetic ancestry group and T2D genetic cluster, we performed a PheWAS using the PheTK package (v0.2.1rc5)[73]. Phenotype code (phecode) counts were calculated using the count_phecode() function of PheTK, with the following options: ‘phecode_version=“X”’ and ‘icd_version=“US”’. Participants without any recorded phecodes were excluded to avoid incomplete EHRs. PheWAS were conducted with the PheWAS function of PheTK using logistic regression and the model ‘phecode ∼ sex + age at last EHR + PCs 1-10 + cluster GRS’. Only phecodes with a minimum case count of 50 were included, and cases were defined as having at least two instances of the respective phecode. We considered phecodes from the following categories: mental, musculoskeletal, neurological, respiratory, and sense organs. Multiple testing correction was applied for each genetic ancestry group separately using a q-value < 0.05. We meta-analyzed the cluster-stratified PheWAS across genetic ancestry groups using the random-effects IVW method and the *rma.uni()* function of the metafor R package (v4.6)[69]. The meta-analysis results were FDR corrected for multiple testing separately.

### Drug repurposing opportunity exemplified by osteoarthritis

By querying the OpenTargets database[74], we identified genes targeted by T2D-approved drugs. Among these genes, we searched for prioritized effector genes for osteoarthritis[29]. The prioritization of osteoarthritis effector genes was based on functional genomics evidence. To gather additional information on the drugs representing repurposing opportunities, we queried the DrugsBank database[75].

## Author contributions

A.L.A, O.B, H.T and D.C. contributed equally to this project. A.L.A, O.B, B.F.V., A.P.M. and E.Z. conceived the project and analysis plan. All authors reviewed the analysis plan. A.L.A, O.B, H.T and D.C. conducted all the analyses. The interpretation of the results was conducted by all authors. A.L.A and O.B. wrote the manuscript, which was reviewed by all authors.

## Acknowledgments

O.B has received funding from the European Union’s Horizon 2020 research and innovation program under Grant Agreement No 101017802 (OPTOMICS). Supported in part by the National Center for Advancing Translational Sciences, CTSI grant UL1TR001881, and the National Institute of Diabetes and Digestive and Kidney Disease Diabetes Research Center (DRC) grant DK063491 to the Southern California Diabetes Endocrinology Research Center. Infrastructure for the CHARGE Consortium is supported in part by the National Heart, Lung, and Blood Institute (NHLBI) grant R01HL105756. J.M.M. is supported by American Diabetes Association grant #11-22-ICTSPM-16 and by NHGRI U01HG011723, by the National Institute Of Diabetes And Digestive And Kidney Diseases of the National Institutes of Health under Award Number R01DK137993 and U01 DK140757, AMP CMD award from RFP 6 from the Foundation for the National Institutes of Health, and a Medical University of Bialystok (MUB) grant from the Ministry of Science and Higher Education (Poland).

## Data availability

The GWAS summary statistics used in this work are publicly available and referenced in Supplementary Tables 1 and 10. Researchers can apply to access the individual-level data of the All of Us program (https://researchallofus.org/) used to perform the PheWAS. The list of prioritized effector genes for osteoarthritis is publicly available in the referenced publication. The GTEx v10 data used in this manuscript were obtained from: the GTEx Portal on 02/21/25 (dbGaP Accession phs000424.v10.p2).

## Code availability

The code used to perform all the MR-related analysis can be found on GitHub under GNU General Public License (GPL-3) at https://github.com/nalu357/T2DGGI_comorbidity and on Zenodo at https://doi.org/10.5281/zenodo.15168490. The PheTK package used to perform the All Of Us PheWAS is freely available on the Python Package Index, on GitHub under GNU General Public License (GPL-3) at https://github.com/nhgritctran/PheTK and on Zenodo at https://doi.org/10.5281/zenodo.14217954.

## Extended figures

**Extended Figure 1:**
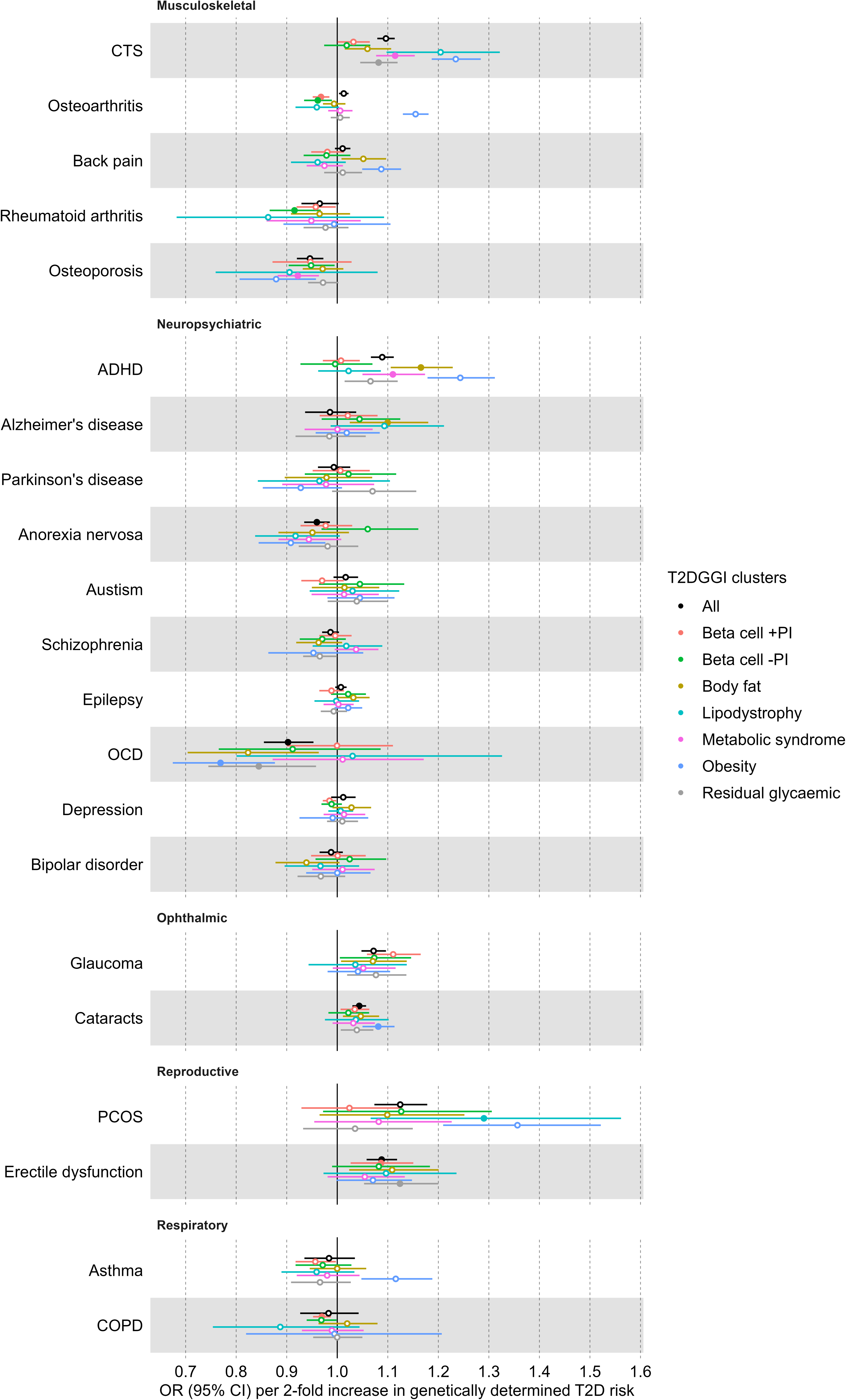
Results of cluster-stratified two-sample Mendelian randomization (MR) analysis of genetic predisposition to type 2 diabetes (T2D) on non-cardiovascular comorbidities risk. Causal estimates are expressed as the odds ratio (OR) of each comorbidity per doubling (2-fold increase) in genetically determined dichotomous T2D risk. Filled circles mark estimates with a q-value < 0.05 that passed all sensitivity analyses to assess the validity of the MR assumptions. (T2DGGI = Type 2 Diabetes Global Genomics Initiative; CI = confidence interval; PI = proinsulin; CTS = Carpal tunnel syndrome; ADHD = attention-deficit/hyperactivity disorder; OCD = obsessive-compulsive disorder; PCOS = polycystic ovary syndrome; COPD = chronic obstructive pulmonary disease)

**Extended Figure 2:**
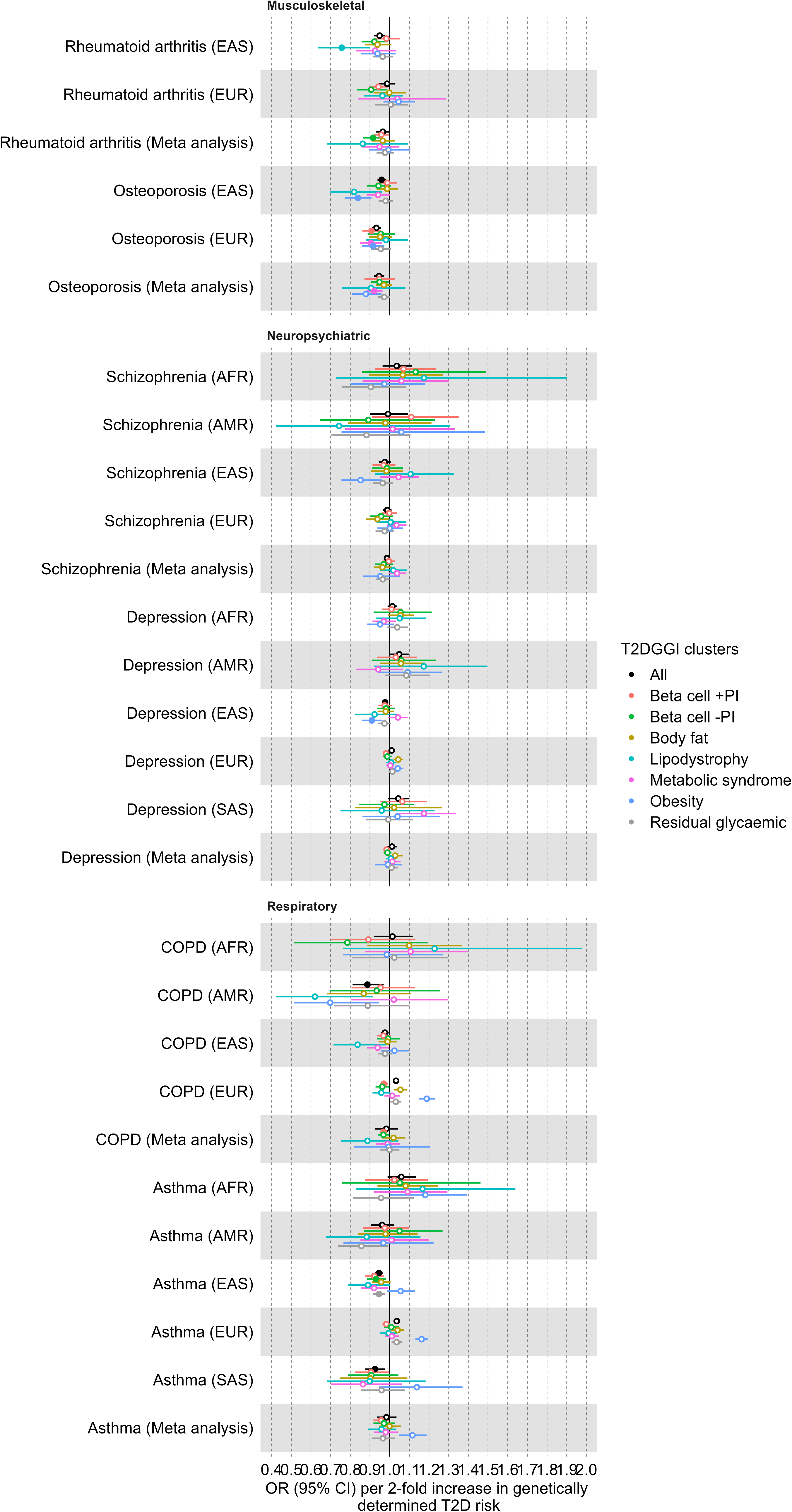
Results of the single-ancestry cluster-stratified two-sample Mendelian randomization (MR) analysis of genetic predisposition to type 2 diabetes (T2D) on non-cardiovascular comorbidities risk. Causal estimates are expressed as the odds ratio (OR) of each comorbidity per doubling (2-fold increase) in genetically determined dichotomous T2D risk. Filled circles mark estimates with a q-value < 0.05 that passed all sensitivity analyses to assess the validity of the MR assumptions. (T2DGGI = Type 2 Diabetes Global Genomics Initiative; CI = confidence interval; PI = proinsulin; COPD = chronic obstructive pulmonary disease)

