## Supplemental Figures for "The effect of type 2 diabetes genetic predisposition on non-cardiovascular comorbidities"

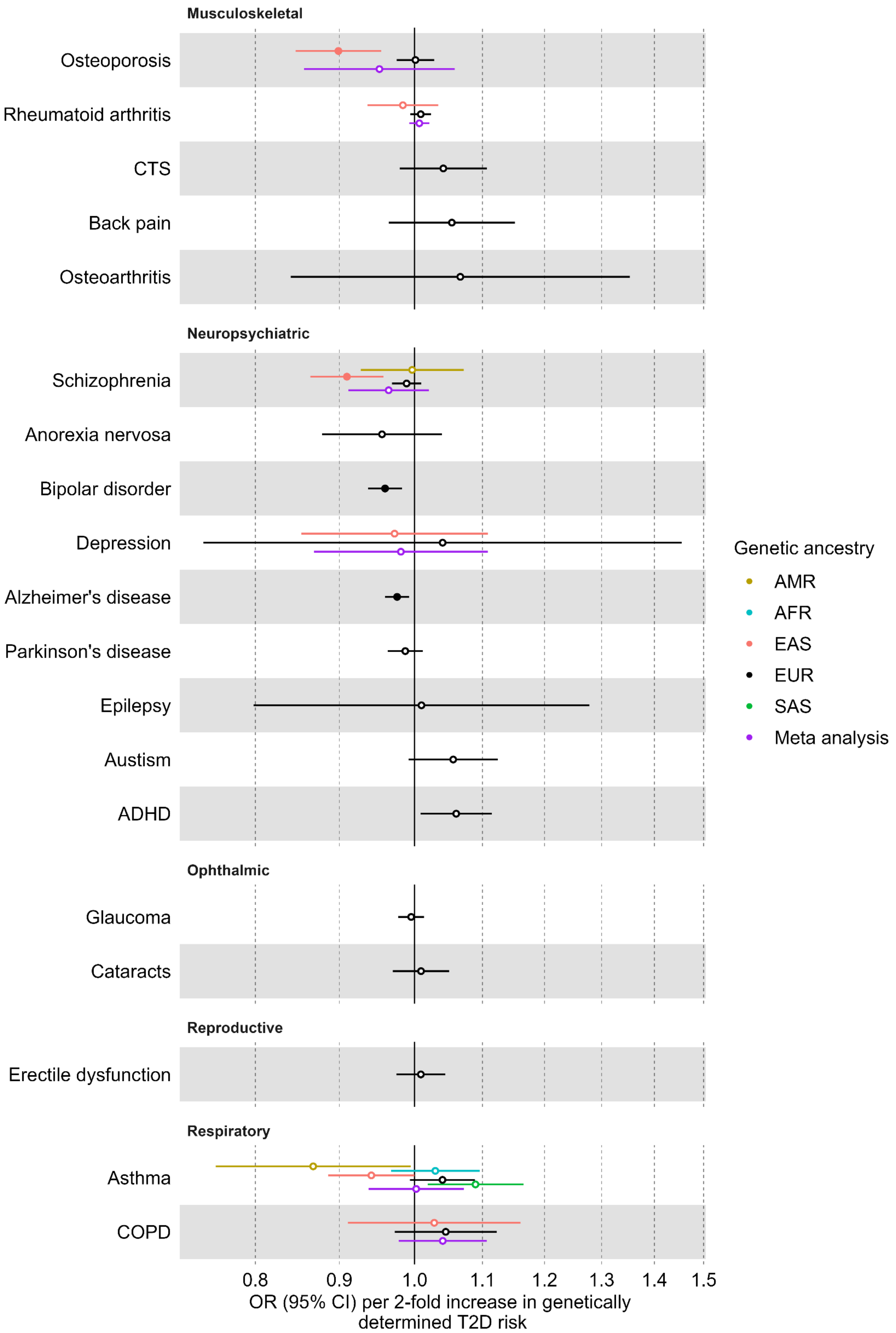


**Supplemental Figure 1**: Results of the reverse Mendelian randomization (MR) analysis using genetic predisposition to type 2 diabetes (T2D) comorbidities as exposure and T2D as the outcome. Causal estimates are expressed as the odds ratio of comorbidity risk per doubling (2-fold increase) in genetically determined dichotomous T2D risk. Filled circles mark estimates with a false-discovery-rate correction of 5%. No estimate passed the MR sensitivity analyses. The genetic ancestry groups represent individuals genetically similar to Africans (AFR), East Asians (EAS), Europeans (EUR), admixed Americans (AMR) and South Asians (SAS) as defined by the 1000 Genomes Project phase 3. (CI = confidence interval).

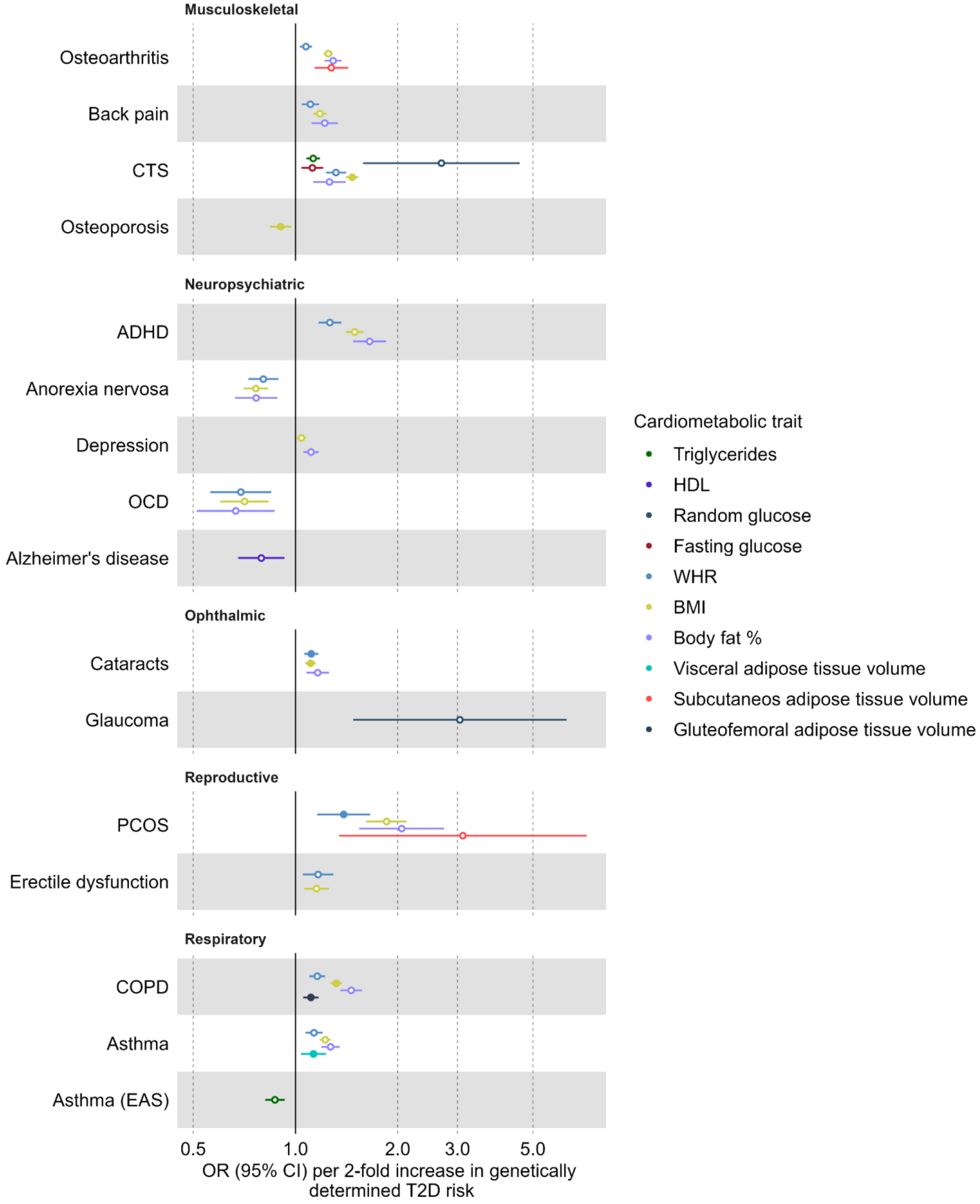


**Supplemental Figure 2:** Univariable Mendelian randomization (MR) results with genetic predisposition to cardiometabolic traits as exposures and type 2 diabetes (T2D) comorbidities as outcomes. Causal estimates are expressed as the odds ratio of comorbidity risk per doubling (2-fold increase) in genetically determined dichotomous T2D risk. Filled circles denote robust causal estimates that passed sensitivity analysis and false-discovery rate correction at 5%. (CI = confidence interval; PI = proinsulin; CTS = Carpal tunnel syndrome; ADHD = attention-deficit/hyperactivity disorder; OCD = obsessive-compulsive disorder; PCOS = polycystic ovary syndrome; COPD = chronic obstructive pulmonary disease; HDL=high-density lipoprotein cholesterol; WHR = waist-to-hip ratio; BMI=body mass index).

**Supplemental Figures 3-27:** Comparison between the results of Mendelian randomization (MR) results using the inverse variance weighted method and different approaches to select genetic instrumental variables (IVs). The forest plots depict all cluster-stratified estimates of genetic predisposition to type 2 diabetes (T2D) on comorbidity risk. Causal estimates are expressed as the odds ratio of comorbidity risk per doubling (2-fold increase) in genetically determined dichotomous T2D risk. Filled circles mark estimates with FDR < 5%. The genetic ancestry groups represent individuals genetically similar to Africans (AFR), East Asians (EAS), Europeans (EUR), admixed Americans (AMR) and South Asians (SAS) as defined by the 1000 Genomes Project. (CI = confidence interval).

3

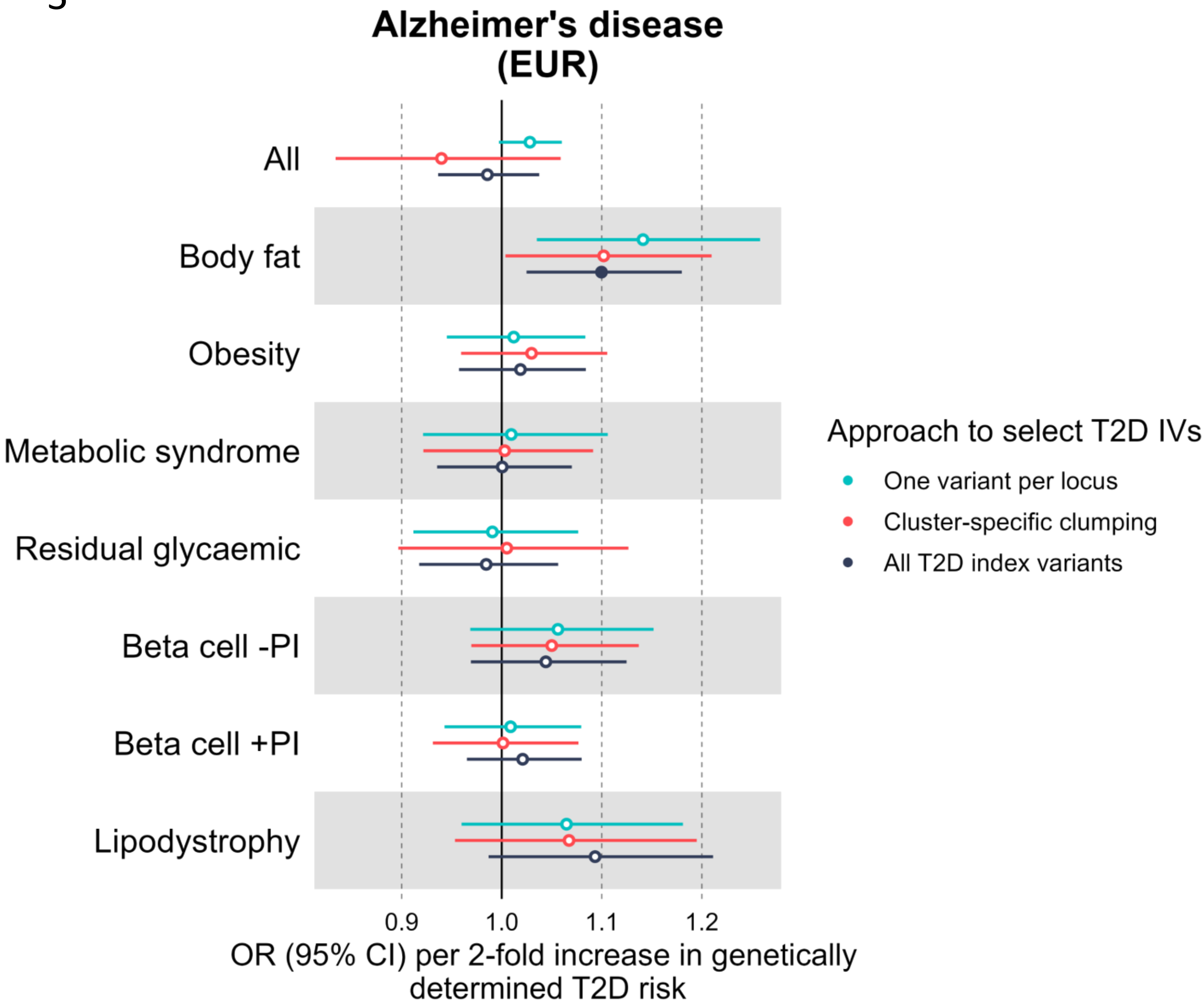

Alzheimer's disease (EUR)
All
Body fat
Obesity
Metabolic syndrome
Residual glycaemic
Beta cell -PI
Beta cell +PI
Lipodystrophy
0.9
1.0
1.1
1.2
OR (95% CI) per 2-fold increase in genetically determined T2D risk
Approach to select T2D IVs
One variant per locus
Cluster-specific clumping
All T2D index variants

4

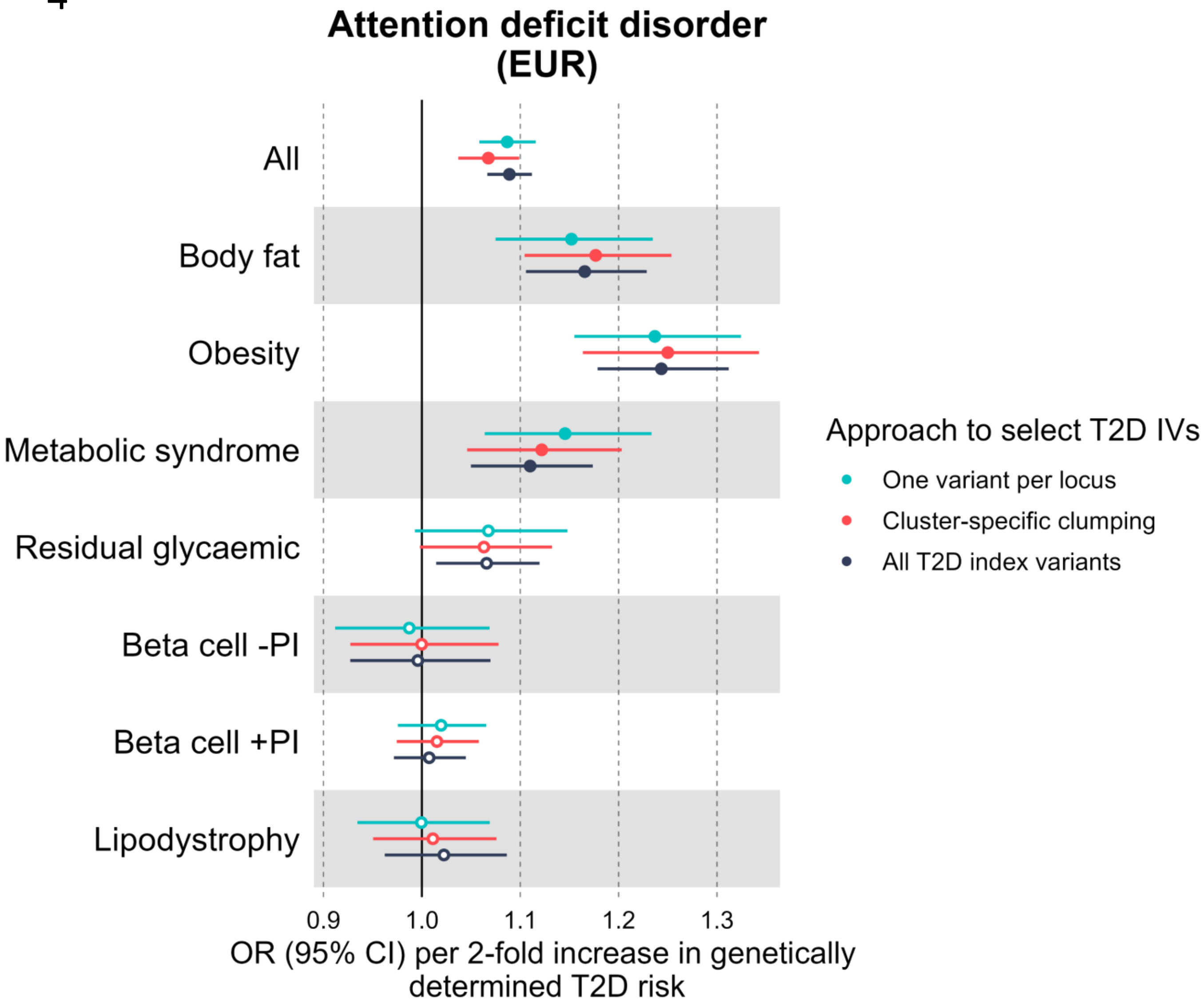
Attention deficit disorder (EUR)
All
Body fat
Obesity
Metabolic syndrome
Residual glycaemic
Beta cell -PI
Beta cell +PI
Lipodystrophy
0.9
1.0
1.1
1.2
1.3
OR (95% CI) per 2-fold increase in genetically determined T2D risk
Approach to select T2D IVs
One variant per locus
Cluster-specific clumping
All T2D index variants

5

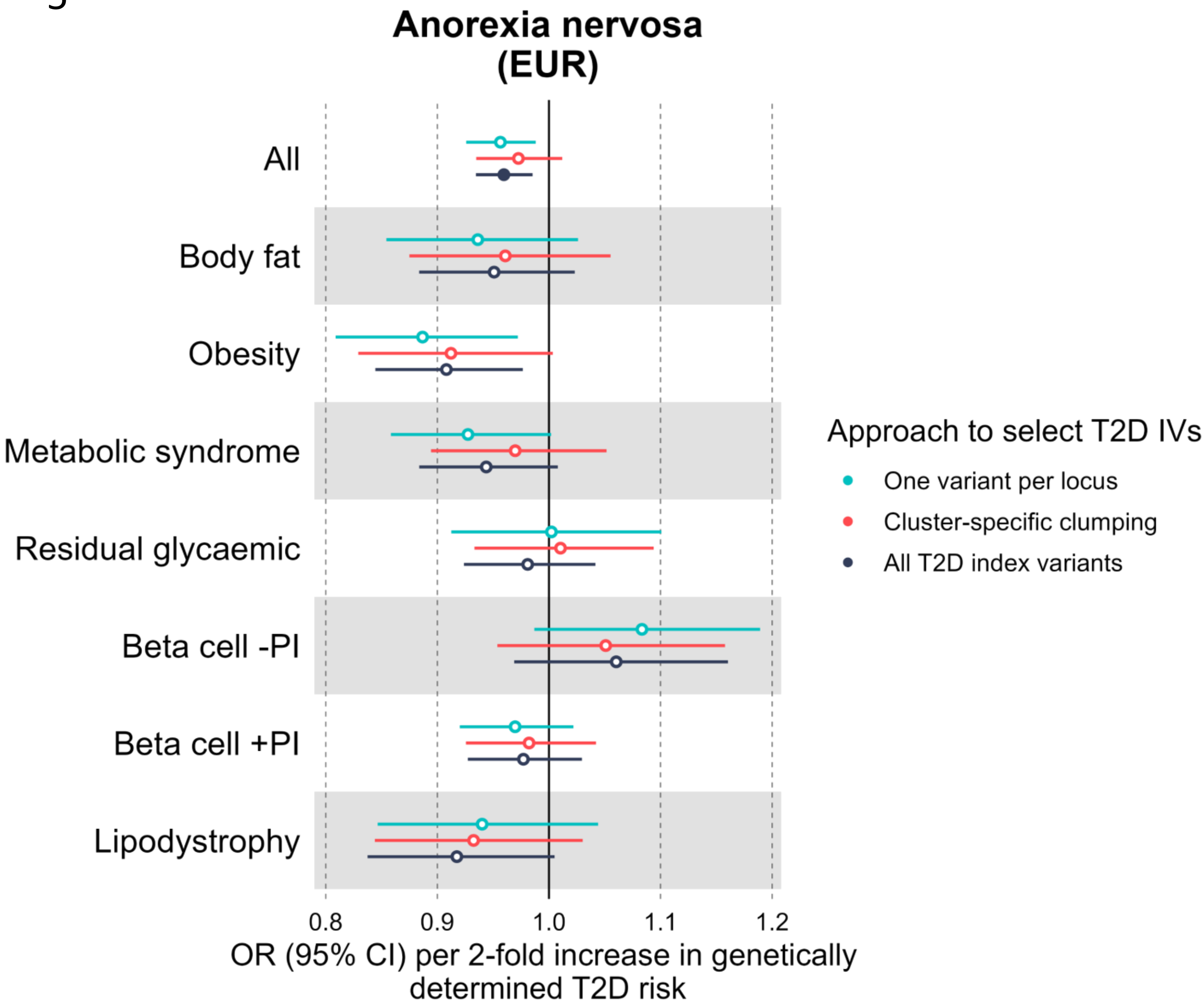

Anorexia nervosa
(EUR)
All
Body fat
Obesity
Metabolic syndrome
Residual glycaemic
Beta cell -PI
Beta cell +PI
Lipodystrophy
0.8
0.9
1.0
1.1
1.2
OR (95% CI) per 2-fold increase in genetically determined T2D risk
Approach to select T2D IVs
One variant per locus
Cluster-specific clumping
All T2D index variants

6

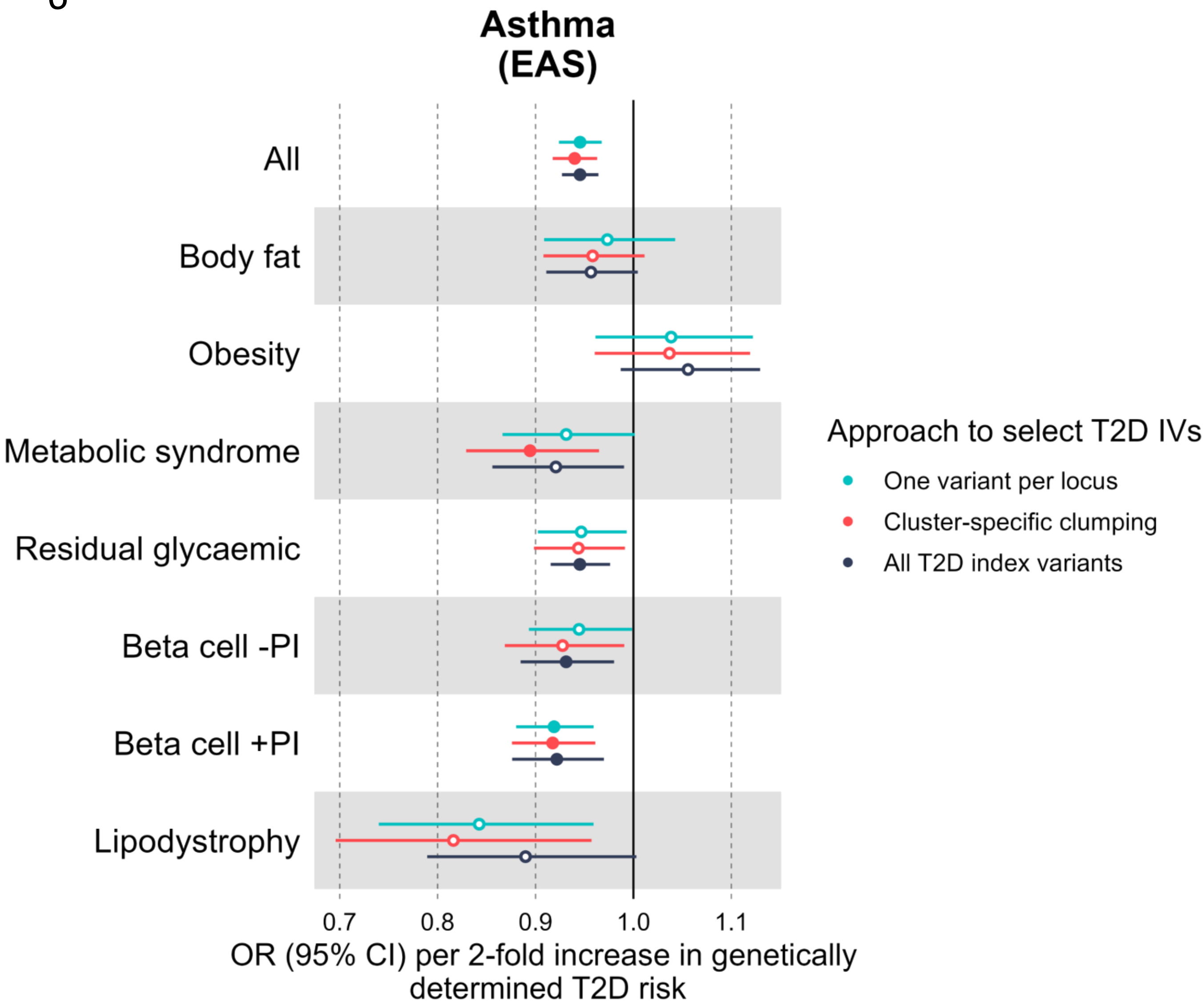

Asthma
(EAS)
All
Body fat
Obesity
Metabolic syndrome
Residual glycaemic
Beta cell -PI
Beta cell +PI
Lipodystrophy
0.7
0.8
0.9
1.0
1.1
OR (95% CI) per 2-fold increase in genetically determined T2D risk
Approach to select T2D IVs
One variant per locus
Cluster-specific clumping
All T2D index variants

7

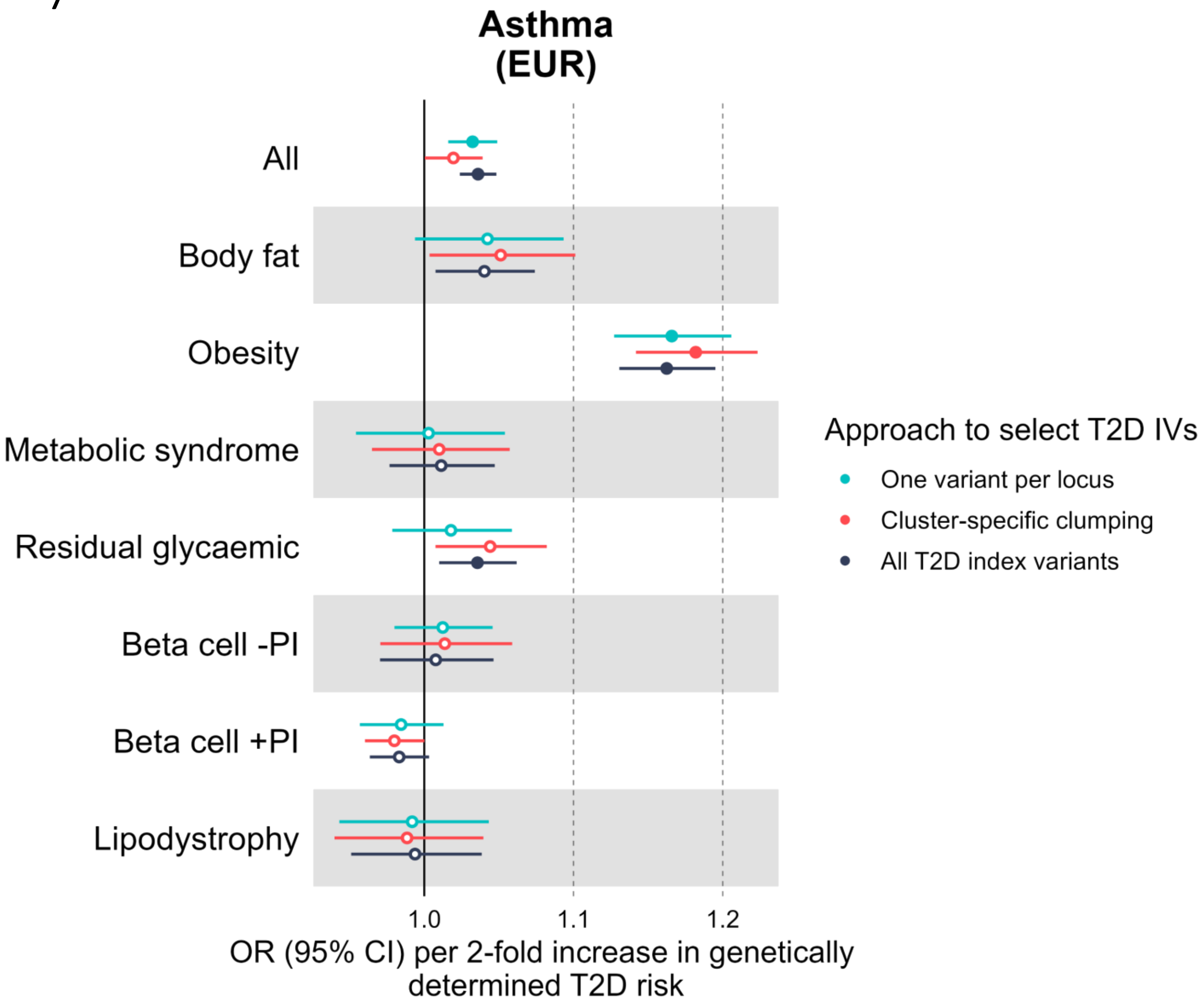
Asthma
(EUR)
All
Body fat
Obesity
Metabolic syndrome
Residual glycaemic
Beta cell -PI
Beta cell +PI
Lipodystrophy
1.0
1.1
1.2
OR (95% CI) per 2-fold increase in genetically determined T2D risk
Approach to select T2D IVs
One variant per locus
Cluster-specific clumping
All T2D index variants

8

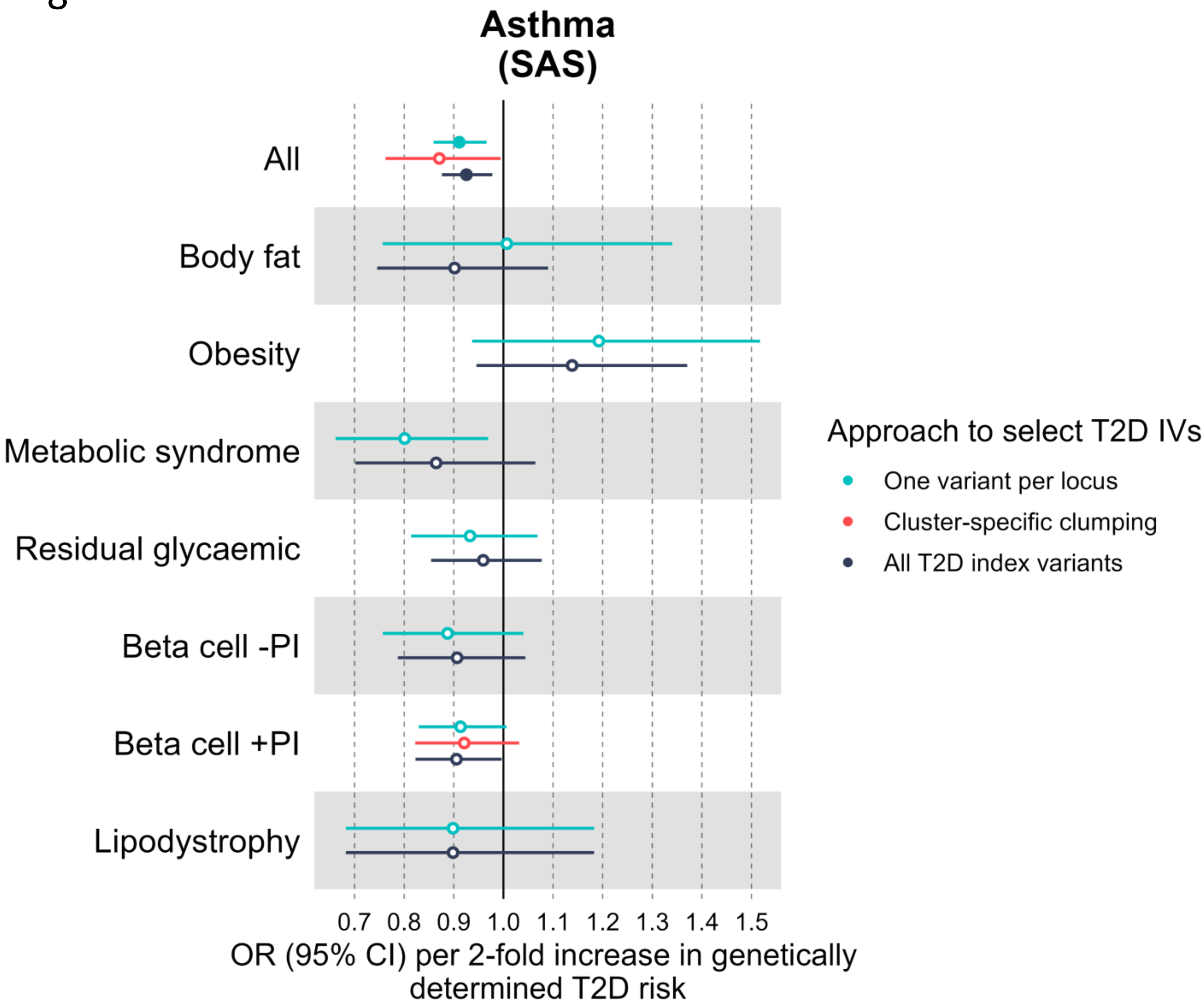

Asthma
(SAS)
All
Body fat
Obesity
Metabolic syndrome
Residual glycaemic
Beta cell -PI
Beta cell +PI
Lipodystrophy
0.7 0.8 0.9 1.0 1.1 1.2 1.3 1.4 1.5
OR (95% CI) per 2-fold increase in genetically determined T2D risk
Approach to select T2D IVs
One variant per locus
Cluster-specific clumping
All T2D index variants

9

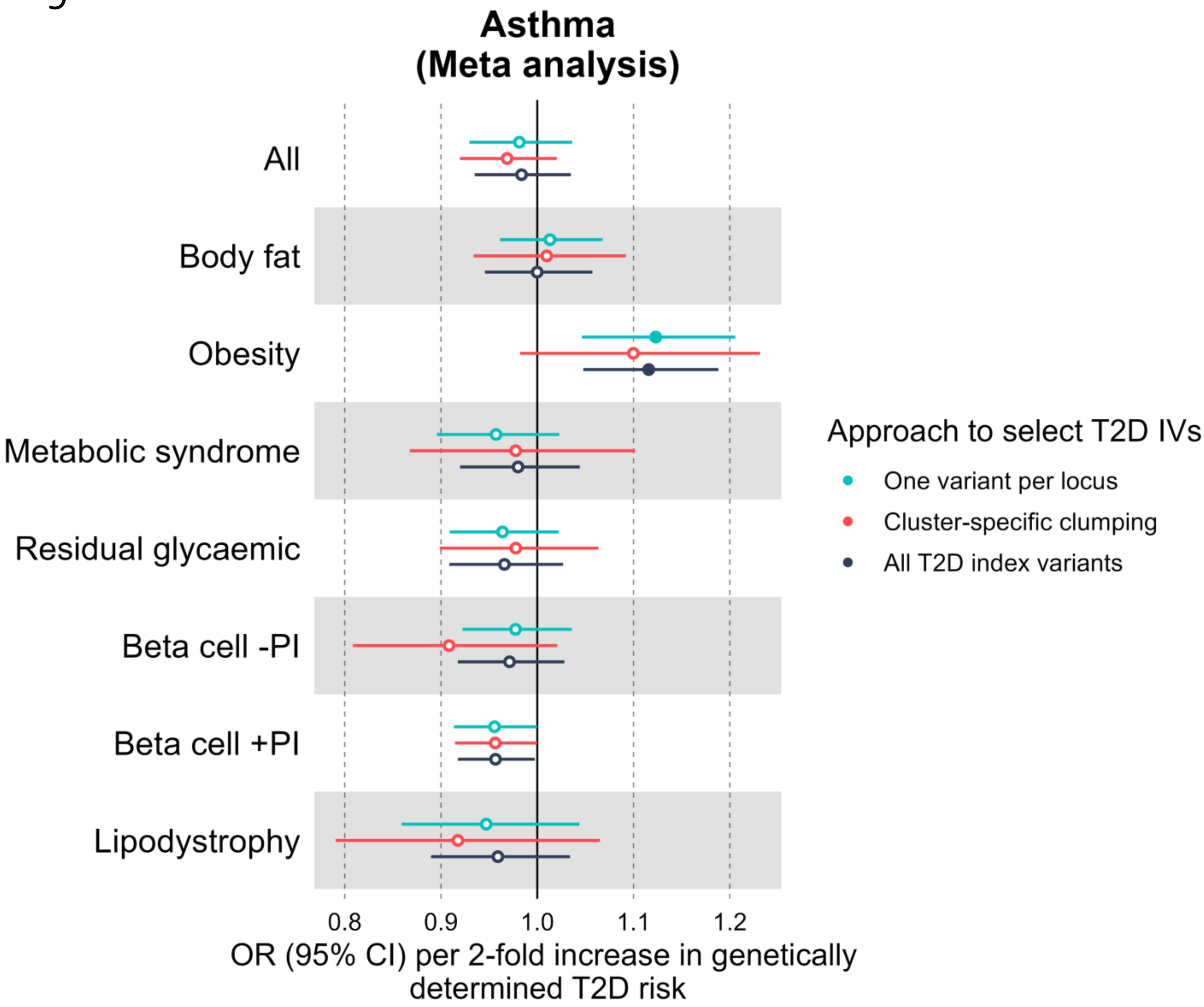
Asthma
(Meta analysis)
All
Body fat
Obesity
Metabolic syndrome
Residual glycaemic
Beta cell -PI
Beta cell +PI
Lipodystrophy
0.8
0.9
1.0
1.1
1.2
OR (95% CI) per 2-fold increase in genetically determined T2D risk
Approach to select T2D IVs
One variant per locus
Cluster-specific clumping
All T2D index variants

10

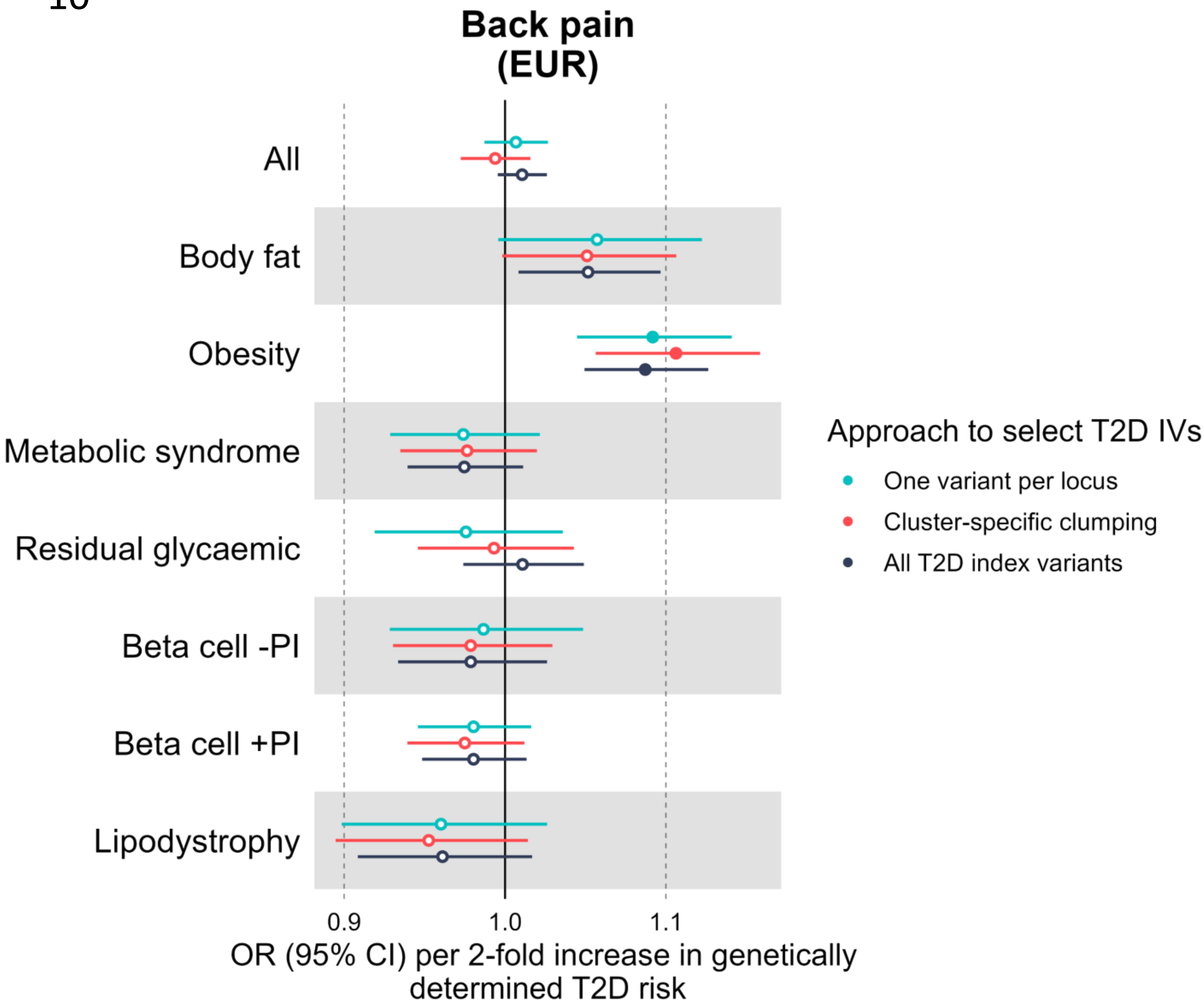

Back pain
(EUR)
All
Body fat
Obesity
Metabolic syndrome
Residual glycaemic
Beta cell -PI
Beta cell +PI
Lipodystrophy
0.9
1.0
1.1
OR (95% CI) per 2-fold increase in genetically determined T2D risk
Approach to select T2D IVs
One variant per locus
Cluster-specific clumping
All T2D index variants

11

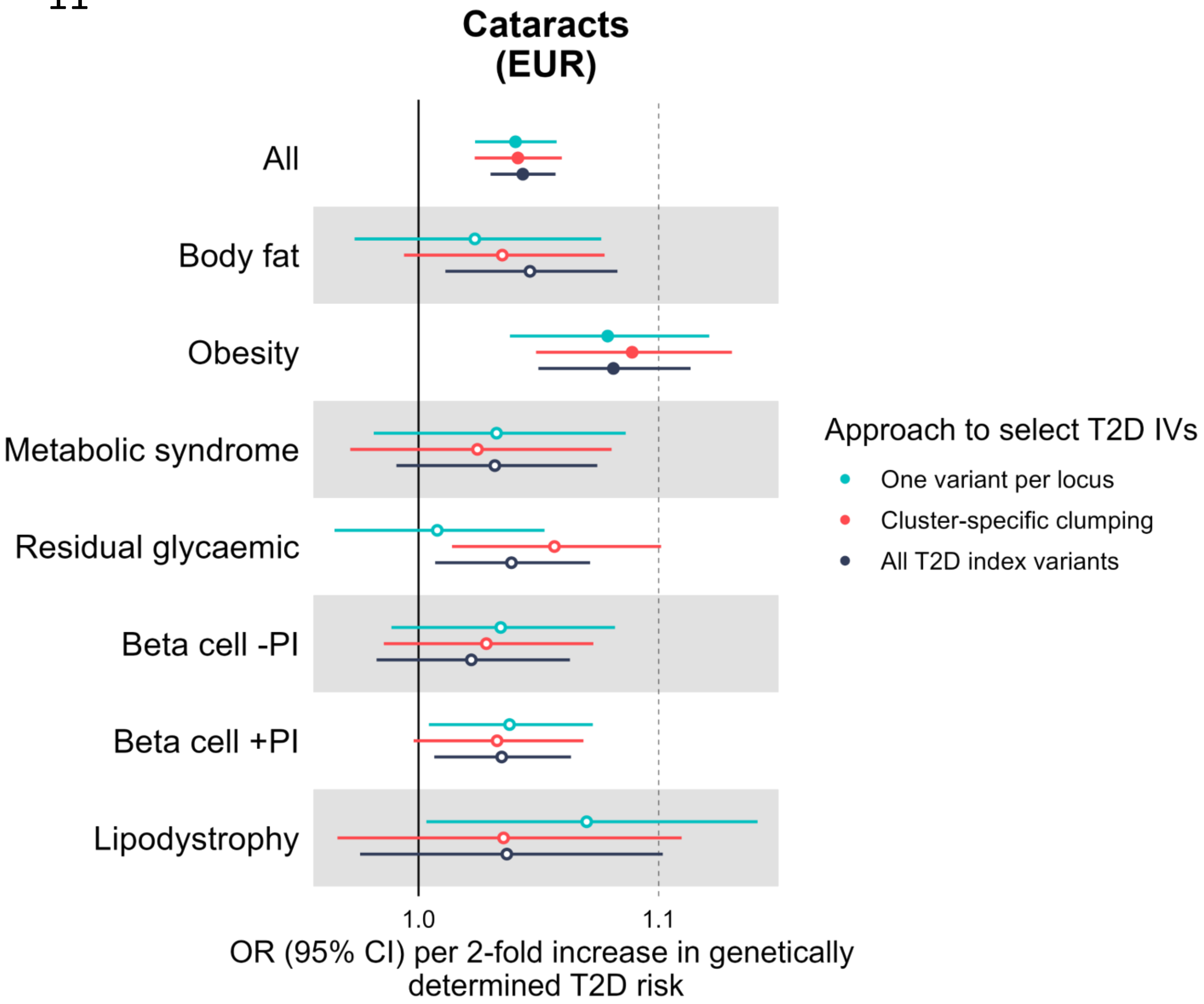

Cataracts
(EUR)
All
Body fat
Obesity
Metabolic syndrome
Residual glycaemic
Beta cell -PI
Beta cell +PI
Lipodystrophy
1.0
1.1
OR (95% CI) per 2-fold increase in genetically determined T2D risk
Approach to select T2D IVs
One variant per locus
Cluster-specific clumping
All T2D index variants

12

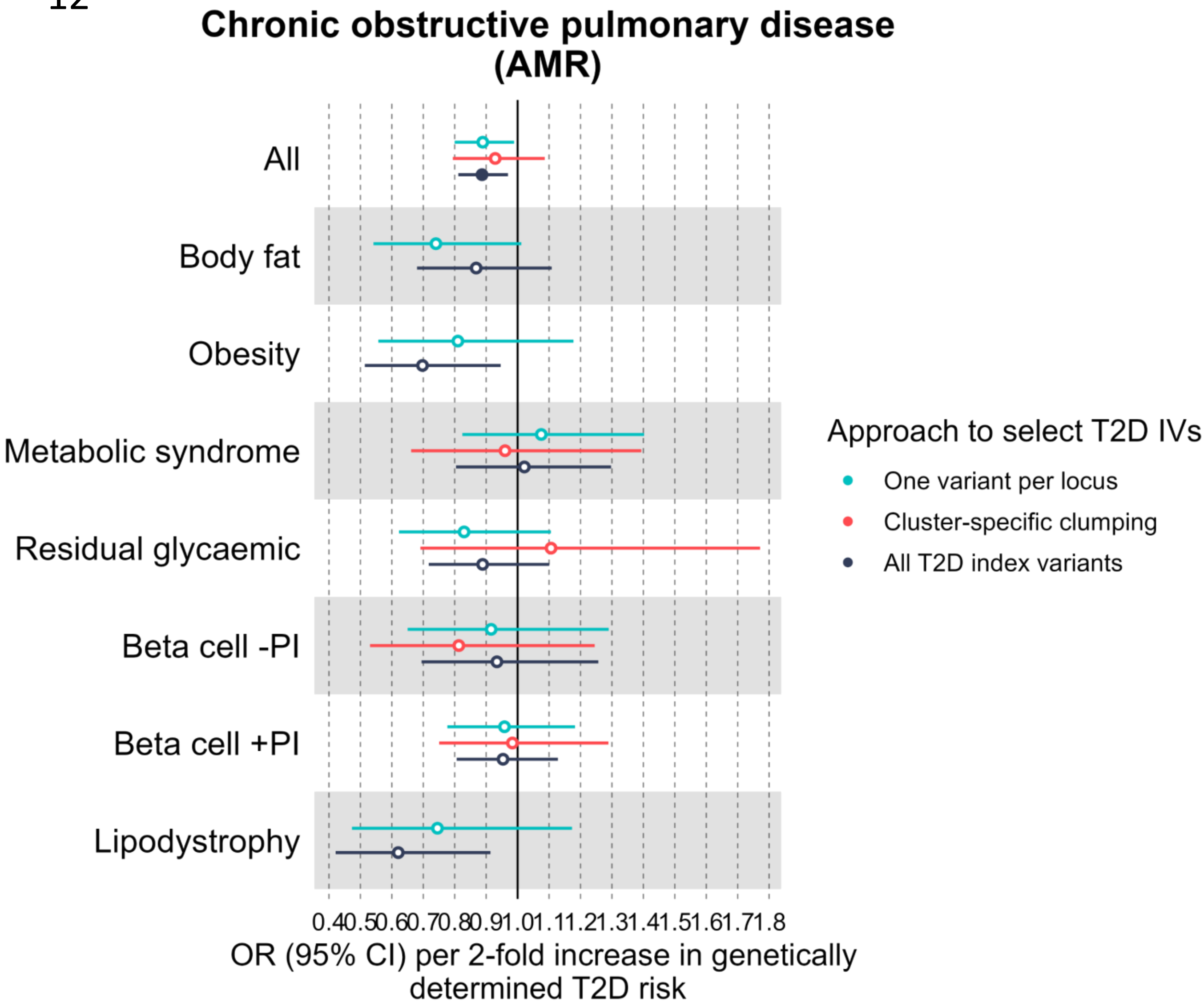
Chronic obstructive pulmonary disease
(AMR)
All
Body fat
Obesity
Metabolic syndrome
Residual glycaemic
Beta cell -PI
Beta cell +PI
Lipodystrophy
0.4 0.5 0.6 0.7 0.8 0.9 1.0 1.1 1.2 1.3 1.4 1.5 1.6 1.7 1.8
OR (95% CI) per 2-fold increase in genetically determined T2D risk
Approach to select T2D IVs
One variant per locus
Cluster-specific clumping
All T2D index variants

13

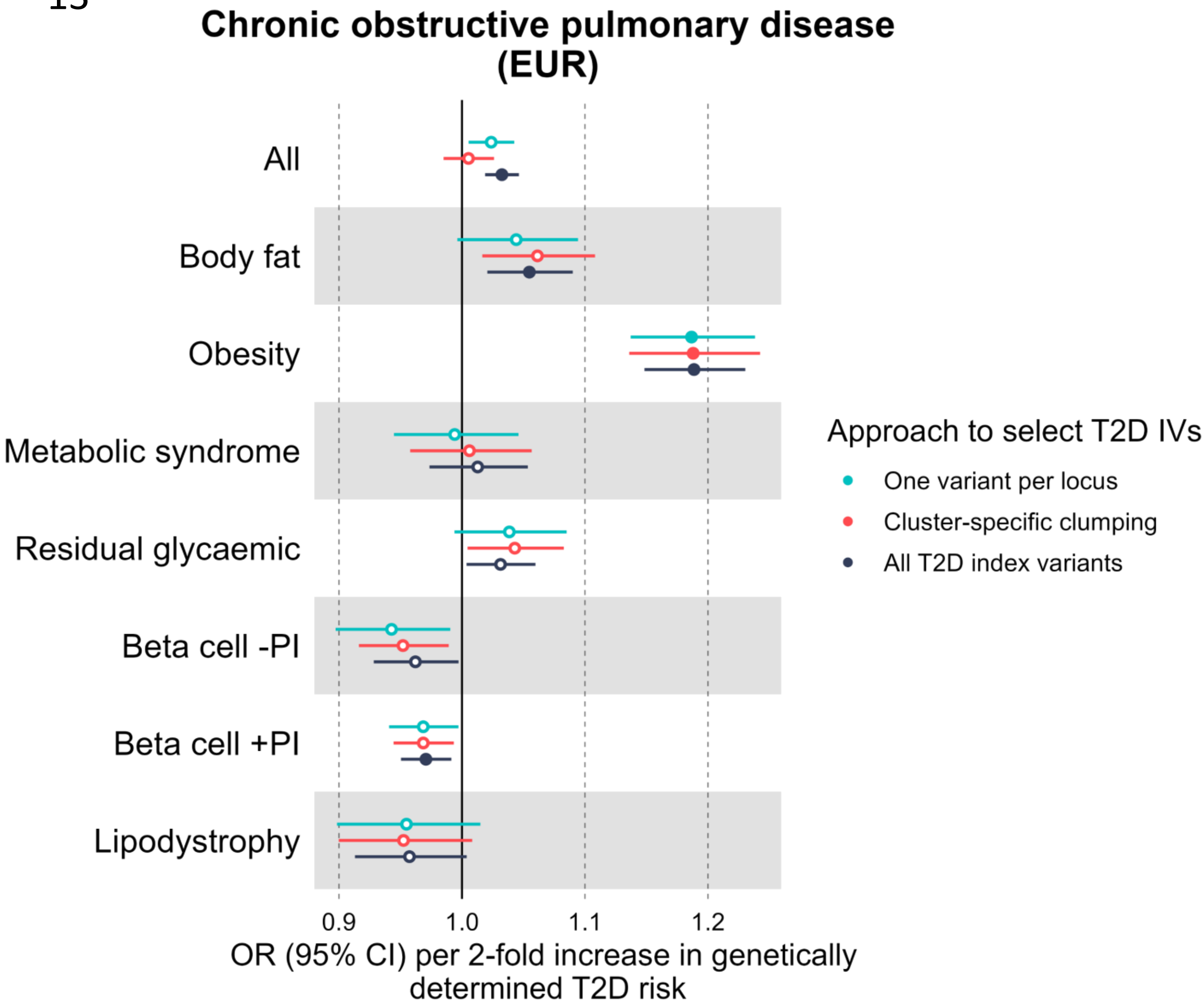

Chronic obstructive pulmonary disease
(EUR)
All
Body fat
Obesity
Metabolic syndrome
Residual glycaemic
Beta cell -PI
Beta cell +PI
Lipodystrophy
0.9
1.0
1.1
1.2
OR (95% CI) per 2-fold increase in genetically determined T2D risk
Approach to select T2D IVs
One variant per locus
Cluster-specific clumping
All T2D index variants

14

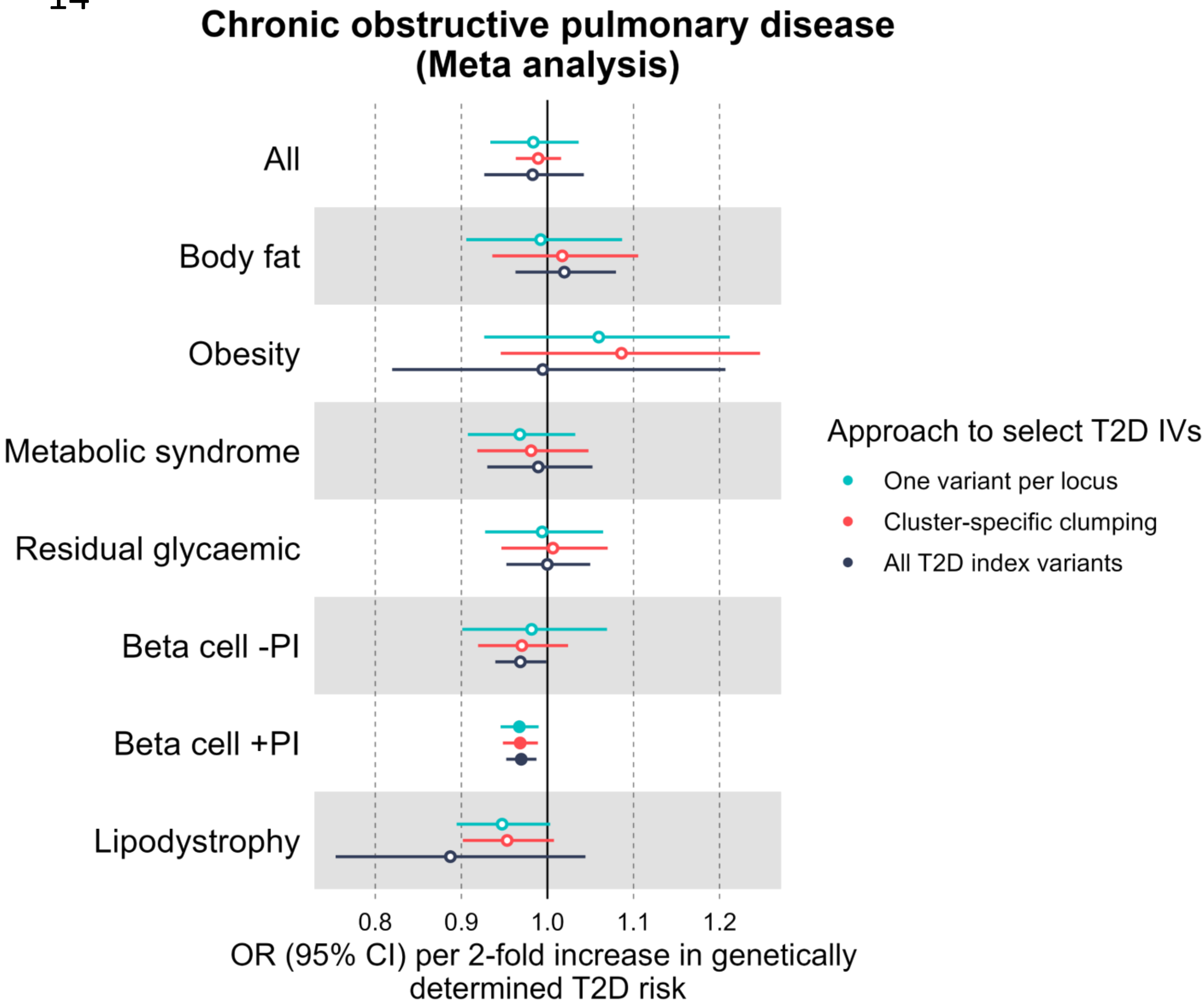
Chronic obstructive pulmonary disease
(Meta analysis)
All
Body fat
Obesity
Metabolic syndrome
Residual glycaemic
Beta cell -PI
Beta cell +PI
Lipodystrophy
0.8
0.9
1.0
1.1
1.2
OR (95% CI) per 2-fold increase in genetically
determined T2D risk
Approach to select T2D IVs
One variant per locus
Cluster-specific clumping
All T2D index variants

15

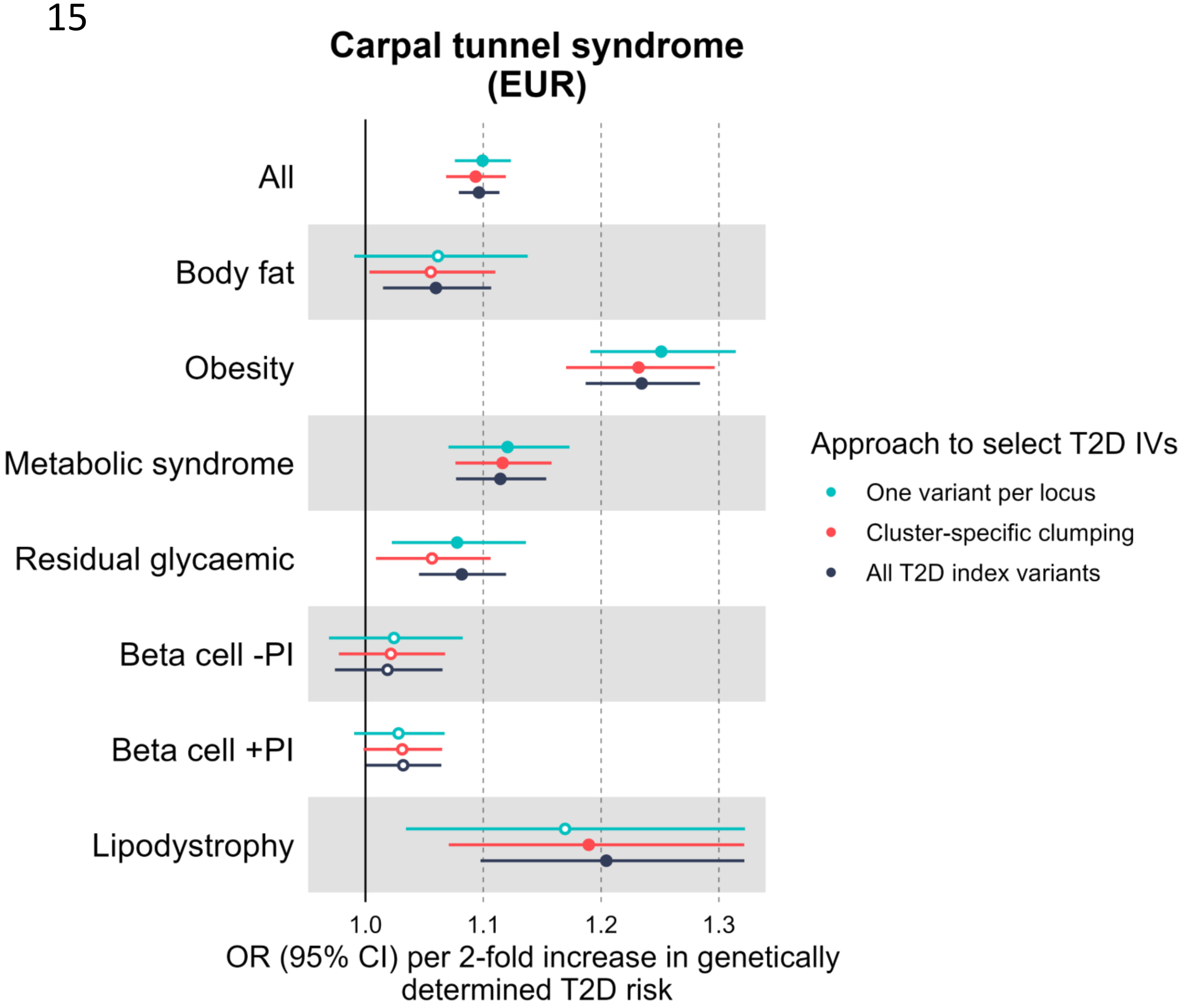

Carpal tunnel syndrome
(EUR)
All
Body fat
Obesity
Metabolic syndrome
Residual glycaemic
Beta cell -PI
Beta cell +PI
Lipodystrophy
1.0
1.1
1.2
1.3
OR (95% CI) per 2-fold increase in genetically determined T2D risk
Approach to select T2D IVs
One variant per locus
Cluster-specific clumping
All T2D index variants

16

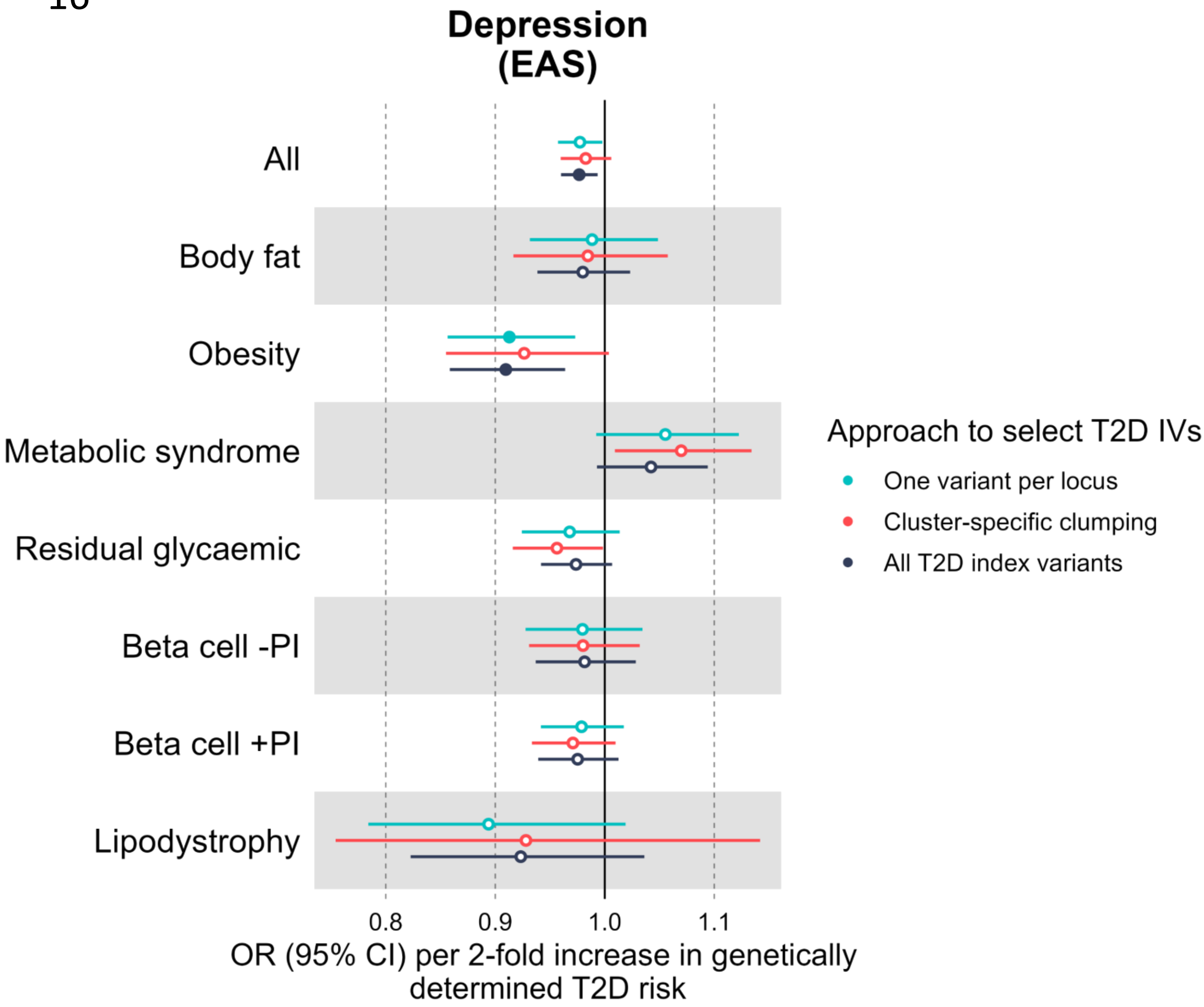

Depression
(EAS)
All
Body fat
Obesity
Metabolic syndrome
Residual glycaemic
Beta cell -PI
Beta cell +PI
Lipodystrophy
0.8
0.9
1.0
1.1
OR (95% CI) per 2-fold increase in genetically determined T2D risk
Approach to select T2D IVs
One variant per locus
Cluster-specific clumping
All T2D index variants

17

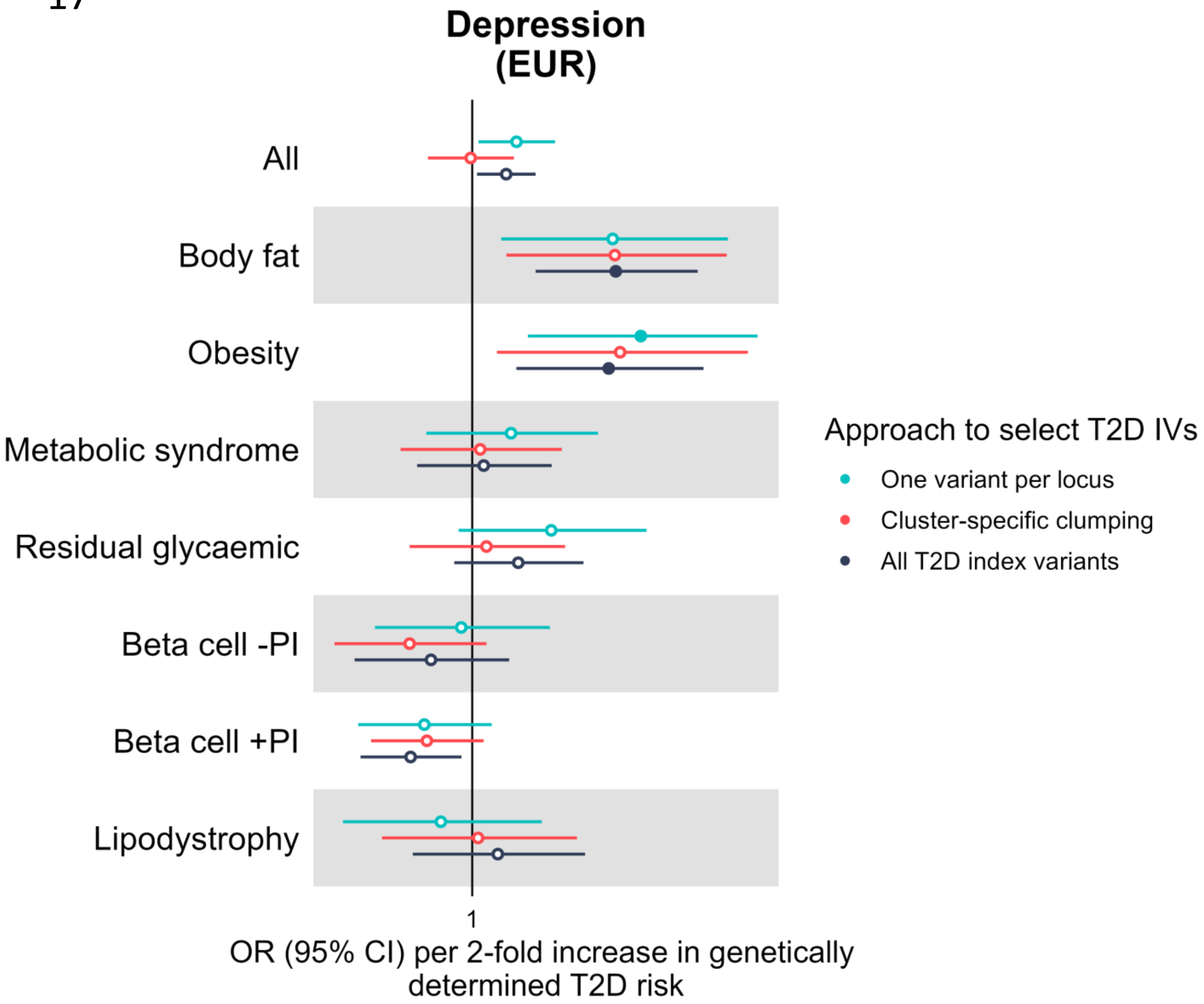

Depression
(EUR)
All
Body fat
Obesity
Metabolic syndrome
Residual glycaemic
Beta cell -PI
Beta cell +PI
Lipodystrophy
1
OR (95% CI) per 2-fold increase in genetically determined T2D risk
Approach to select T2D IVs
One variant per locus
Cluster-specific clumping
All T2D index variants

18

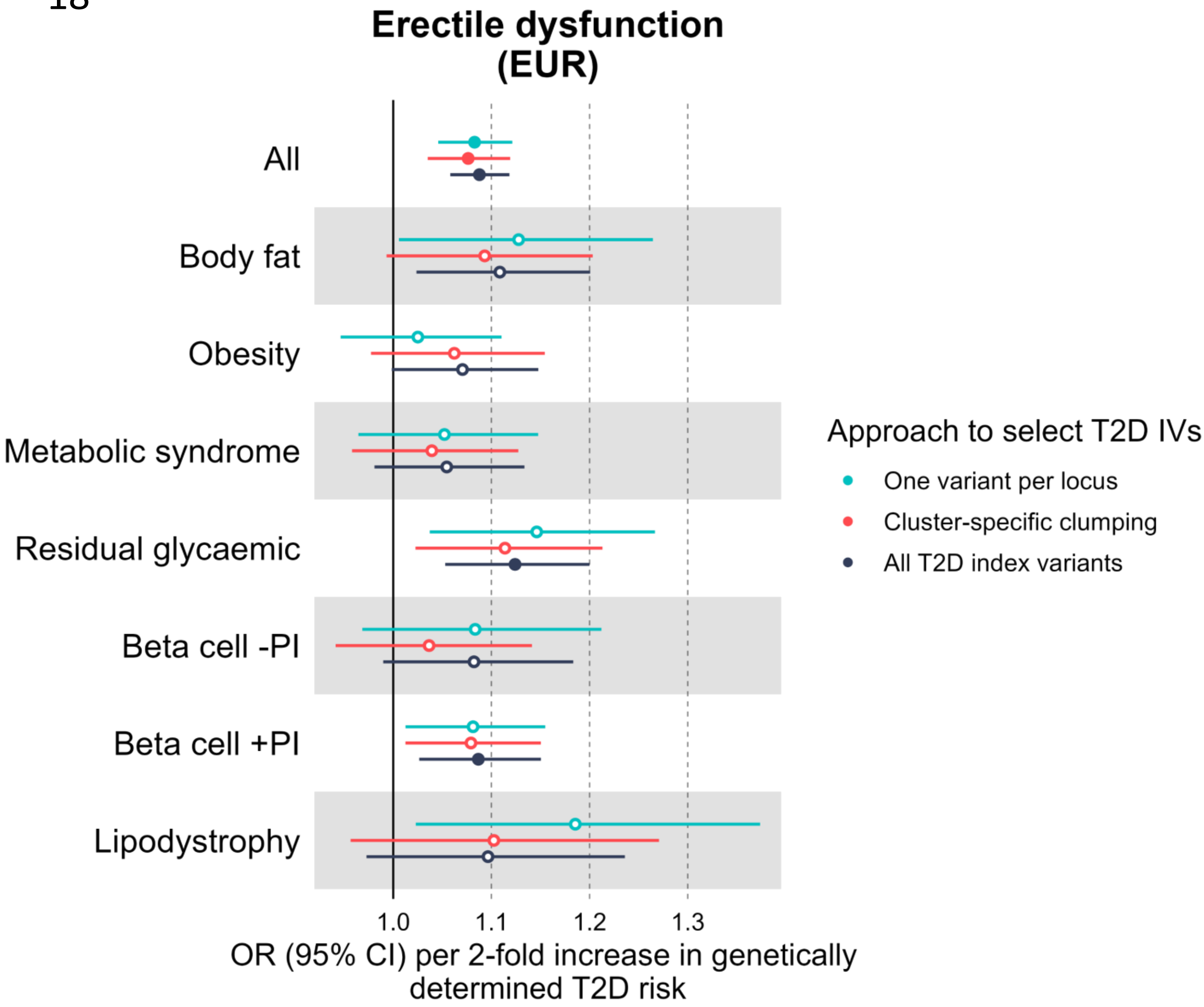
Erectile dysfunction
(EUR)
All
Body fat
Obesity
Metabolic syndrome
Residual glycaemic
Beta cell -PI
Beta cell +PI
Lipodystrophy
1.0
1.1
1.2
1.3
OR (95% CI) per 2-fold increase in genetically determined T2D risk
Approach to select T2D IVs
One variant per locus
Cluster-specific clumping
All T2D index variants

19

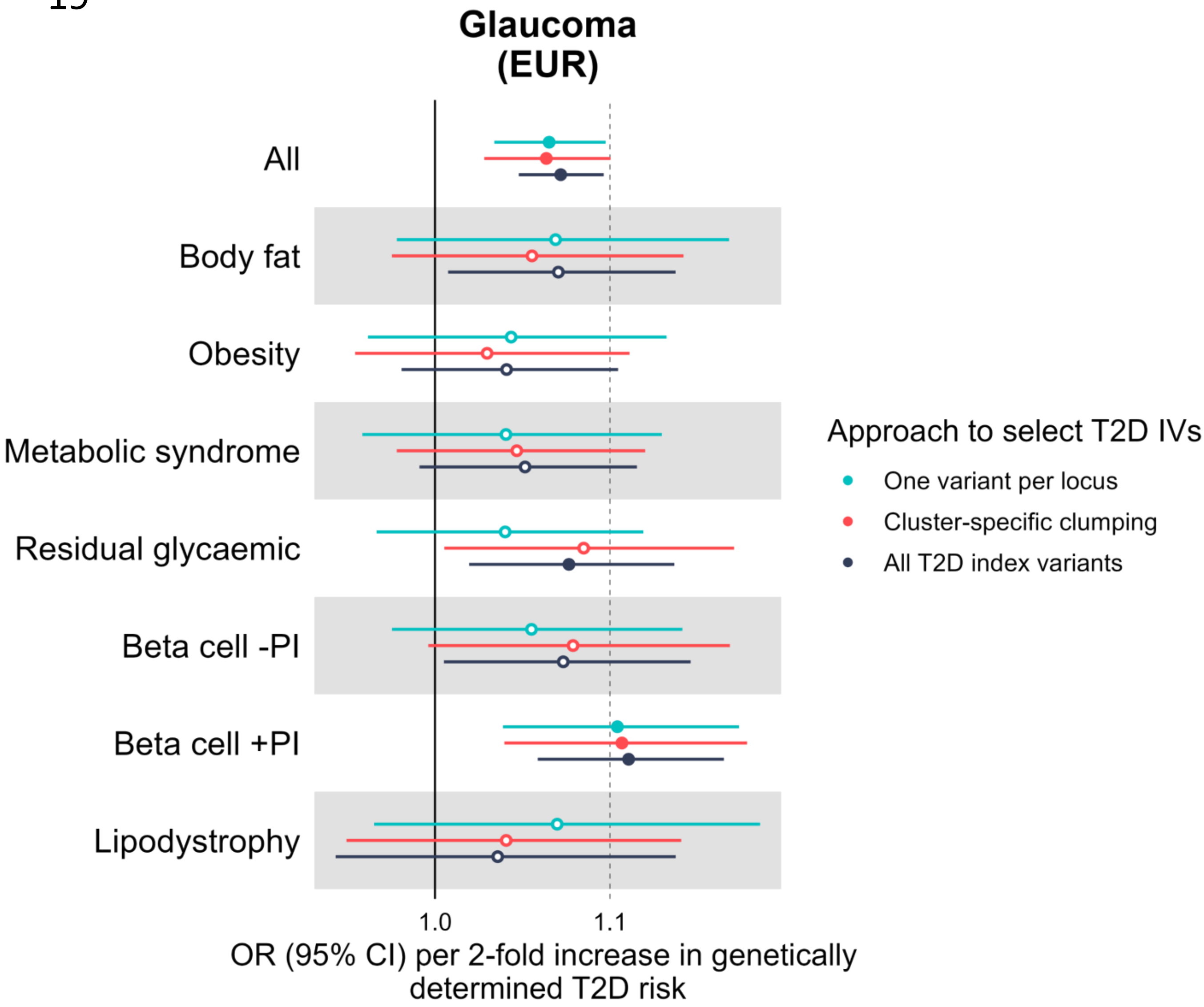
Glaucoma
(EUR)
All
Body fat
Obesity
Metabolic syndrome
Residual glycaemic
Beta cell -PI
Beta cell +PI
Lipodystrophy
1.0
1.1
OR (95% CI) per 2-fold increase in genetically determined T2D risk
Approach to select T2D IVs
One variant per locus
Cluster-specific clumping
All T2D index variants

20

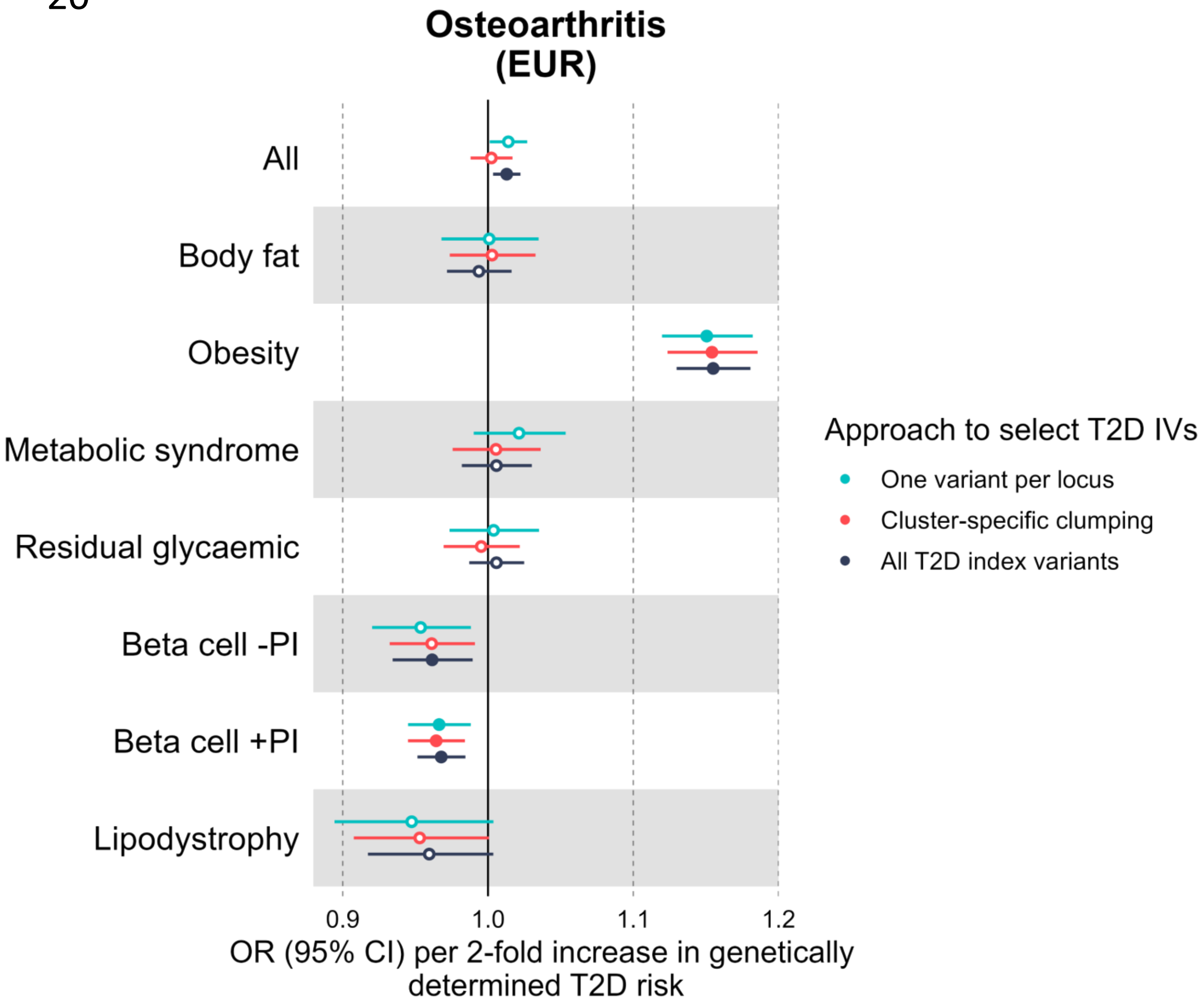

Osteoarthritis
(EUR)
All
Body fat
Obesity
Metabolic syndrome
Residual glycaemic
Beta cell -PI
Beta cell +PI
Lipodystrophy
0.9
1.0
1.1
1.2
OR (95% CI) per 2-fold increase in genetically determined T2D risk
Approach to select T2D IVs
One variant per locus
Cluster-specific clumping
All T2D index variants

21

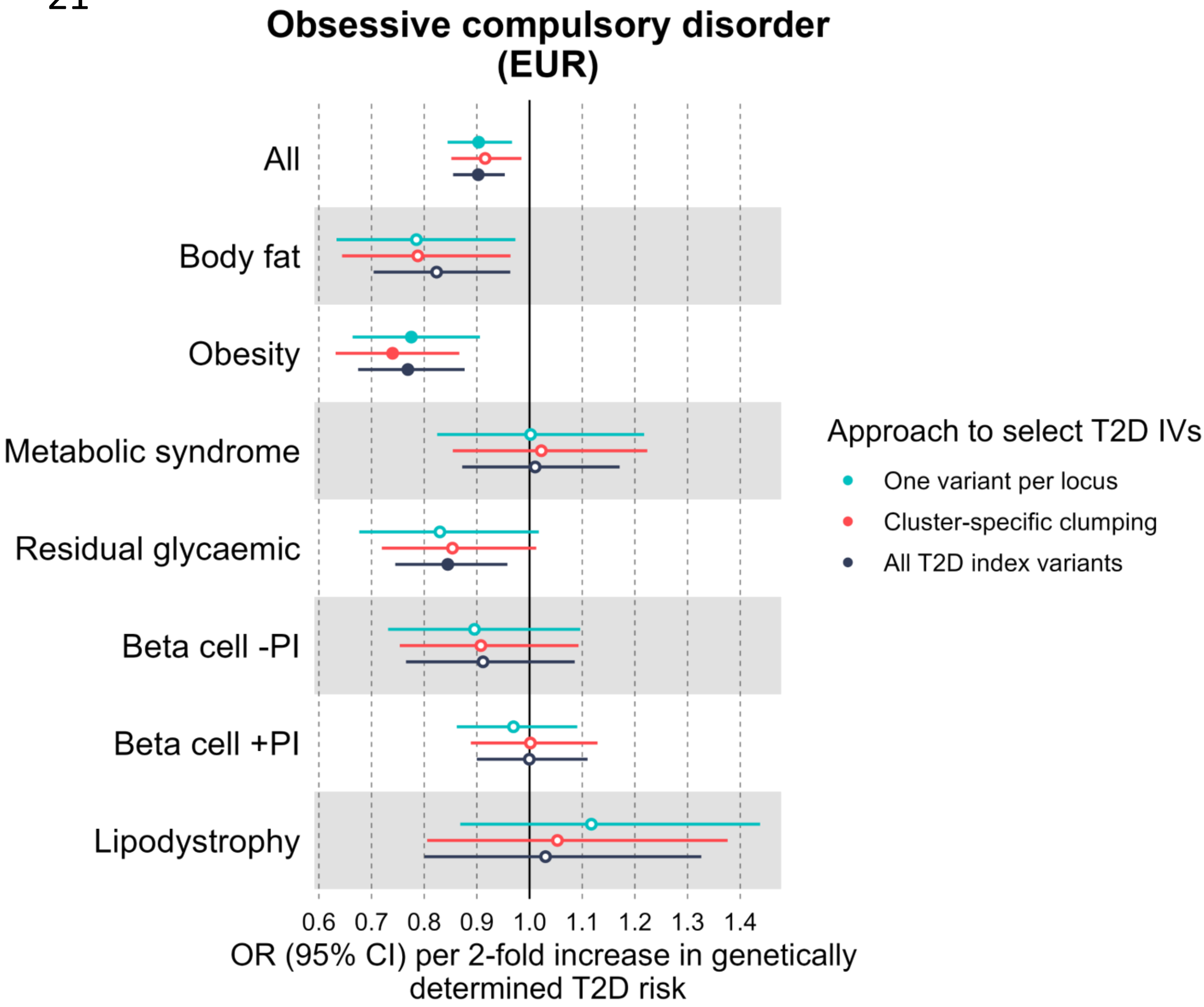

Obsessive compulsory disorder
(EUR)
All
Body fat
Obesity
Metabolic syndrome
Residual glycaemic
Beta cell -PI
Beta cell +PI
Lipodystrophy
0.6 0.7 0.8 0.9 1.0 1.1 1.2 1.3 1.4
OR (95% CI) per 2-fold increase in genetically determined T2D risk
Approach to select T2D IVs
One variant per locus
Cluster-specific clumping
All T2D index variants

22

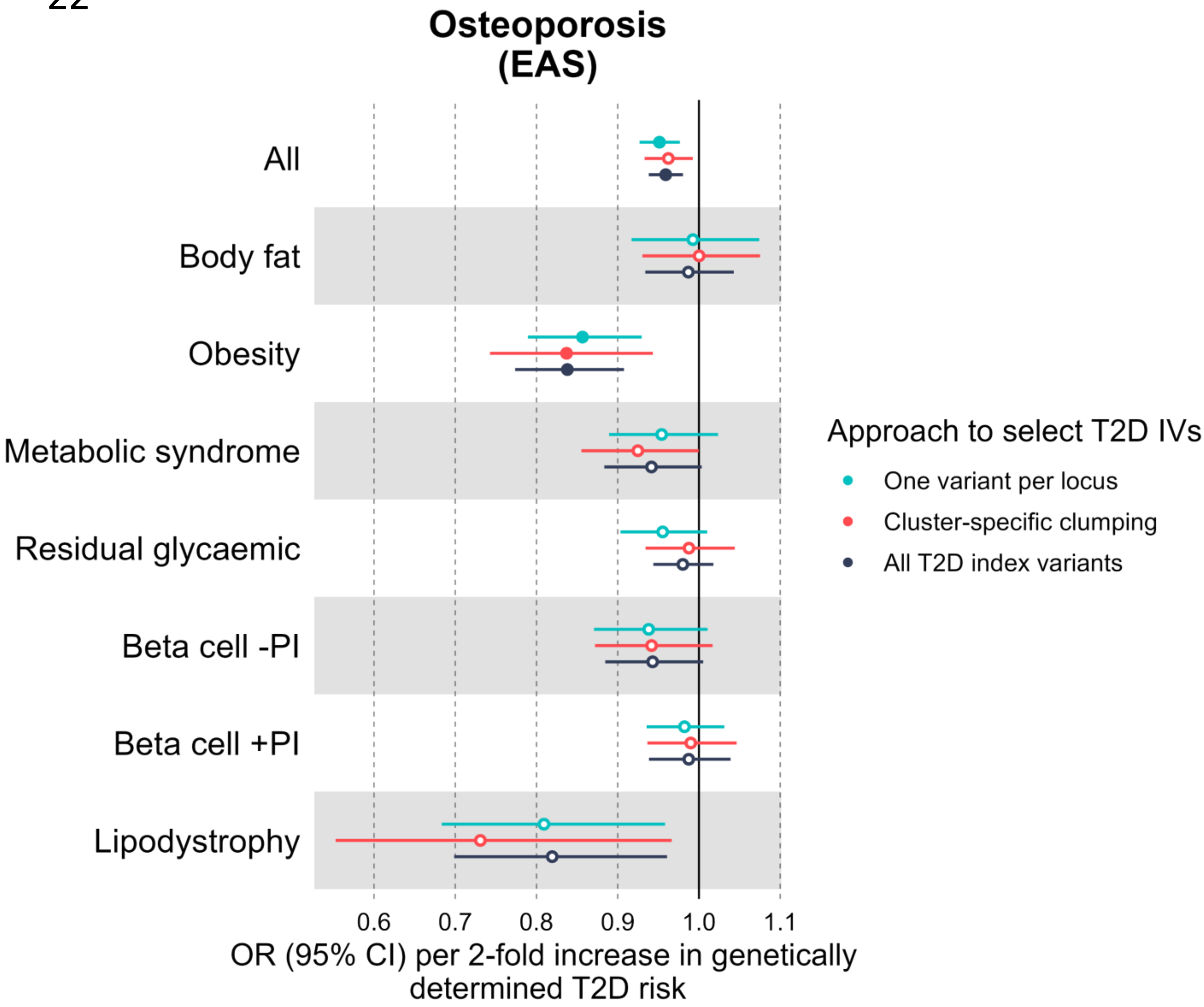

Osteoporosis
(EAS)
All
Body fat
Obesity
Metabolic syndrome
Residual glycaemic
Beta cell -PI
Beta cell +PI
Lipodystrophy
0.6
0.7
0.8
0.9
1.0
1.1
OR (95% CI) per 2-fold increase in genetically determined T2D risk
Approach to select T2D IVs
One variant per locus
Cluster-specific clumping
All T2D index variants

23

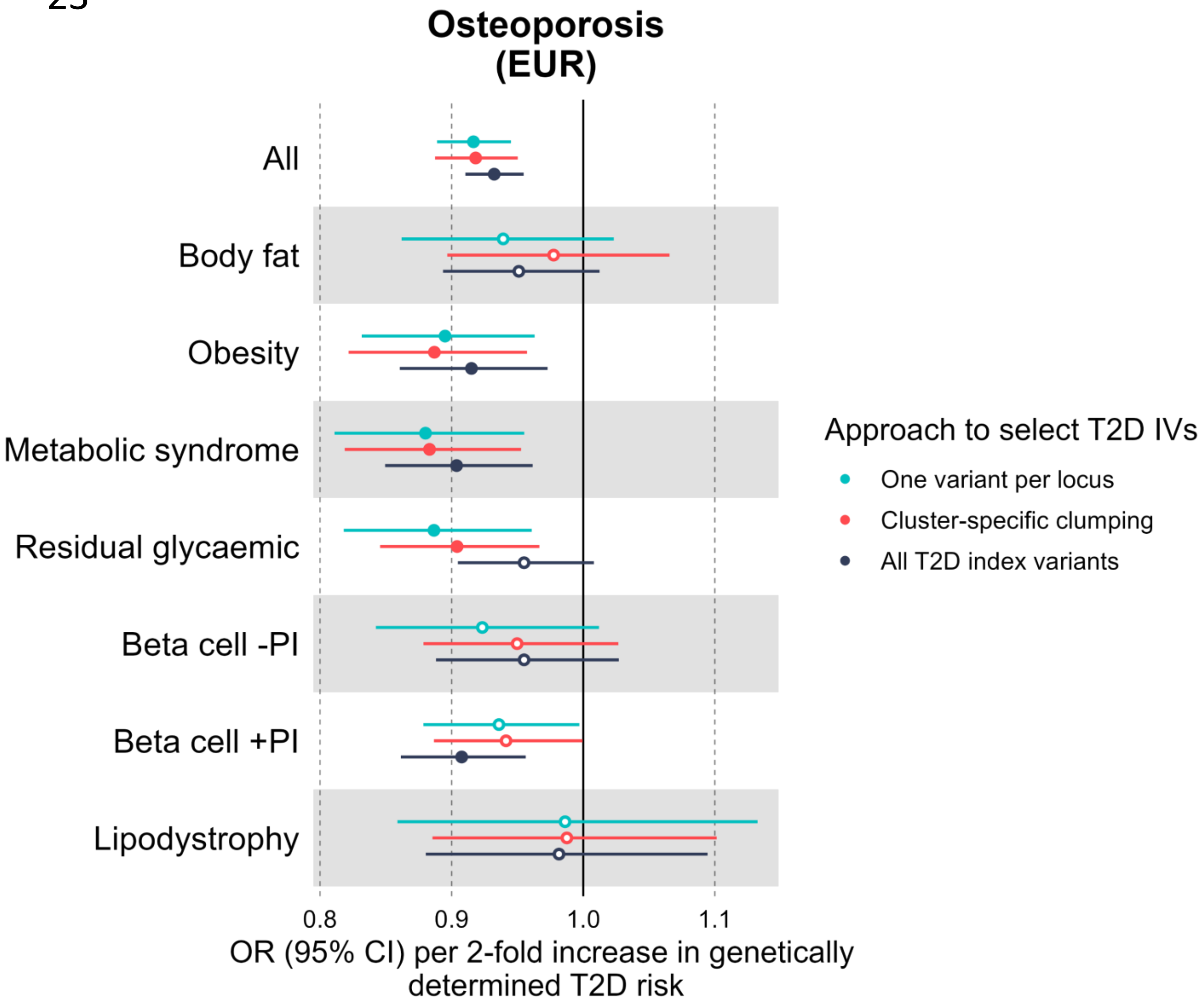

Osteoporosis
(EUR)
All
Body fat
Obesity
Metabolic syndrome
Residual glycaemic
Beta cell -PI
Beta cell +PI
Lipodystrophy
0.8
0.9
1.0
1.1
OR (95% CI) per 2-fold increase in genetically determined T2D risk
Approach to select T2D IVs
One variant per locus
Cluster-specific clumping
All T2D index variants

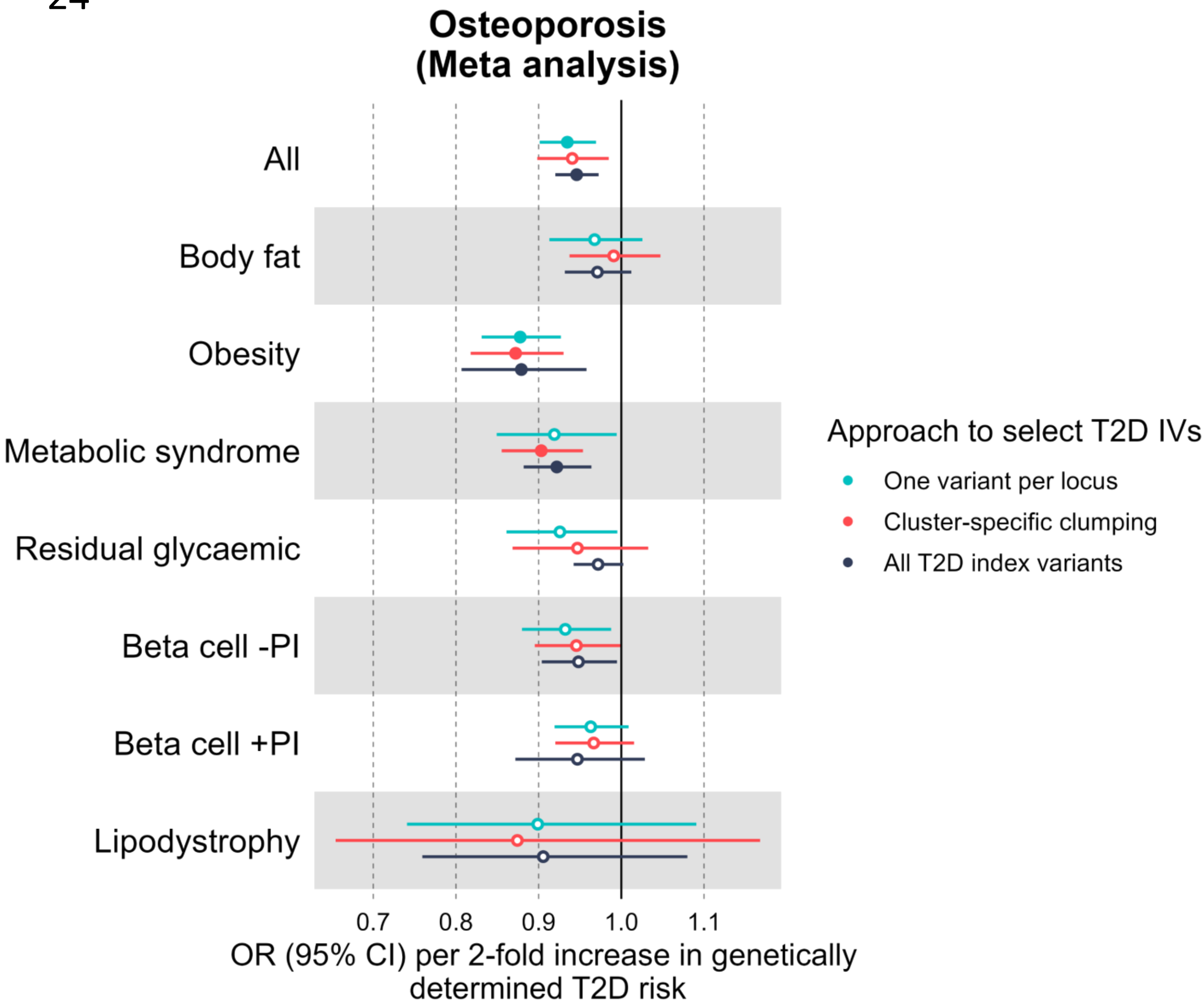
Osteoporosis
(Meta analysis)
All
Body fat
Obesity
Metabolic syndrome
Residual glycaemic
Beta cell -PI
Beta cell +PI
Lipodystrophy
0.7
0.8
0.9
1.0
1.1
OR (95% CI) per 2-fold increase in genetically determined T2D risk
Approach to select T2D IVs
One variant per locus
Cluster-specific clumping
All T2D index variants

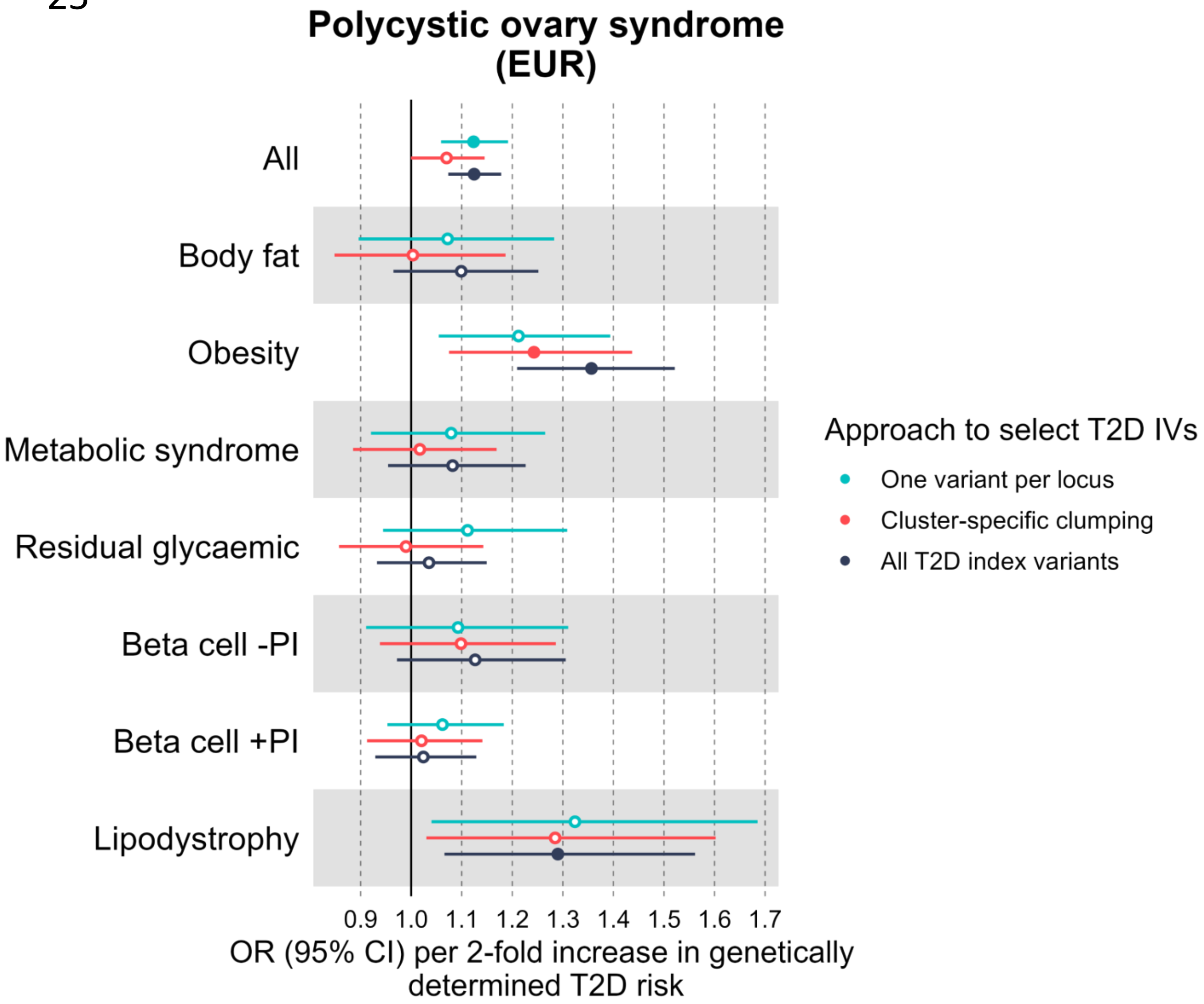
Polycystic ovary syndrome (EUR)
All
Body fat
Obesity
Metabolic syndrome
Residual glycaemic
Beta cell -PI
Beta cell +PI
Lipodystrophy
0.9 1.0 1.1 1.2 1.3 1.4 1.5 1.6 1.7
OR (95% CI) per 2-fold increase in genetically determined T2D risk
Approach to select T2D IVs
One variant per locus
Cluster-specific clumping
All T2D index variants

26

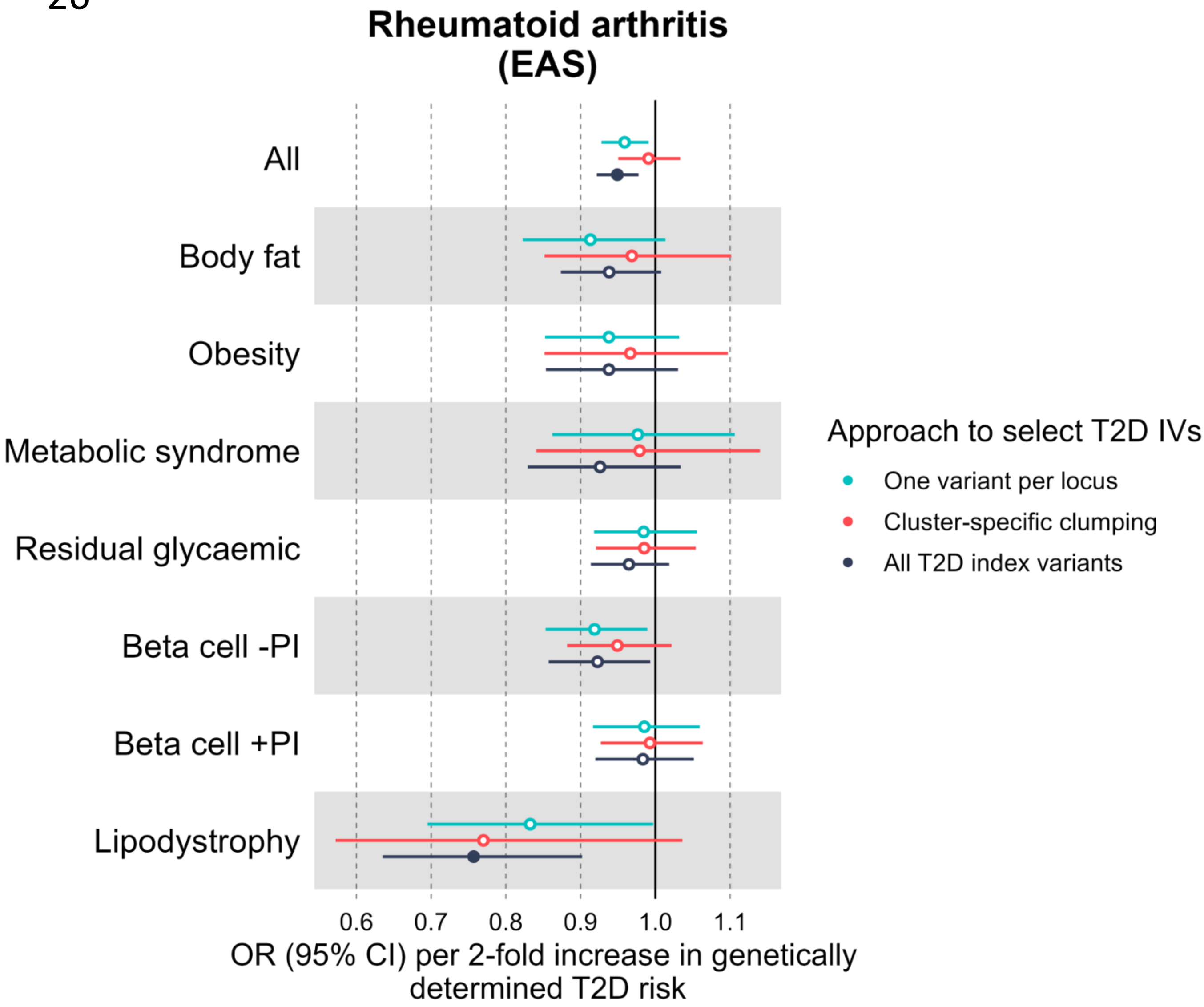
Rheumatoid arthritis
(EAS)
All
Body fat
Obesity
Metabolic syndrome
Residual glycaemic
Beta cell -PI
Beta cell +PI
Lipodystrophy
0.6
0.7
0.8
0.9
1.0
1.1
OR (95% CI) per 2-fold increase in genetically determined T2D risk
Approach to select T2D IVs
One variant per locus
Cluster-specific clumping
All T2D index variants

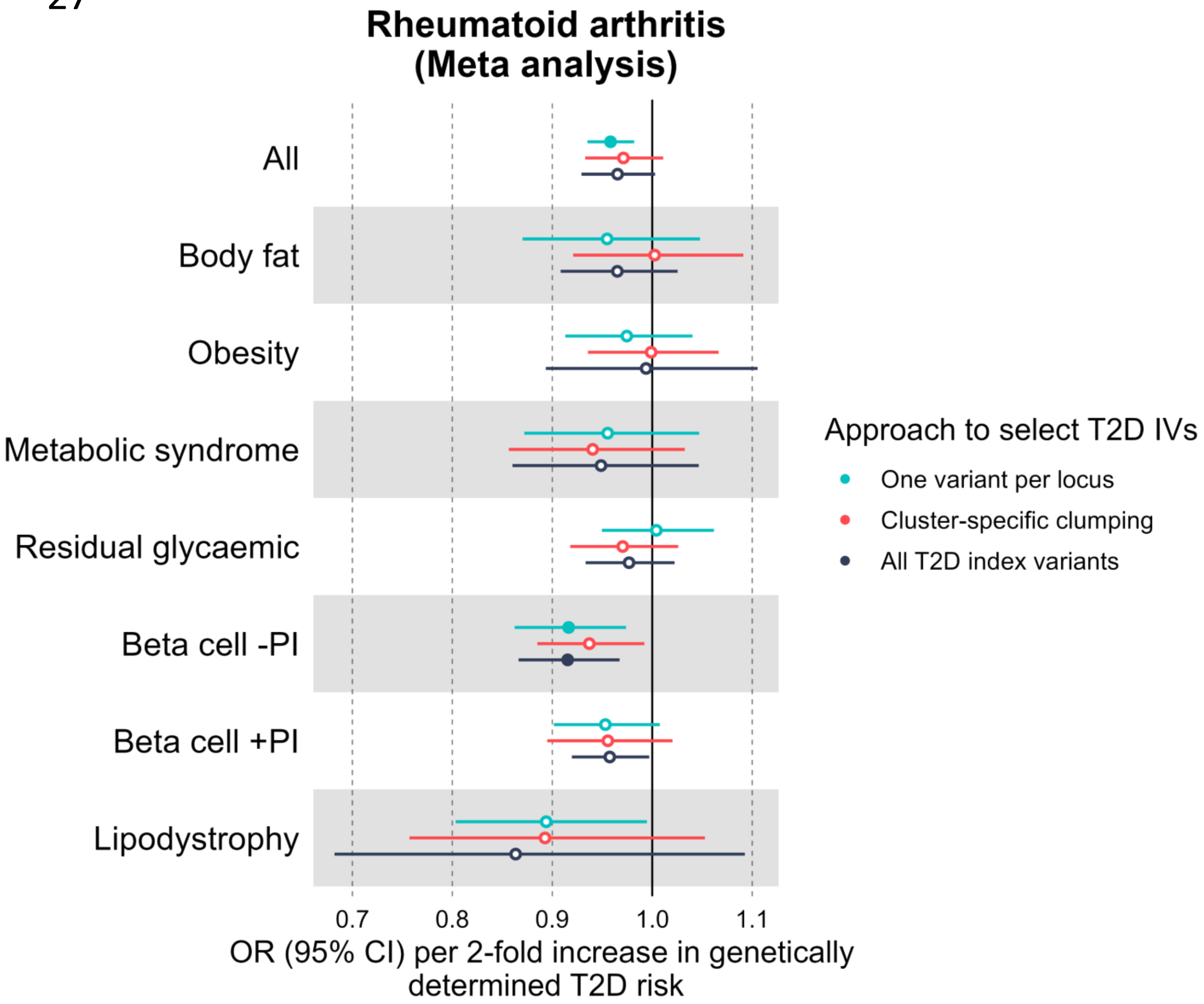
Rheumatoid arthritis
(Meta analysis)
All
Body fat
Obesity
Metabolic syndrome
Residual glycaemic
Beta cell -PI
Beta cell +PI
Lipodystrophy
0.7
0.8
0.9
1.0
1.1
OR (95% CI) per 2-fold increase in genetically determined T2D risk
Approach to select T2D IVs
One variant per locus
Cluster-specific clumping
All T2D index variants

**Supplemental Figures 28-48:** Forest plots comparing the results of the univariable and multivariable Mendelian randomization (MR) analyses using data from individuals genetically similar to Europeans. Causal estimates are expressed as the odds ratio of comorbidity risk per doubling (2-fold increase) in genetically determined dichotomous T2D risk. (HDL=high-density lipoprotein cholesterol; WHR = waist-to-hip ratio; BMI=body mass index; CI=confidence interval; FDR=false-discovery-rate at 5%).

28

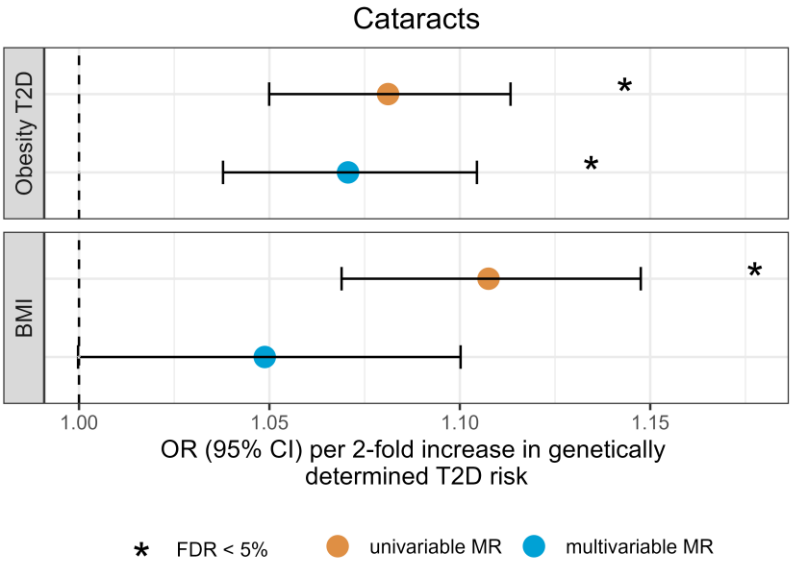
Cataracts
Obesity T2D
BMI
1.00
1.05
1.10
1.15
OR (95% CI) per 2-fold increase in genetically determined T2D risk
* FDR < 5%
univariable MR
multivariable MR

29

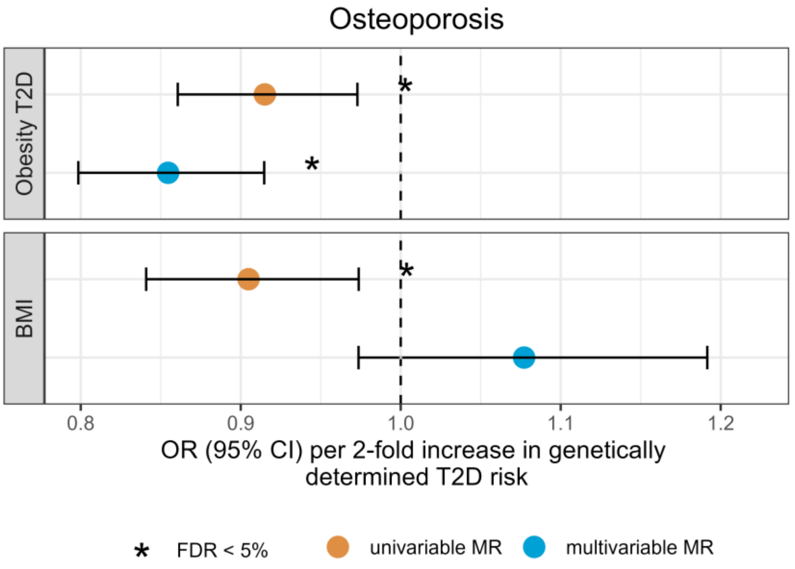
Osteoporosis
Obesity T2D
BMI
0.8
0.9
1.0
1.1
1.2
OR (95% CI) per 2-fold increase in genetically determined T2D risk
* FDR < 5%
univariable MR
multivariable MR

30

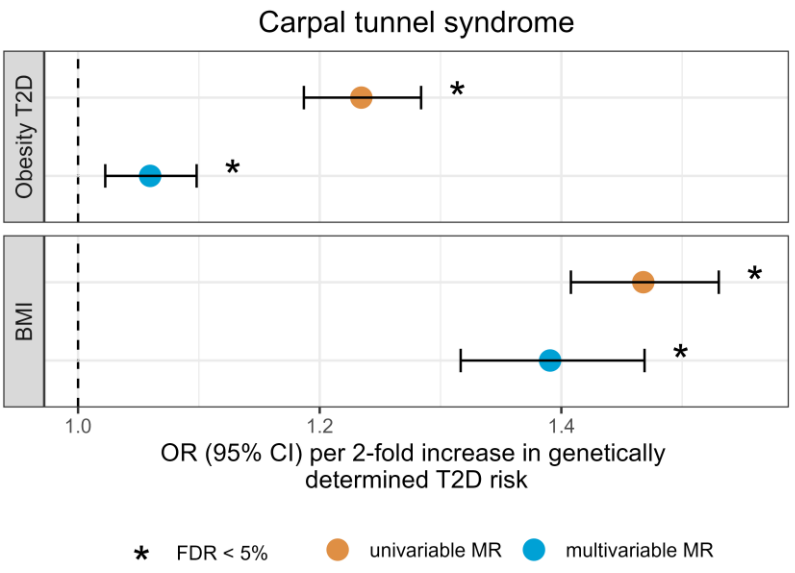
Carpal tunnel syndrome
Obesity T2D
BMI
1.0
1.2
1.4
OR (95% CI) per 2-fold increase in genetically determined T2D risk
* FDR < 5%
univariable MR
multivariable MR

31

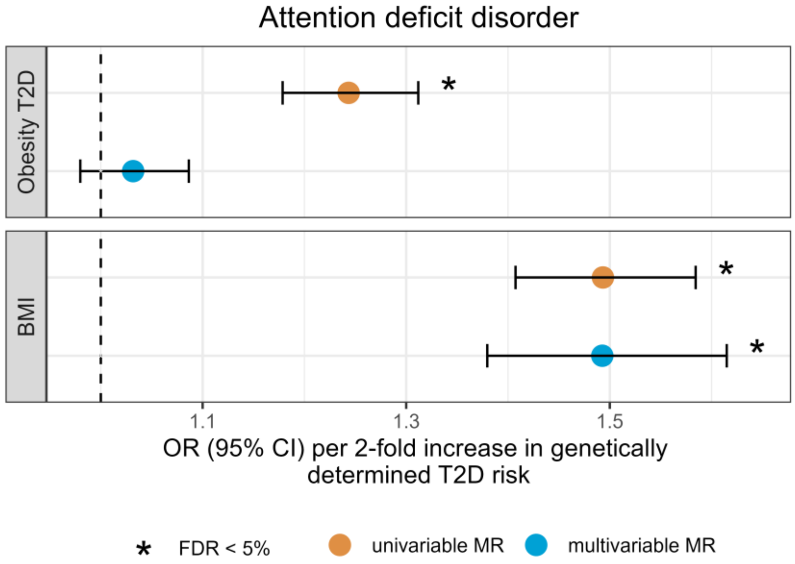
Attention deficit disorder
Obesity T2D
BMI
1.1
1.3
1.5
OR (95% CI) per 2-fold increase in genetically determined T2D risk
* FDR < 5%
univariable MR
multivariable MR

32

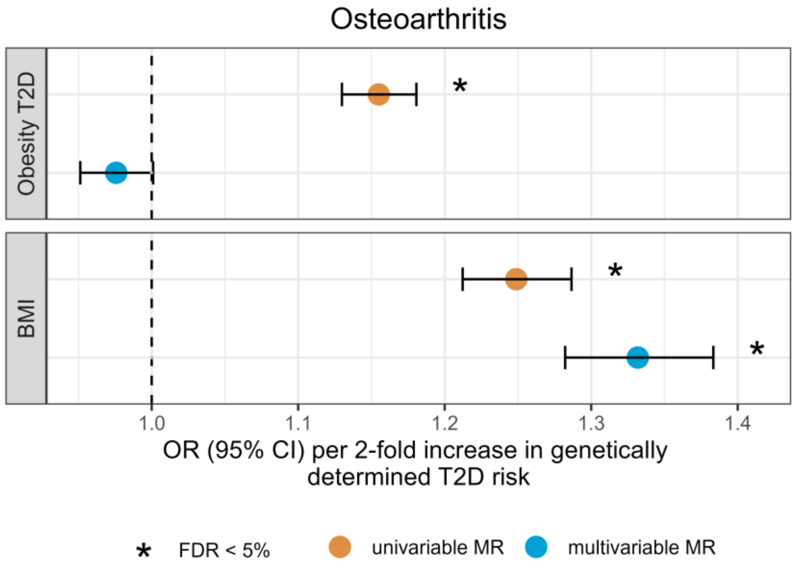
Osteoarthritis
Obesity T2D
BMI
1.0
1.1
1.2
1.3
1.4
OR (95% CI) per 2-fold increase in genetically determined T2D risk
* FDR < 5%
univariable MR
multivariable MR

33

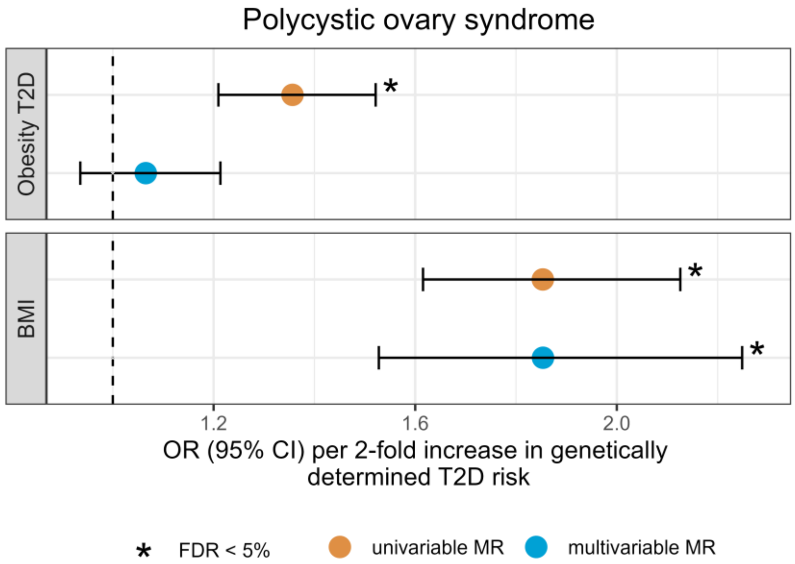
Polycystic ovary syndrome
Obesity T2D
BMI
1.2
1.6
2.0
OR (95% CI) per 2-fold increase in genetically determined T2D risk
* FDR < 5%
univariable MR
multivariable MR

34

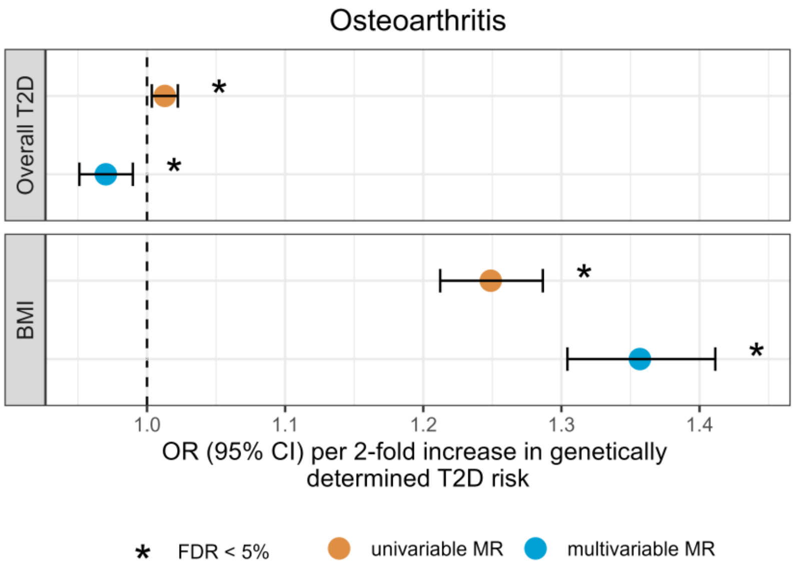
Osteoarthritis
Overall T2D
BMI
1.0
1.1
1.2
1.3
1.4
OR (95% CI) per 2-fold increase in genetically determined T2D risk
* FDR < 5%
univariable MR
multivariable MR

35

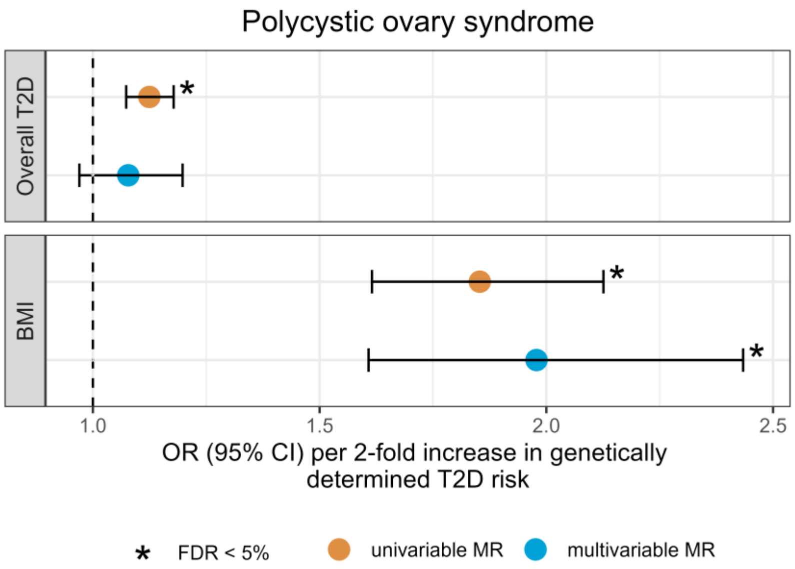
Polycystic ovary syndrome
Overall T2D
BMI
1.0
1.5
2.0
2.5
OR (95% CI) per 2-fold increase in genetically determined T2D risk
* FDR < 5%
univariable MR
multivariable MR

36

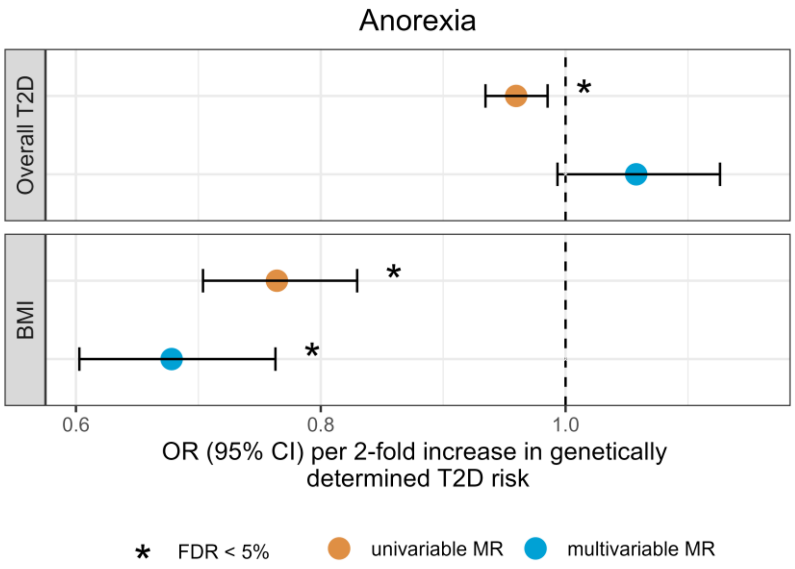
Anorexia
Overall T2D
BMI
0.6
0.8
1.0
OR (95% CI) per 2-fold increase in genetically determined T2D risk
* FDR < 5%
univariable MR
multivariable MR

37

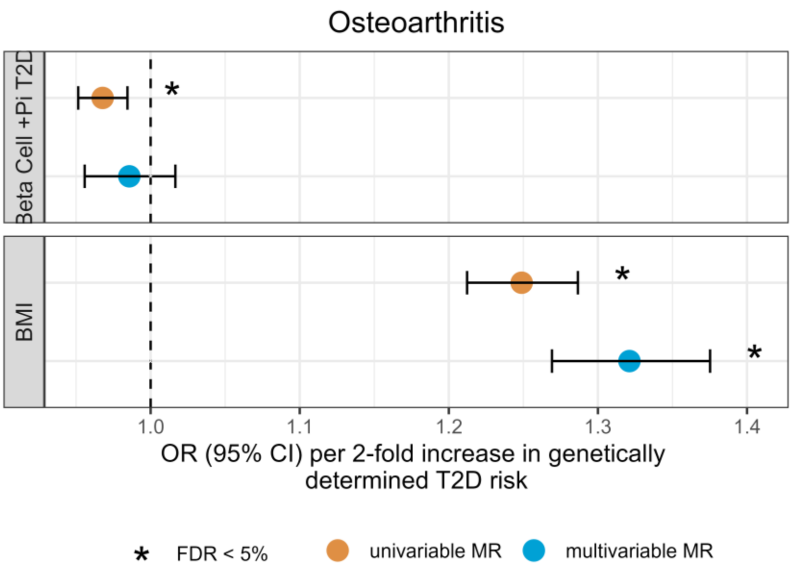
Osteoarthritis
Beta Cell +Pi T2D
BMI
1.0
1.1
1.2
1.3
1.4
OR (95% CI) per 2-fold increase in genetically determined T2D risk
FDR < 5%
univariable MR
multivariable MR

38

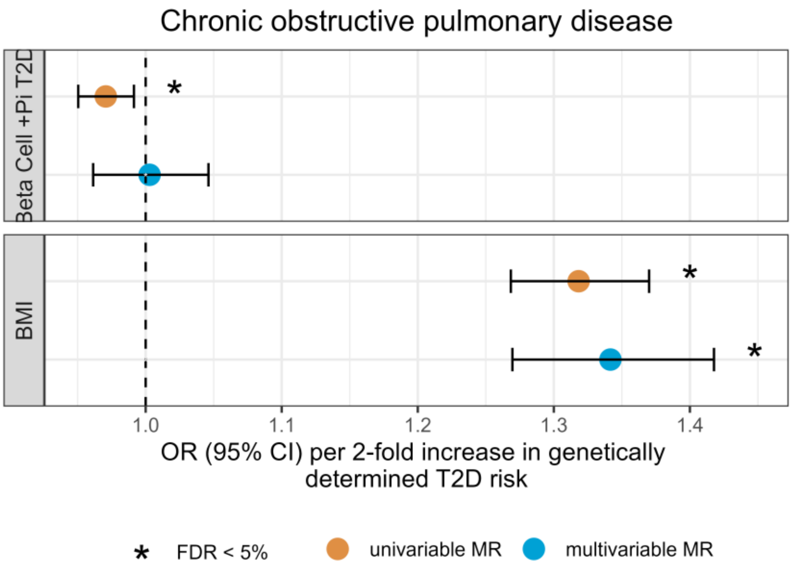
Chronic obstructive pulmonary disease
Beta Cell +Pi T2D
BMI
1.0
1.1
1.2
1.3
1.4
OR (95% CI) per 2-fold increase in genetically determined T2D risk
FDR < 5%
univariable MR
multivariable MR

39

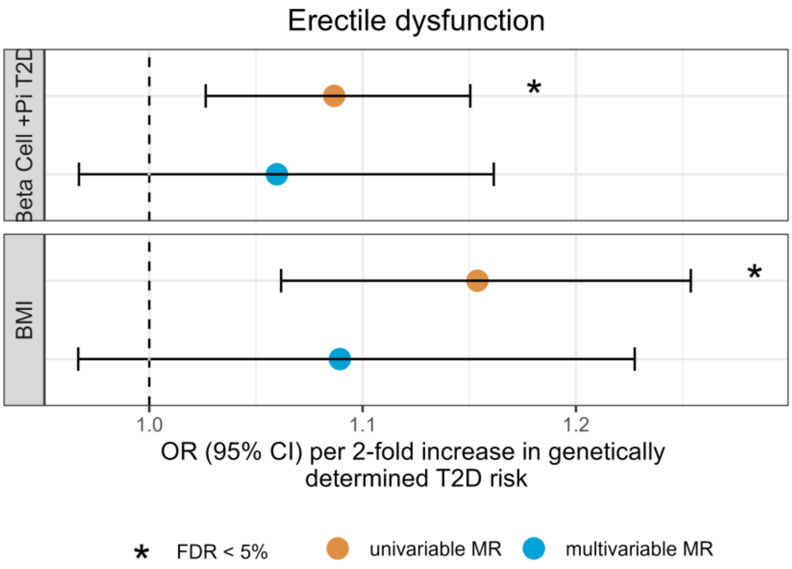
Erectile dysfunction
Beta Cell +Pi T2D
BMI
1.0
1.1
1.2
OR (95% CI) per 2-fold increase in genetically determined T2D risk
FDR < 5%
univariable MR
multivariable MR

40

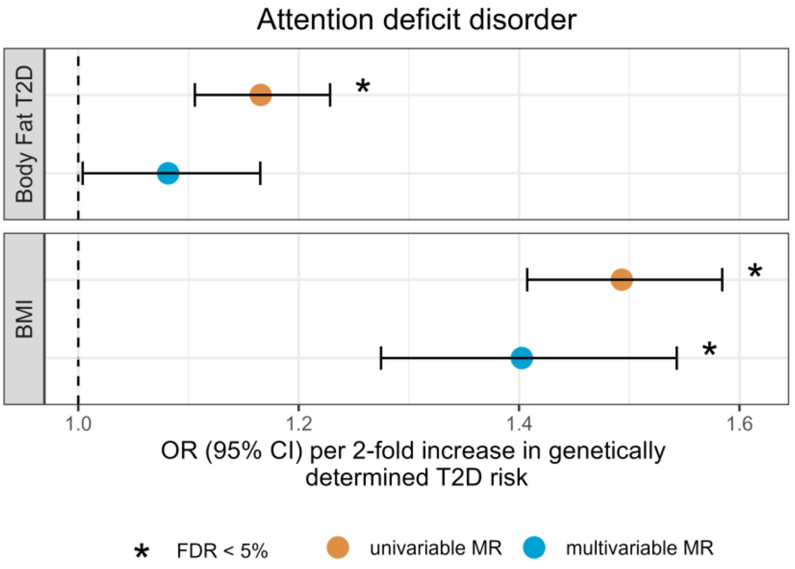

Attention deficit disorder
Body Fat T2D
BMI
*
1.0
1.2
1.4
1.6
OR (95% CI) per 2-fold increase in genetically determined T2D risk
* FDR < 5%
univariable MR
multivariable MR


41

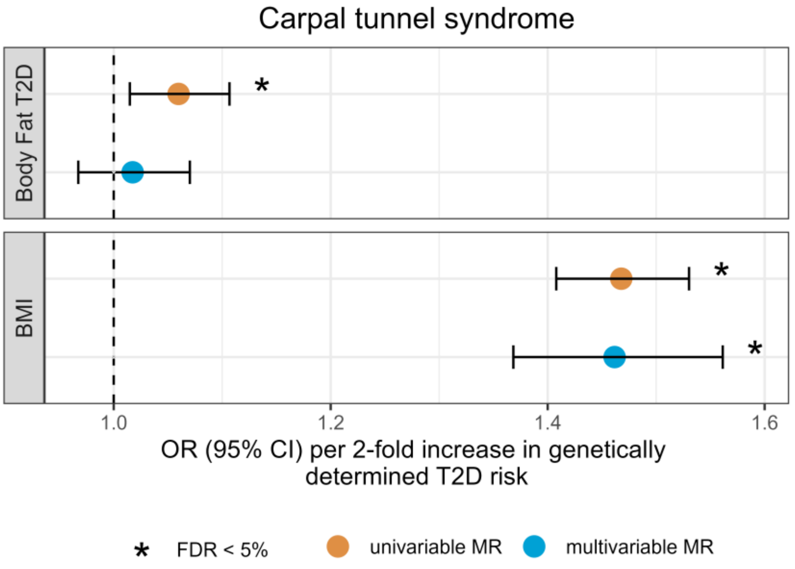

Carpal tunnel syndrome
Body Fat T2D
BMI
*
1.0
1.2
1.4
1.6
OR (95% CI) per 2-fold increase in genetically determined T2D risk
* FDR < 5%
univariable MR
multivariable MR


42

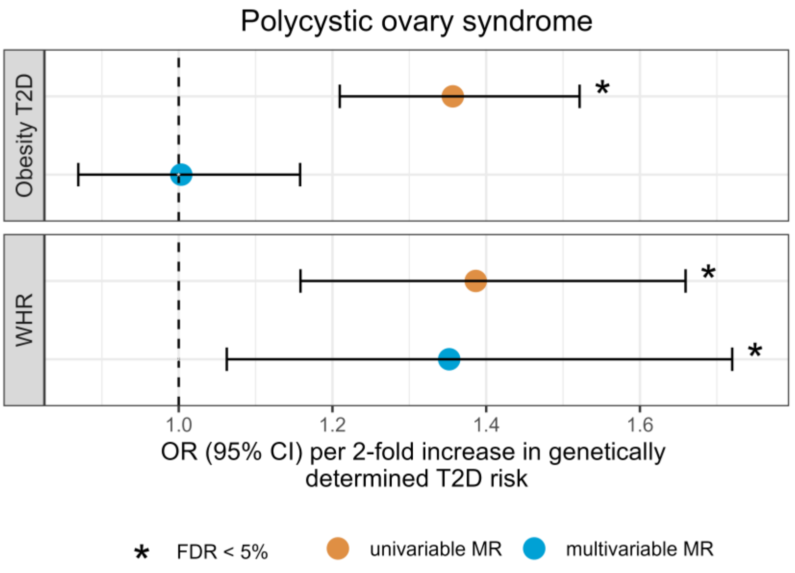

Polycystic ovary syndrome
Obesity T2D
WHR
*
1.0
1.2
1.4
1.6
OR (95% CI) per 2-fold increase in genetically determined T2D risk
* FDR < 5%
univariable MR
multivariable MR

43

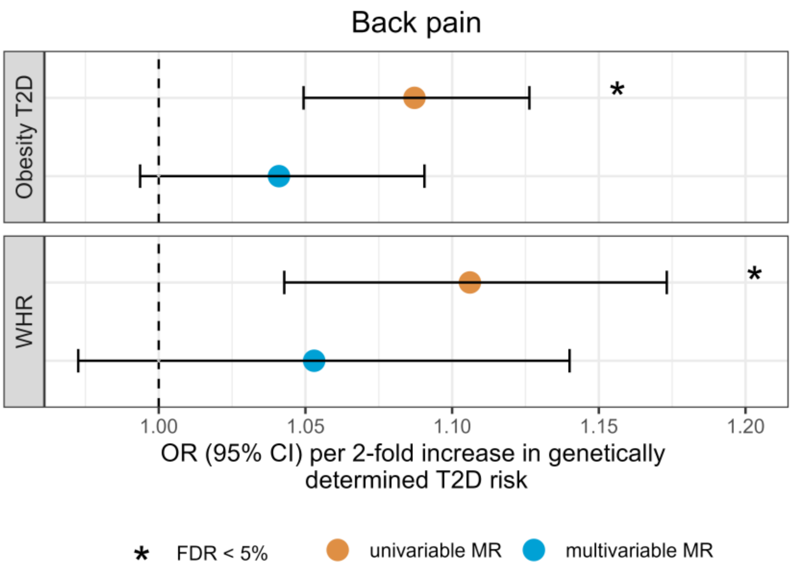

Back pain
Obesity T2D
WHR
*
1.00
1.05
1.10
1.15
1.20
OR (95% CI) per 2-fold increase in genetically determined T2D risk
* FDR < 5%
univariable MR
multivariable MR


44

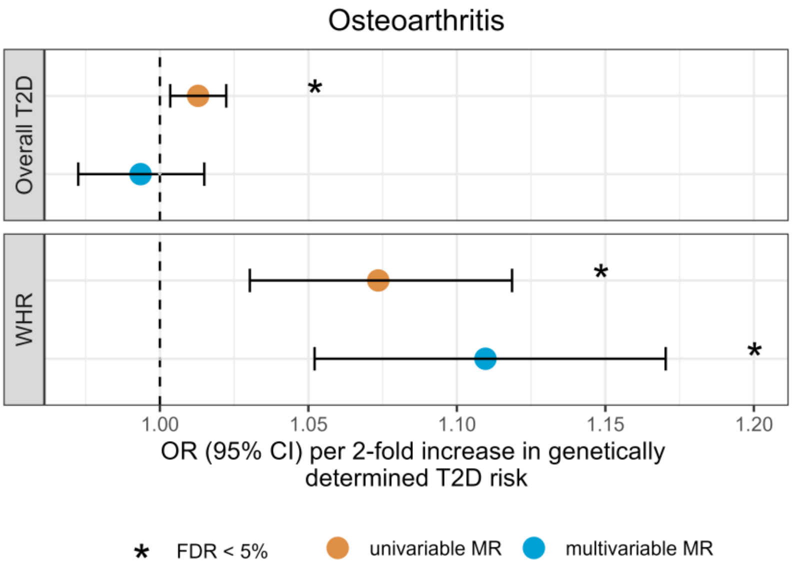

Osteoarthritis
Overall T2D
WHR
*
1.00
1.05
1.10
1.15
1.20
OR (95% CI) per 2-fold increase in genetically determined T2D risk
* FDR < 5%
univariable MR
multivariable MR


45

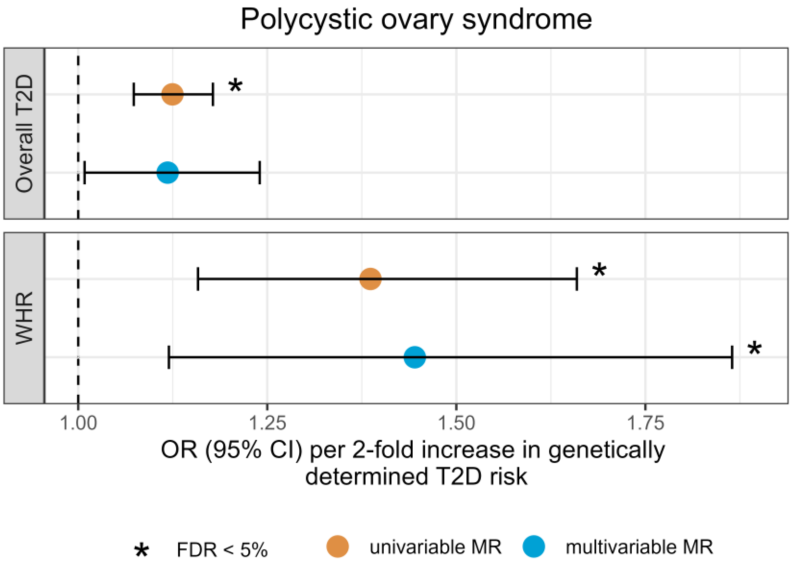

Polycystic ovary syndrome
Overall T2D
WHR
*
1.00
1.25
1.50
1.75
OR (95% CI) per 2-fold increase in genetically determined T2D risk
* FDR < 5%
univariable MR
multivariable MR

46

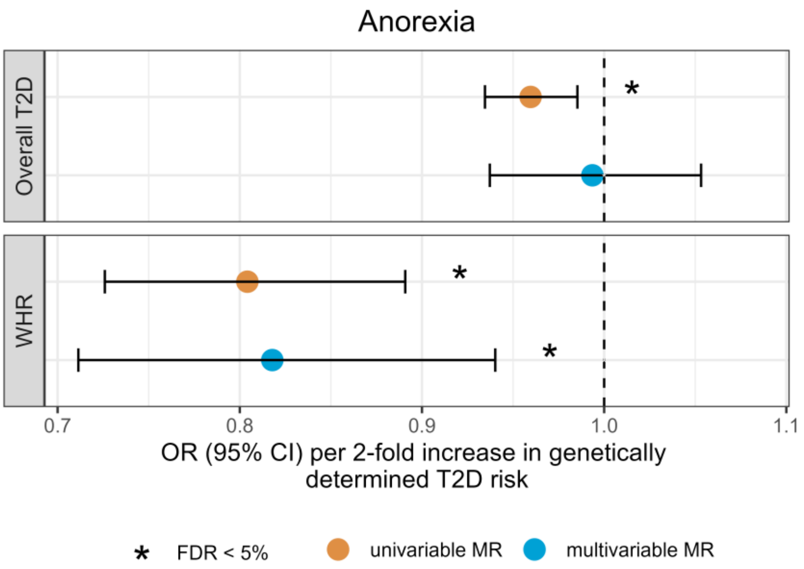
Anorexia
Overall T2D
WHR
*
0.7
0.8
0.9
1.0
1.1
OR (95% CI) per 2-fold increase in genetically determined T2D risk
* FDR < 5%
univariable MR
multivariable MR

47

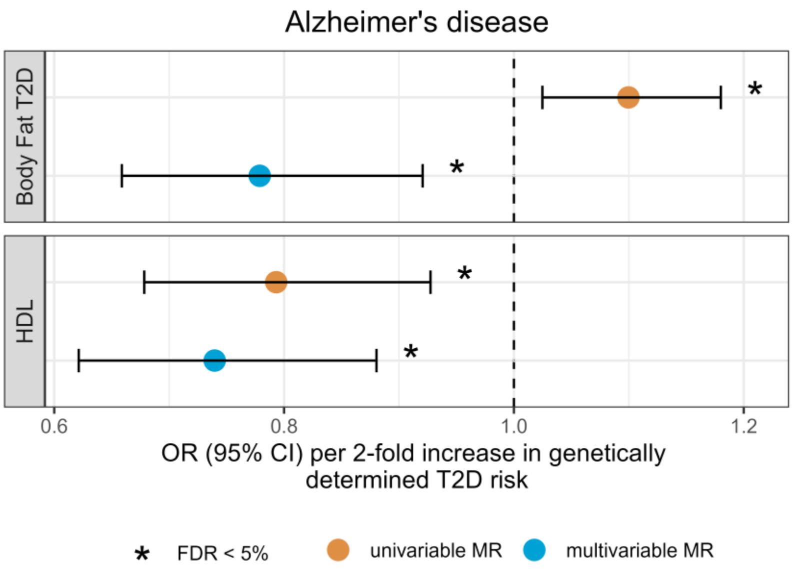
Alzheimer's disease
Body Fat T2D
HDL
*
0.6
0.8
1.0
1.2
OR (95% CI) per 2-fold increase in genetically determined T2D risk
* FDR < 5%
univariable MR
multivariable MR

48

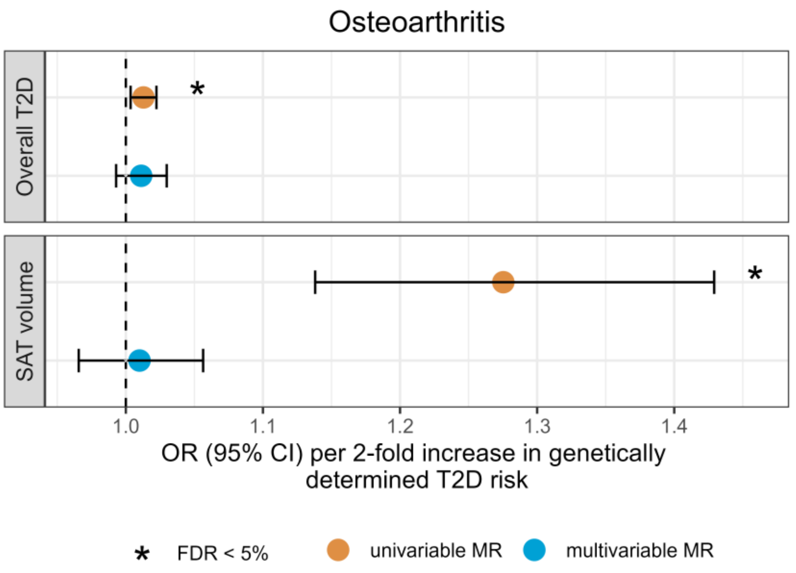
Osteoarthritis
Overall T2D
SAT volume
*
1.0
1.1
1.2
1.3
1.4
OR (95% CI) per 2-fold increase in genetically determined T2D risk
* FDR < 5%
univariable MR
multivariable MR
