## Supplemental Notes for "The effect of type 2 diabetes genetic predisposition on non-cardiovascular comorbidities"

### Supplemental Note

#### Overview of the sensitivity analyses performed to assess the validity of the Mendelian randomization assumptions

| Mendelian randomization (MR) assumption | Sensitivity analysis |
| --- | --- |
| Relevance assumption | - F-statistic - Steiger-filtered inverse variance weighted (IVW) |
| Independence assumption | - Multivariable MR with potential confounders/mediators* - Analysis within genetic similarity groups - Exposure and outcome GWAS summary statistics adjusted for population structure with the removal of related individuals |
| Exclusion-restriction assumption | - MR-Egger intercept test - Heterogeneity assessed by the I^2^ statistic - Same direction of effect across different MR methods:   - Steiger-filtered IVW   - Correlated IVW   - MR-Egger   - Weighted median   - MR-PRESSO |

* Definition of potential confounders/mediators: cardiometabolic traits used to cluster the type 2 diabetes (T2D) variants that have a potential causal IVW effect on the comorbidity at a false discovery rate (FDR) of 5%.

##### Further sensitivity analyses

- Direction of association:
  - Steiger-filter-based directionality test
  - Steiger-filtered IVW
  - Reverse MR
- i.i.d. assumption of genetic instrumental variables (IVs):
  - Different set of T2D IVs: we compared the effect magnitude of our results with alternative approaches to select T2D IVs. We have employed three additional approaches to define T2D IVs: selecting one variant per locus or clumping the independent risk variants either all at once or per cluster
  - Correlated IVW (need FDR-adjusted p-value < 0.05)
- Inverse cluster-stratified MR analysis (Leave-One-Cluster-Out Analysis)

##### Additional analyses to triangulate evidence of the Mendelian randomization analyses

- PheWAS
- MR-Clust

#### Detailed description of the results of the sensitivity analysis to assess the validity of the Mendelian randomization assumptions for the statistically significant IVW results at an FDR of 5%

We describe attenuation as an adjusted IVW estimate that is no longer statistically significant at FDR of 5%. Mutual attenuation describes the case where both exposure estimates are no longer statistically significant at FDR of 5% after the adjustment. Mutual attenuation suggests a shared etiology. The results of the multivariable MR analysis can be found in Supplemental Table 6, the results the sensitivity analyses that assess the validity of the exclusion-restriction criteria can be found in Supplemental Table 2 and Supplemental Table 8, and the results of the Steiger-filter directionality test can be found in Supplemental Table 13.

##### Osteoarthritis

For osteoarthritis, we find evidence of a risk-increasing causal effect of T2D genetic predisposition and genetic risk linked to the obesity cluster. In contrast, T2D genetic predisposition linked to both beta cell clusters is protective to osteoarthritis. We adjust all estimates for cardiometabolic traits that show a statistically significant IVW effect after FDR correction at 5%, namely body mass index (BMI), waist-to-hip ratio (WHR), and subcutaneous adipose tissue (SAT) volume.

All:

- Statistical evidence of horizontal pleiotropy
- Statistical evidence of heterogeneity
- Different directions between MR methods: MR-Egger, penalized weighted median, weighted median, and weighted mode
- Changes direction but remains significant after adjustment for BMI
- No longer significant after adjusting for WHR
- Mutual attenuation: SAT volume

Obesity:

- Statistical evidence of heterogeneity
- Changes direction but remains significant after adjustment for BMI
- No longer significant after adjusting for BMI
- T2D attenuates the effect of WHR

Beta cell +PI/-PI:

- No longer significant after adjusting for BMI, WHR and SAT volume

##### Carpal tunnel syndrome (CTS)

We either observe no or mutual attenuation for all estimates of T2D genetic predisposition on CTS upon adjustment for cardiometabolic traits.

All:

- Statistical evidence of horizontal pleiotropy
- Statistical evidence of heterogeneity
- T2D attenuates the effect of fasting glucose (FG) (changes direction), random glucose (RG) and triglycerides (TG)
- Steiger-filter directionality test: incorrect direction

Obesity:

- Statistical evidence of horizontal pleiotropy
- Statistical evidence of heterogeneity
- T2D attenuates the effect of FG (changes direction), RG and TG

Body fat:

- Statistical evidence of heterogeneity
- Attenuated by BMI
- Mutual attenuation with FG, RG
- T2D attenuates the effect of TG

Lipodystrophy:

- Statistical evidence of heterogeneity
- T2D attenuates the effect of TG, FG
- Mutual attenuation with RG

Metabolic syndrome:

- T2D attenuates the effect of TG, FG
- Mutual attenuation with RG

Residual glycaemic:

- No longer significant after adjusting for BMI
- T2D attenuates the effect of TG, FG
- Mutual attenuation with RG

##### Back pain

The risk-increasing effect of T2D genetic predisposition linked to the obesity cluster on chronic back pain attenuates to zero upon adjustment for BMI and shows mutual attenuation when controlling for WHR, suggesting no direct effect of T2D on disease and shared etiology via obesity-related measures.

Obesity:

- Different directions between MR methods: Steiger IVW, MR-Egger, weighted mode
- Statistical evidence of horizontal pleiotropy
- No longer significant after adjusting for BMI
- Mutual attenuation with WHR

##### Osteoporosis

Individuals genetically similar to Europeans:

- All:
  - Statistical evidence of horizontal pleiotropy
  - T2D attenuates the effect of BMI
- Beta cell +PI: T2D attenuates the effect of BMI
- Obesity: T2D attenuates the effect of BMI
- MetSyn: T2D attenuates the effect of BMI

Meta-analysis across genetic similarity groups:

- All: statistical evidence of heterogeneity
- Obesity: statistical evidence of heterogeneity

##### Rheumatoid analysis

All (Individuals genetically similar to East Asians):

- Different directions between MR methods: MR-Egger
- Statistical evidence of horizontal pleiotropy

##### ADHD

All:

- Statistical evidence of horizontal pleiotropy

Obesity:

- Statistical evidence of heterogeneity
- No longer significant after adjusting for BMI
- T2D attenuates the effect of WHR

Body fat:

- No longer significant after adjusting for BMI

Metabolic syndrome:

- T2D attenuates the effect of WHR

##### Alzheimer’s disease

Body fat:

- Changes direction but remains significant after adjustment for high-density lipoprotein (HDL) cholesterol levels

##### Anorexia nervosa

We identify a robust causal association between overall T2D genetic predisposition and decreased anorexia nervosa risk without any evidence of a cluster-stratified effect.

All:

- No longer significant after adjusting for BMI or WHR

##### OCD

We find evidence of reverse causation for all clusters, estimated by the Steiger directionality test. However, we do not find a statistically significant effect at FDR 5% of genetic predisposition to OCD on T2D risk.

All:

- No longer significant after adjusting for BMI
- T2D attenuates the effect of WHR
- Steiger-filter directionality test: incorrect direction

Obesity:

- No longer significant after adjusting for BMI and WHR
- Steiger-filter directionality test: incorrect direction

Residual glycaemic:

- No longer significant after adjusting for BMI
- Mutual attenuation with WHR
- Steiger-filter directionality test: incorrect direction

##### Glaucoma

All:

- Statistical evidence of heterogeneity
- Mutual attenuation with RG

Residual glycaemic:

- Statistical evidence of heterogeneity
- Mutual attenuation with RG

Beta cell +PI:

- Statistical evidence of heterogeneity
- Mutual attenuation with RG

##### Cataracts

All:

- No longer significant after adjusting for BMI and WHR

Obesity:

- T2D attenuates the effect of BMI and WHR

##### PCOS

All:

- Different directions between MR methods: Steiger IVW, MR-Egger, weighted mode
- Statistical evidence of horizontal pleiotropy
- No longer significant after adjusting for BMI and WHR
- T2D attenuates the effect of SAT volume

Obesity:

- Different directions between MR methods: MR-Egger
- Statistical evidence of horizontal pleiotropy
- No longer significant after adjusting for BMI and WHR
- T2D attenuates the effect of SAT volume

##### Erectile dysfunction

All:

- T2D attenuates the effect of BMI
- Mutual attenuation with WHR

Beta cell +PI:

- Mutual attenuation with BMI and WHR

Residual glycaemic:

- Mutual attenuation with BMI and WHR

##### Asthma

Individuals genetically similar to Europeans:

- All:
  - Different directions between MR methods: MR-Egger, penalized weighted median, weighted median, weighted mode
  - Statistical evidence of horizontal pleiotropy
  - Statistical evidence of heterogeneity
  - No longer significant after adjusting for BMI and WHR
  - T2D attenuates the effect of VAT volume
- Residual glycaemic:
  - Different directions between MR methods: MR-Egger, weighted mode
  - Statistical evidence of horizontal pleiotropy
  - Statistical evidence of heterogeneity
  - No longer significant after adjusting for BMI
  - T2D attenuates the effect of VAT volume
- Obesity:
  - Statistical evidence of heterogeneity
  - No longer significant after adjusting for BMI
  - T2D attenuates the effect of VAT volume

Individuals genetically similar to East Asians:

- All:
  - TG effect changes direction to positive after adjustment for T2D effect, but it remains significant
- Residual glycaemic:
  - No longer significant after adjusting for TG levels
  - TG effect changes direction to positive after adjustment for T2D
- Beta cell +PI:
  - Statistical evidence of heterogeneity
  - TG effect changes direction to positive after adjustment for T2D, but it remains significant

Meta-analysis across genetic similarity groups:

- Statistical evidence of heterogeneity

##### COPD

Individuals genetically similar to Europeans:

- All:
  - Different directions between MR methods: Steiger IVW, MR-Egger, penalized weighted median, weighted median, weighted mode
  - Statistical evidence of horizontal pleiotropy
  - Statistical evidence of heterogeneity
  - T2D attenuates the effect of GFAT volume
  - No longer significant after adjusting for WHR
- Body fat:
  - Different directions between MR methods: Steiger IVW, MR-Egger, weighted mode
  - T2D attenuates the effect of GFAT volume
  - No longer significant after adjusting for BMI
- Obesity:
  - Statistical evidence of horizontal pleiotropy
  - Statistical evidence of heterogeneity
  - T2D attenuates the effect of GFAT volume
  - No longer significant after adjusting for BMI
- Beta cell +PI:
  - No longer significant after adjusting for BMI and WHR
  - Mutual attenuation with GFAT volume

##### Depression

Individuals genetically similar to Europeans:

- Body fat:
  - Different directions between MR methods: MR-Egger
  - Mutual attenuation with BMI
- Obesity:
  - Statistical evidence of heterogeneity
  - No longer significant after adjusting for BMI

#### MR-Clust identifies many clusters with opposite directions of effect

As an additional comparison with our main analysis, we used the MR-Clust method, which groups together IVs with similar causal effect estimates on the outcome trait into distinct cluster[1]. In theory, IVs in these causal clusters may be involved in similar biological pathways, which allows one to identify unique pathways through which exposure has a causal effect on the outcome.

We tested T2D-comorbidity pairs with evidence of a causal relationship from the main analysis in MR-Clust. For each relationship analyzed with the MR-Clust algorithm, we used all 1,289 T2D IVs. MR-Clust clusters were considered causal if they had at least 4 IVs with an 80% or greater probability of being included in the cluster. “Junk” and “Null” clusters identified by MR-Clust were ignored, and any IVs with less than 80% probability of cluster inclusion were removed from the cluster. Following the identification of causal clusters in each T2D-comorbidity pair, we used the IVW method to assess their direction of effect and significance (FDR adjusted p-value < 0.05). The cardiometabolic cluster information for each IV was then merged with each causal cluster to see if there were consistent relationships between an IV’s cardiometabolic cluster and MR-Clust cluster.

We compared our biologically informed results with a statistical approach based on the similarity of the causal estimates implemented in the MR-Clust method[1]. MR-Clust identifies at least one causal cluster for 11 out of 15 comorbidities (73.3%) causally affected by T2D genetic risk (Supplemental Table 14, Supplemental Note Figure 1). For anorexia nervosa, osteoarthritis, ADHD, rheumatoid arthritis, and erectile dysfunction, MR-Clust and our results show concordant directions of effects. For instance, MR-Clust identifies a single cluster of T2D genetic predisposition causally associated with increased risk for anorexia nervosa consisting of 16 IVs (OR=0.23, q-value=1.08x10^-36^). MR-Clust finds two causal clusters of T2D genetic predisposition causally associated with increased osteoarthritis risk, primarily composed of variants assigned to the obesity cluster, supporting our results (Supplemental Note Figure 3). However, many causal clusters for different comorbidities have a highly heterogeneous combination of variants contributing to their observed effect, such as ADHD’s single causal cluster, which includes risk variants from all T2DGGI clusters (Supplemental Note Figures 2-16). A notable difference between both approaches is that MR-Clust often identifies cluster-stratified effects for one disease in opposite directions. In summary, we find consistent directions of causal estimates between our biologically informed approach and MR-Clust for five diseases. The MR-Clust clusters are, however, more complex to interpret biologically and tend to have opposing directions of effect for one comorbidity compared to our biologically informed clustering approach.

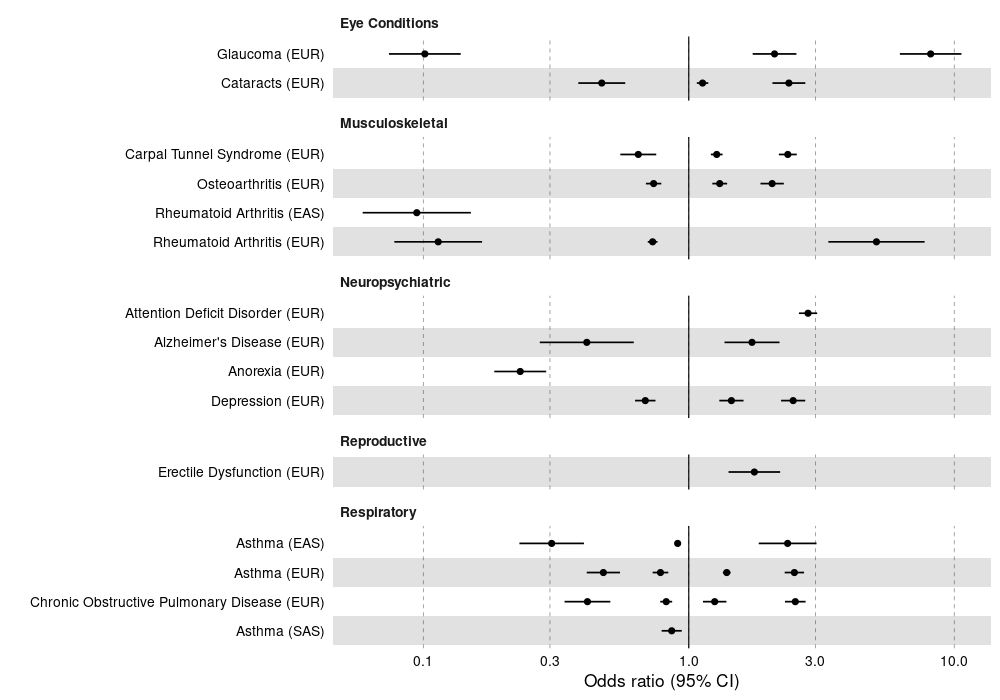

**Supplemental Note Figure 1**: MR-Clust results for the estimated cluster-stratified MR results significant after FDR correction. Each estimate corresponds to one cluster identified by MR-Clust.

**Supplemental Note Figures 2-16**: MR-Clust scatterplots of putative T2D causal clusters on non-cardiometabolic comorbidities.
